# Sex-specific efficacy of acetazolamide for acute mountain sickness. A randomised clinical trial

**DOI:** 10.64898/2026.09.09.26362614

**Authors:** Aijan Taalaibekova, Johanna Roche, Alina Häfliger, Dinah Hertig, Maamed Mademilov, Kamila Magdieva, Azat Bolotbek, Gulzada Mirzalieva, Taomei Li, Kay von Grünigen, Alessandro Vella, Stefanie Zahner-Ulrich, Benoit Champigneulle, Julian Müller, Mona Lichtblau, Samuel Verges, Konrad E. Bloch, Silvia Ulrich, Talant M. Sooronbaev, Michael Furian

## Abstract

**Importance:** Acute mountain sickness (AMS) is a common high-altitude illness partly preventable by acetazolamide, but sex-specific efficacy and tolerability have not been prospectively evaluated.

**Objective:** To test the a priori hypothesis that preventive acetazolamide achieves a 20% greater relative risk reduction (RRR) of AMS in female vs male lowlanders ascending to 3600 m.

**Design:** Randomized, double-blind, placebo-controlled, parallel-group trial conducted from March 2024 to August 2025; participants, outcome assessors, and statistician were blinded until analysis was complete.

**Setting:** Baseline at 760 m in Bishkek and altitude exposure at the Kumtor High Altitude Facility (3600 m), Kyrgyz Republic.

**Participants:** Healthy female and male low-altitude residents (age 18-44 years, BMI 18-30 kg/m2, residence <1000 m); females were premenopausal, eumenorrheic, and not using hormonal contraception. Exclusion criteria included pre-existing disease, regular medication, pregnancy, anaemia, and altitude sojourn >2500 m within 4 weeks.

**Intervention:** Acetazolamide 125 mg or matching placebo twice daily, from 24 hours before ascent and throughout the 2-day, 2-night stay at 3600 m.

**Main Outcomes and Measures:** Primary outcome was the between-sex difference in RRR of AMS, defined as a 2018 revised Lake Louise Score of 3 or higher including headache. Secondary outcomes included absolute risk reduction (ARR) and drug tolerability.

**Results:** Of 303 randomized participants (180 females, 123 males; mean [SD] age, 23 [4] years), AMS incidence under placebo (n=155) was 34% (females 42% vs males 23%; *P*=.02) and under acetazolamide (n=148) was 24% (females 30% vs males 14%; *P*=.02). Overall RRR was 31% (95% CI, 0% to 52%) and ARR was 10% (95% CI, 0% to 22%). Sex-stratified RRR was 27% (95% CI, -9% to 52%) in females and 42% (95% CI, -28% to 74%) in males; between-sex RRR difference, -15% (95% CI, -65% to 57%). Females had a 4.8-fold higher risk of moderate-to-severe drug-related side effects than males.

**Conclusions and Relevance:** Preventive acetazolamide reduced overall AMS incidence at 3600 m, but the pre-specified 20% greater RRR in females (trial powered at 90%) was not observed. The nearly 5-fold higher risk of moderate-to-severe side effects in females warrants further investigation of sex-specific dosing.

**Trial Registration:** ClinicalTrials.gov Identifier: NCT06499727

**Key Points:** *Question:* Does preventive acetazolamide reduce acute mountain sickness (AMS) more effectively in female than in male lowlanders ascending to 3600m?

**Findings:** In this randomized, double-blind, placebo-controlled trial of 303 healthy lowlanders, preventive acetazolamide did not achieve a greater relative risk reduction of AMS in females than in males; the pre-specified 20% between-sex difference, for which the trial was 90% powered, was not observed. However, females experienced a 5-fold higher risk of moderate-to-severe drug-related side effects than males.

**Meaning:** Preventive acetazolamide reduces AMS in healthy lowlanders without demonstrating sex-specific efficacy, but its worse tolerability in females warrants investigation of sex-specific dosing.

## Background

Altitude travel exposes individuals to lower barometric pressures and thereby environmental hypoxia, which leads to reduced arterial blood oxygenation (hypoxaemia)^1^ and induces extensive physiological adaptations to safeguard the body from imminent hypoxaemia-related dysfunction and damage.^2^ Despite these acclimatization effects, moderate hypoxaemia can precipitate acute mountain sickness (AMS), a prevalent altitude-related illness affecting 20-60% of unacclimatised low altitude residents staying overnight at altitudes between 2500 and 4000 m.^3^ AMS manifests within hours to 1-2 days post-ascent, presenting with primary symptoms such as headache, accompanied by malaise, weakness, and fatigue, which typically resolve after 48 hours at high altitude.^4^ Severe cases may compel individuals to seek medical care or abandon their high-altitude plans, and at even higher altitudes AMS might escalate into life-threatening high-altitude cerebral oedema.^5^ Despite the growing body of research related to AMS,^2,4,6^ the pathophysiology and risk factors of AMS remain incompletely understood.

For AMS prevention, current guidelines advocate moderate ascent rates and low sleeping altitudes, supplemented by pharmacological prophylaxes, mainly with acetazolamide.^4,7^ Sex-related physiological differences and a recent post hoc analysis indicate that acetazolamide could be more effective in preventing AMS in females compared with males.^6,8^ Lower blood volume in females may lead to higher acetazolamide plasma concentrations with standard dosing,^9^ potentially enhancing AMS prevention but also increasing side effects.^7,10^

The purpose of this study was to compare the efficacy of preventive acetazolamide for AMS in females compared with males. Findings of this study are expected to provide a foundation for sex-specific high-altitude medical guidelines.

## Methods

### Study Design and SeFng

This randomised, placebo-controlled, double-blind, parallel-group trial was conducted from March 2024 to August 2025 at the National Center of Cardiology and Internal Medicine in Bishkek (760 m), and the Kumtor High Altitude Facility (3600 m), Kyrgyz Republic. The trial was approved by the Ethics Committee of the National Center of Cardiology and Internal Medicine (protocol No. 3-2024) and was pre-registered at ClinicalTrials.gov (NCT06499727). Participants provided written informed consent.

### Participants

Healthy female and male participants, identified by physician-assigned sex at birth, participated. Inclusion criteria for females included premenopausal status, eumenorrheic cycles, non-smoking, body mass index (BMI) between 18 and 30 kg/m2, age between 18 and 44 years, and residence at altitudes <1000 m. Exclusion criteria encompassed pre-existing diseases, regular medication use (including oral contraceptives or other hormonal contraceptives such as hormonal intrauterine devices, vaginal rings, subcutaneous injections, or implants), pregnancy, nursing, anaemia (haemoglobin <10 g/dl), and any altitude sojourn >2500 m within 4 weeks prior to the study. Male participants met the same applicable inclusion and exclusion criteria.

### Intervention

Baseline measurements were performed at 760 m, and participants travelled thereafter for 6 to 8 hours by minibus to a high-altitude location at 3600 m, where they stayed for 2 days and nights. Acetazolamide (125 mg, one capsule in the morning and one in the evening) or identical-looking organoleptic placebo capsules were administered under the supervision of an investigator, starting 24 hours before and during the 2-day stay at 3600 m. Safety considerations are outlined in the **Supplementary Appendix 3.**

### Random Assignment and Blinding

Participants were randomly assigned to acetazolamide or placebo using a computer-generated procedure, with an initial 2:1 allocation ratio (females : males), revised to 1:1 after the pre-specified interim analysis (see below). An independent pharmacist prepared active and identical-looking placebo capsules, packed in neutral boxes. Participants, outcome assessors and statistician remained blinded until data analysis was completed. The investigator in charge of the random assignment was not involved in data collection.

### Questionnaire evaluations

AMS was assessed using the 2018 revised LLS,^11^ which includes self-rated evaluation of four symptoms: headache, fatigue, gastrointestinal discomfort, and dizziness. For comparing the AMS incidences and severity with previous studies, the original LLS score, and the Environmental Symptoms Questionnaire cerebral score (AMSc) were assessed.^12^ The AMS questionnaires were administered at baseline, after arrival and thereafter, twice daily, after awakening and in the evening. In case of symptom worsening, supplemental questionnaires were administered accordingly. Further details are outlined in the **Supplementary Appendix 3.**

### Arterial blood gas analysis

Arterial blood gas analyses (ABG; EPOC, Siemens Healthineers AG, Zurich, Switzerland) were performed on samples obtained from the radial artery after awakening in supine position at 760 m and after the first night at 3600 m. All samples were analysed point-of-care after collection.

### Drug-related tolerability

During the evaluation of AMS symptoms, participants were also asked to rate the occurrence and severity of typical acetazolamide-associated effects, such as paraesthesia (tingling sensation), increased urination, and altered taste, rated as none, mild, moderate, or severe.

### Outcomes and primary hypothesis

The primary outcome of this trial was the difference in the relative risk reduction (RRR) of AMS with preventive 250 mg/day acetazolamide in females compared with males. AMS was defined as a 2018 revised LLS score of ≥3 including headache over the 2-day stay at 3600 m. We hypothesised that 250 mg/day preventive acetazolamide is more effective in reducing the AMS incidence in females compared with males, while staying for 2 days at 3600 m. Secondary outcomes included altitude-, sex- and acetazolamide-related differences in AMS incidences defined by different measures, AMS severity, drug tolerability, arterial blood gases and clinical outcomes.

### Interim analysis

To minimise any unforeseen deviations from the anticipated study progression, such as extreme benefit or harm of acetazolamide or futility and unexpected AMS incidences, a planned interim analysis was scheduled after the completion of the first study year. Further details are outlined in the **Supplementary Appendix 3**.

### Statistical Analysis

Reporting follows the CONsolidated Standards Of Reporting Trials (CONSORT) guidelines.^13^ The study protocol and SAP versions are available in the Supplementary Appendix 1; the CONSORT checklist in the **Supplementary** Appendix 2. Data are summarised by numbers and proportions and means ± SD. The primary analysis was performed on the intention-to-treat population including all randomised participants using the modified Poisson regression with robust variance comparing the sex-related mean difference (95% CI) in the acetazolamide effect, reported as RRR, for the outcome of AMS incidence. In case of missing values in the primary outcome (AMS incidence), the missing data were imputed as AMS positive, for not overestimating the treatment efficacy.

Absolute risk reduction (ARR) was defined as the difference in outcome risk between treatment groups within each sex (acetazolamide vs. placebo). Secondary outcomes were analysed on the per-protocol population, defined as participants with available data. Continuous variables were analysed using mixed linear regression models, with the variable of interest as the dependent variable and drug, location and sex as fixed effects, including the interaction term with drug, location and sex. A p<0.05 or 95% CI excluding zero were considered to indicate statistical significance. Sample size estimation and further details about the statistical analyses are outlined in the **Supplemental Appendices 1 & 3**.

## Results

From March 2024 to August 2025, 551 individuals were screened for eligibility at 760 m in Bishkek, Kyrgyzstan. Of these, 248 did not meet eligibility criteria or declined participation. A total of 303 (59% females) participants were randomised and included in the intention-to-treat analysis of the primary outcome (Figure 1). After randomisation, 12 participants were excluded due to not fulfilling the inclusion criteria or loss to follow-up. Thus, 291 participants were included in the per-protocol analyses of the secondary outcomes. Baseline characteristics of the intention-to-treat (ITT) and per-protocol (PP) populations are presented in Table 1. The mean ± SD age was 22.8 ± 3.9 years and body mass index was 22.4 ± 3.1 kg/m2 in the ITT population. Males were taller and heavier than females. No differences were observed in other demographic characteristics.

**Figure 1.**
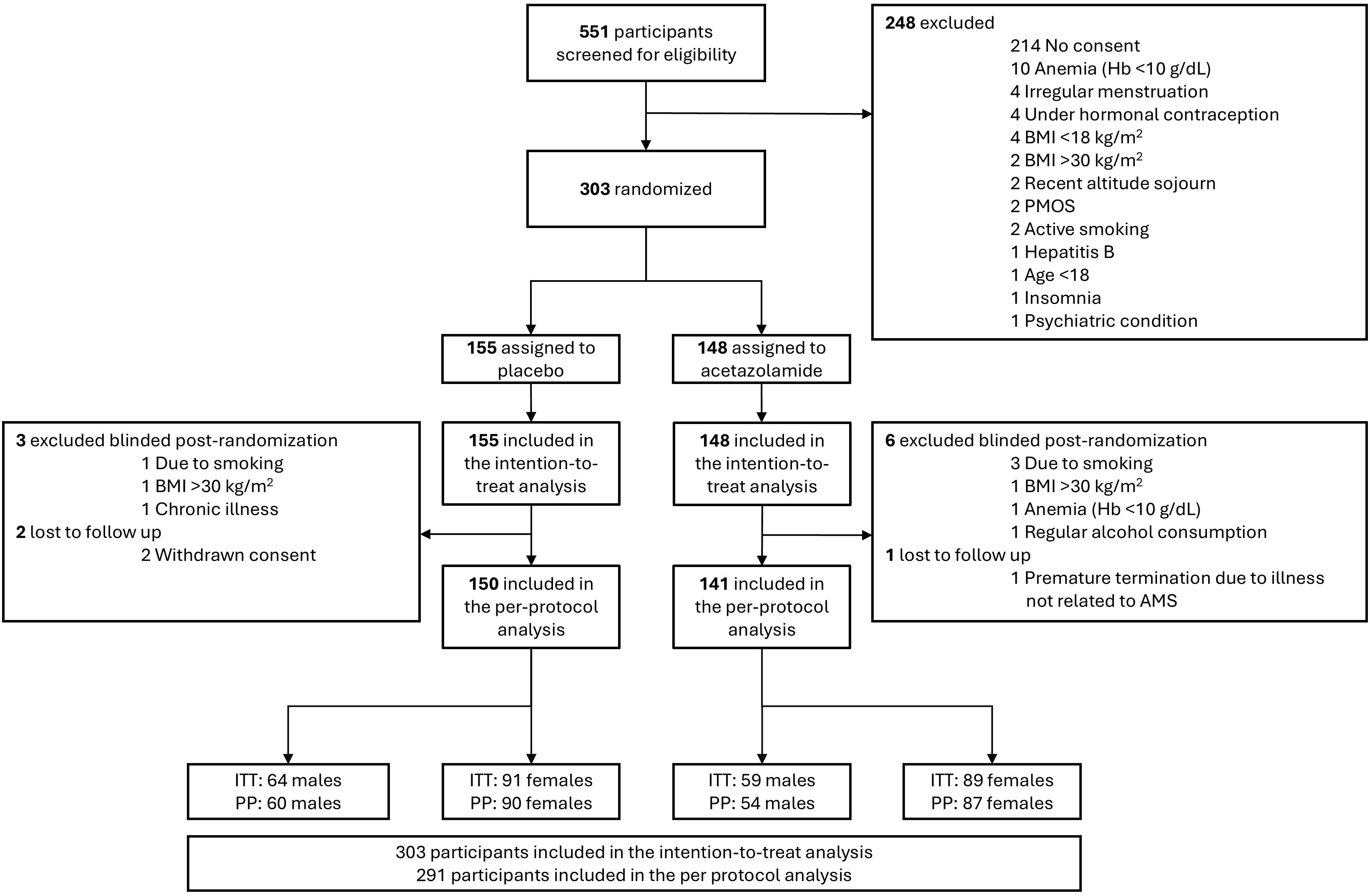
Trial profile. BMI, body mass index; PMOS, polyendocrine metabolic ovarian syndrome; AMS, acute mountain sickness; PP, per-protocol; ITT, intention-to-treat.

**Table 1.**
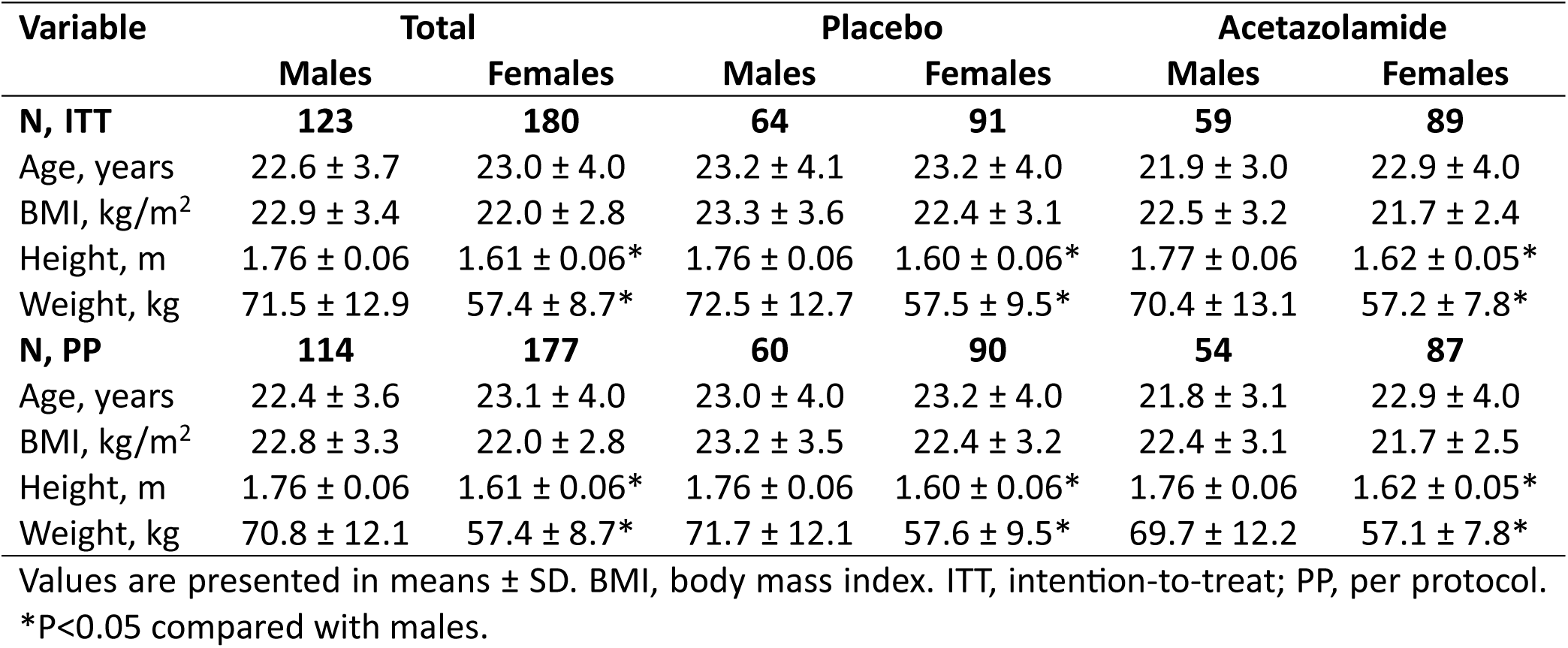
Demographic characteristics.

| Variable | Total |  | Placebo |  | Acetazolamide |  |
| --- | --- | --- | --- | --- | --- | --- |
|  | Males | Females | Males | Females | Males | Females |
| **N, ITT** | **123** | **180** | **64** | **91** | **59** | **89** |
| Age, years | 22.6 ± 3.7 | 23.0 ± 4.0 | 23.2 ± 4.1 | 23.2 ± 4.0 | 21.9 ± 3.0 | 22.9 ± 4.0 |
| BMI, kg/m<sup>2</sup> | 22.9 ± 3.4 | 22.0 ± 2.8 | 23.3 ± 3.6 | 22.4 ± 3.1 | 22.5 ± 3.2 | 21.7 ± 2.4 |
| Height, m | 1.76 ± 0.06 | 1.61 ± 0.06* | 1.76 ± 0.06 | 1.60 ± 0.06* | 1.77 ± 0.06 | 1.62 ± 0.05* |
| Weight, kg | 71.5 ± 12.9 | 57.4 ± 8.7* | 72.5 ± 12.7 | 57.5 ± 9.5* | 70.4 ± 13.1 | 57.2 ± 7.8* |
| **N, PP** | **114** | **177** | **60** | **90** | **54** | **87** |
| Age, years | 22.4 ± 3.6 | 23.1 ± 4.0 | 23.0 ± 4.0 | 23.2 ± 4.0 | 21.8 ± 3.1 | 22.9 ± 4.0 |
| BMI, kg/m<sup>2</sup> | 22.8 ± 3.3 | 22.0 ± 2.8 | 23.2 ± 3.5 | 22.4 ± 3.2 | 22.4 ± 3.1 | 21.7 ± 2.5 |
| Height, m | 1.76 ± 0.06 | 1.61 ± 0.06* | 1.76 ± 0.06 | 1.60 ± 0.06* | 1.76 ± 0.06 | 1.62 ± 0.05* |
| Weight, kg | 70.8 ± 12.1 | 57.4 ± 8.7* | 71.7 ± 12.1 | 57.6 ± 9.5* | 69.7 ± 12.2 | 57.1 ± 7.8* |

Values are presented in means ± SD. BMI, body mass index. ITT, intention-to-treat; PP, per protocol.
\*P<0.05 compared with males.

### Acute mountain sickness

The incidence of AMS at 3600 m, defined according to the 2018 LLS criteria (score ≥3), is shown in Table 2, and the time-to-event in Figure 2. Overall, the AMS incidence in the ITT population was 34% with placebo, and 24% with acetazolamide (p=0.043). The corresponding RRR (95% CI) with acetazolamide was 31% (0 to 52); ARR (95% CI) of 10% (0 to 22); hazard ratio of 0.46 (0.25 to 0.82; p=0.009). In the placebo group, the AMS incidence was significantly higher in females compared with males (42% vs 23%; p=0.02) (Table 2). In the acetazolamide group, AMS incidence remained higher in females compared with males (30% vs. 14%; p=0.02), corresponding to an odds ratio of 2.72 (95% CI 1.20 to 6.15). The RRRs with acetazolamide in females and males were 27% (−9 to 52) and 42% (−28 to 74), respectively, resulting in an RRR-difference in females compared with males, the primary outcome of the study, of −15% (−65 to 57). Furthermore, the corresponding ARRs with acetazolamide were 12% (−2 to 26) in females and 10% (−4 to 23) in males.

**Figure 2.**
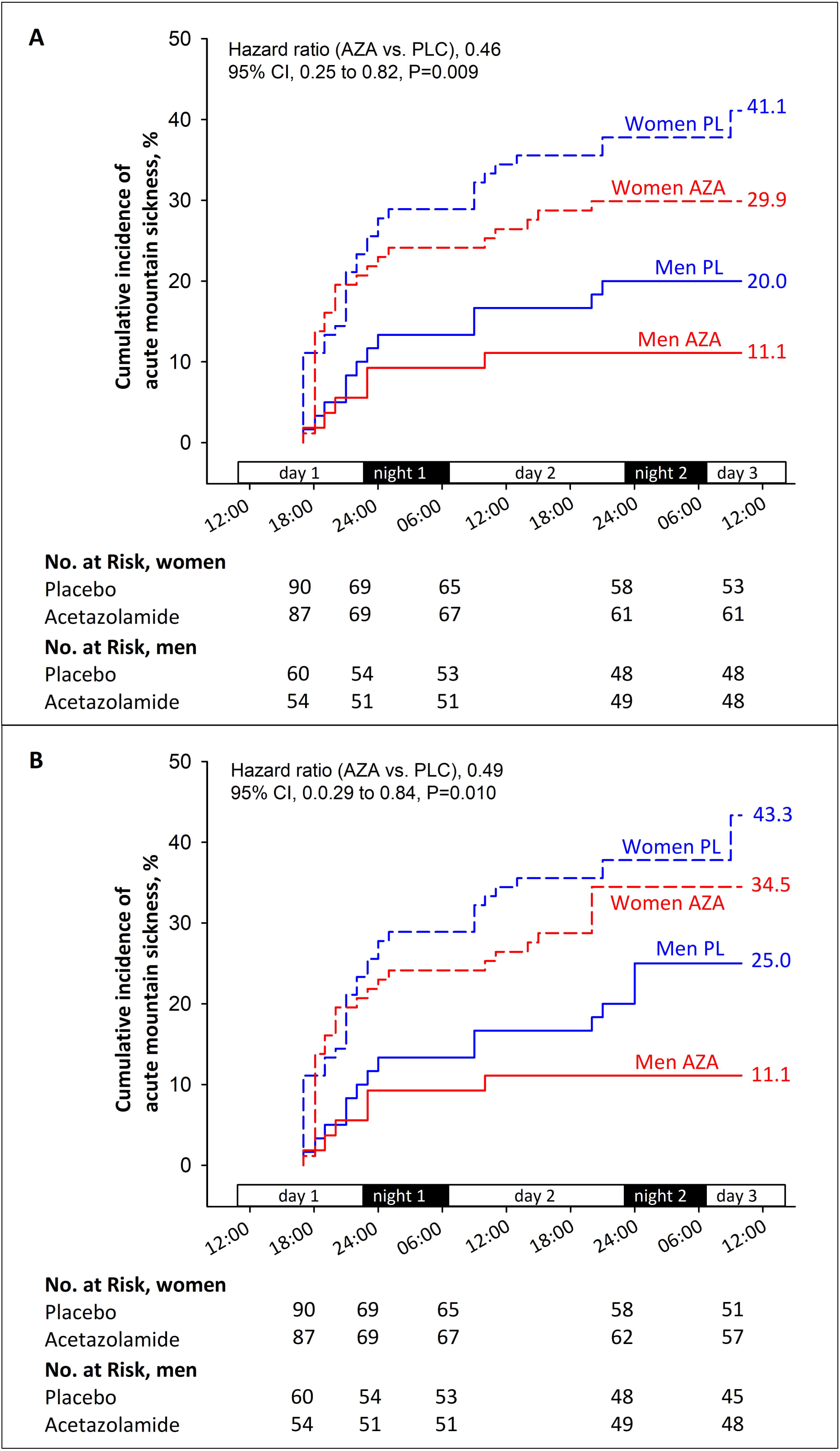
Cumulative incidence of acute mountain sickness over the first 2 days at 3600 m. Panel. **A** Cumulative incidence plot of acute mountain sickness defined by the 2018 Lake Louise criteria (LLS ≥3 with headache). **Panel B** Kaplan–Meier plot using the 1993 Lake Louise criteria (score ≥3 with headache). Corresponding Cox proportional hazard models are shown in **Table S1 & S2**. PLC, placebo; AZA, acetazolamide.

**Table 2.** Acute mountain sickness incidence at 3600 m.

| Variable | Placebo |  | Acetazolamide |  | RRR (95% CI)<br>with ACZ in all<br>participants | RRR (95% CI)<br>with ACZ in<br>males | RRR (95% CI) with<br>ACZ in females |
| --- | --- | --- | --- | --- | --- | --- | --- |
|  | Males | Females | Males | Females |  |  |  |
| **Primary outcome, n ITT** | 64 | 91 | 59 | 89 |  |  |  |
| 2018 LLS $\geq 3$, n ITT (%) | 15 (23%) | 38 (42%)* | 8 (14%) | 27 (30%)* | 31% (0 to 52) | 42% (-28 to 74) | 27% (-9 to 52) |
| **Secondary AMS definitions, n PP** | 60 | 90 | 54 | 87 |  |  |  |
| 2018 LLS $\geq 3$, (%) | 12 (20%) | 37 (41%)* | 6 (11%) | 26 (30%)* | 31% (-2 to 53) | 44% (-40 to 78) | 27% (-10 to 52) |
| 1993 LLS $\geq 3$, (%) | 15 (25%) | 39 (43%)* | 6 (11%) | 30 (35%)* | 29% (-1 to 50) | 56% (-8 to 82) | 20% (-16 to 46) |
| 2018 LLS 3 - 5, (%) | 9 (15%) | 30 (33%)* | 6 (11%) | 12 (14%) | 51% (18 to 71) | 26% (-98 to 72) | 59% (24 to 78) |
| 2018 LLS $\geq 6$, (%) | 3 (5%) | 7 (8%) | 0 (0%) | 14 (16%)* | -49% (-226 to 32) | 100% | -107% (-393 to 13) |
| 1993 LLS 3 - 5, (%) | 10 (17%) | 27 (30%)* | 6 (11%) | 16 (18%) | 37% (-2 to 61) | 33% (-74 to 75) | 39% (-6 to 65) |
| 1993 LLS $\geq 6$, (%) | 5 (8%) | 12 (13%) | 0 (0%) | 14 (16%)* | 12% (-72 to 55) | 100% | -21% (-148 to 41) |
| AMSc score<sup>1</sup> $\geq 0.7$, (%) | 5 (8%) | 24 (27%)* | 1 (2%) | 17 (20%)* | 34% (-14 to 62) | 78% (-91 to 97) | 27% (-28 to 58) |

Values are presented as numbers (proportions) or mean (95% CI). ACZ, acetazolamide; AMS, acute mountain sickness; LLS, Lake Louise Score; ITT, intention-to-treat; PP, per protocol; RRR, relative risk reduction calculated by a modified Poisson regression with robust variance. <sup>1</sup>AMSc score= Environmental Symptoms questionnaire cerebral score ranges from 0 to 5 points (not at all to severe). \* P<0.05 between females and males within the same intervention.

Incidence of AMS using the 1993 LLS and other AMS cut-off thresholds (LLS ≥6, and AMSc score ≥0.7) is also presented in Table 2. The higher AMS incidence in females compared with males under placebo was consistent across different AMS definitions, and in Cox proportional hazard analyses shown in **Supplementary Appendix 3, TableS1 and S2**. However, the treatment effect of acetazolamide remained variable and not statistically significant across these definitions. Mild AMS was more common in the placebo group. Strikingly, moderate-to-severe AMS cases were overall infrequent but occurred significantly more often in females receiving acetazolamide than in females with placebo (Table 2). No consistent difference in sex-related treatment effect was observed across severity categories and AMS definitions (Table 2).

### Drug tolerability and other adverse health effects

Drug-related side effects are presented in Table 3. With placebo, no sex differences in self-reported side effects were detected, except for a higher incidence of mild polyuria in females compared with males. Nevertheless, 45% of females and 25% of males (p = 0.01) reported at least once mild or more severe side effects at 3600 m with placebo. With acetazolamide, females more frequently reported both mild and moderate-to-severe side effects than males. In particular, moderate-to-severe paraesthesia and polyuria were more common in females (25% and 22%, respectively) compared with males (7% and 8%), corresponding to odds ratios of 4.5 (95% CI 1.5 to 13.9) for paraesthesia and 3.1 (1.1 to 8.9) for polyuria. Mild paraesthesia was also more frequent in females (33% vs 19%; OR 2.11 [1.0 to 4.7]), while mild polyuria showed no clear sex difference (30% vs 37%; OR 0.7 [0.4 to 1.5]) (Table 3). Despite these sex-specific patterns within the acetazolamide group, between-drug comparisons did not demonstrate statistically significant differences in males. In females, however, acetazolamide was associated with a markedly higher likelihood of paraesthesia compared with placebo (OR 14.6 [3.3 to 64.3]), whereas the increase in polyuria was not statistically significant (OR 1.6 [0.8 to 3.4]). Moderate-to-severe side effects overall were substantially more frequent in females receiving acetazolamide compared with placebo (43% vs 20%; OR 4.8 [2.0 to 11.2]), while no such difference was observed in males (14% vs 17%; OR 0.8 [0.3 to 2.1]).

**Table 3.** Incidence and severity of self-reported side effects.

|  | Placebo |  |  | Acetazolamide |  |  | Between-drug difference |  |
| --- | --- | --- | --- | --- | --- | --- | --- | --- |
|  | Males<br>N = 64 | Females<br>N = 91 | Odds ratio<br>(95% CI) | Males<br>N = 59 | Females<br>N = 89 | Odds ratio<br>(95% CI) | Odds ratio (95% CI)<br>(ACZ vs. PLC)<br>Males | Odds ratio (95% CI)<br>(ACZ vs. PLC)<br>Females |
| **Paraesthesia** |  |  |  |  |  |  |  |  |
| Mild | 4 (6%) | 8 (9%) | 1.5 (0.4 to 5.0) | 11 (19%) | 29 (33%) | 2.1 (1.0 to 4.7) | 3.4 (1.0 to 11.5) | 5.0 (2.1 to 11.7) |
| Moderate to severe | 2 (3%) | 2 (2%) | 0.7 (0.1 to 5.1) | 4 (7%) | 22 (25%) | 4.5 (1.5 to 13.9) | 2.6 (0.4 to 12.8) | 14.6 (3.3 to 64.3) |
| **Polyuria** |  |  |  |  |  |  |  |  |
| Mild | 9 (14%) | 31 (34%) | 3.2 (1.4 to 7.2) | 22 (37%) | 27 (30%) | 0.7 (0.4 to 1.5) | 3.6 (1.5 to 8.8) | 0.8 (0.5 to 1.6) |
| Moderate to severe | 8 (12%) | 14 (15%) | 1.3 (0.5 to 3.2) | 5 (8%) | 20 (22%) | 3.1 (1.1 to 8.9) | 0.7 (0.2 to 2.1) | 1.6 (0.8 to 3.4) |
| **Change in taste** |  |  |  |  |  |  |  |  |
| Mild | 2 (3%) | 5 (5%) | 1.8 (0.3 to 9.6) | 2 (3%) | 5 (6%) | 1.7 (0.3 to 9.1) | 1.1 (0.2 to 8.0) | 1.0 (0.3 to 3.7) |
| Moderate to severe | 0 (0%) | 1 (1%) | — | 0 (0%) | 1 (1%) | — | — | — |
| **Other** |  |  |  |  |  |  |  |  |
| Mild | 2 (3%) | 7 (8%) | 2.6 (0.5 to 2.9) | 2 (3%) | 6 (7%) | 2.1 (0.4 to 10.6) | 1.1 (0.2 to 8.0) | 0.9 (0.3 to 2.7) |
| Moderate to severe | 2 (3%) | 2 (2%) | 0.7 (0.1 to 5.1) | 1 (2%) | 3 (3%) | 2.0 (0.2 to 19.9) | 0.5 (0.1 to 6.1) | 1.6 (0.3 to 9.5) |
| **Any mild side effect** | 16 (25%) | 41 (45%) | 2.5 (1.2 to 5.0) | 29 (49%) | 54 (61%) | 1.6 (0.8 to 3.1) | 2.9 (1.4 to 6.2) | 1.9 (1.0 to 3.4) |
| **Any moderate to severe side effects** | 11 (17%) | 18 (20%) | 1.2 (0.5 to 2.7) | 8 (14%) | 38 (43%) | 4.8 (2.0 to 11.2) | 0.8 (0.3 to 2.0) | 3.0 (1.6 to 5.9) |

Values are numbers (%) of participants with side effects of any severity measured on a 4-point Likert scale (0=absent, 1=mild, 2=moderate, 3=severe). ACZ, acetazolamide; PLC, placebo.

Severe AMS symptoms and other intercurrent symptoms requiring medical intervention occurred in 10 (8 females and 2 males) participants (5 with placebo, 5 with acetazolamide; **TableS3**). One male participant in the acetazolamide group experienced severe abdominal and back pain, vomiting and microhaematuria. After medical evacuation a kidney stone was diagnosed and treated in a low altitude medical clinic. The participant fully recovered and was discharged after a few days.

## Clinical examination and arterial blood gases

Clinical examinations and arterial blood gases, obtained at 760 m and on the morning after the first night at 3600 m are presented in **TableS4**. Altitude-related and treatment-related changes are shown in Table 4. SpO₂ was lower at 3600 m in both sexes and both treatment groups, with no differences between sexes. Arterial pH increased with altitude in males and females receiving placebo and decreased in both sexes receiving acetazolamide, with a tendency of a stronger effect in females compared with males (Table 4). PaCO₂ and PaO₂ decreased, and haemoglobin concentration and haematocrit increased at altitude in all four subgroups, with greater reductions in bicarbonate in the acetazolamide groups (Table 4).

**Table 4.**
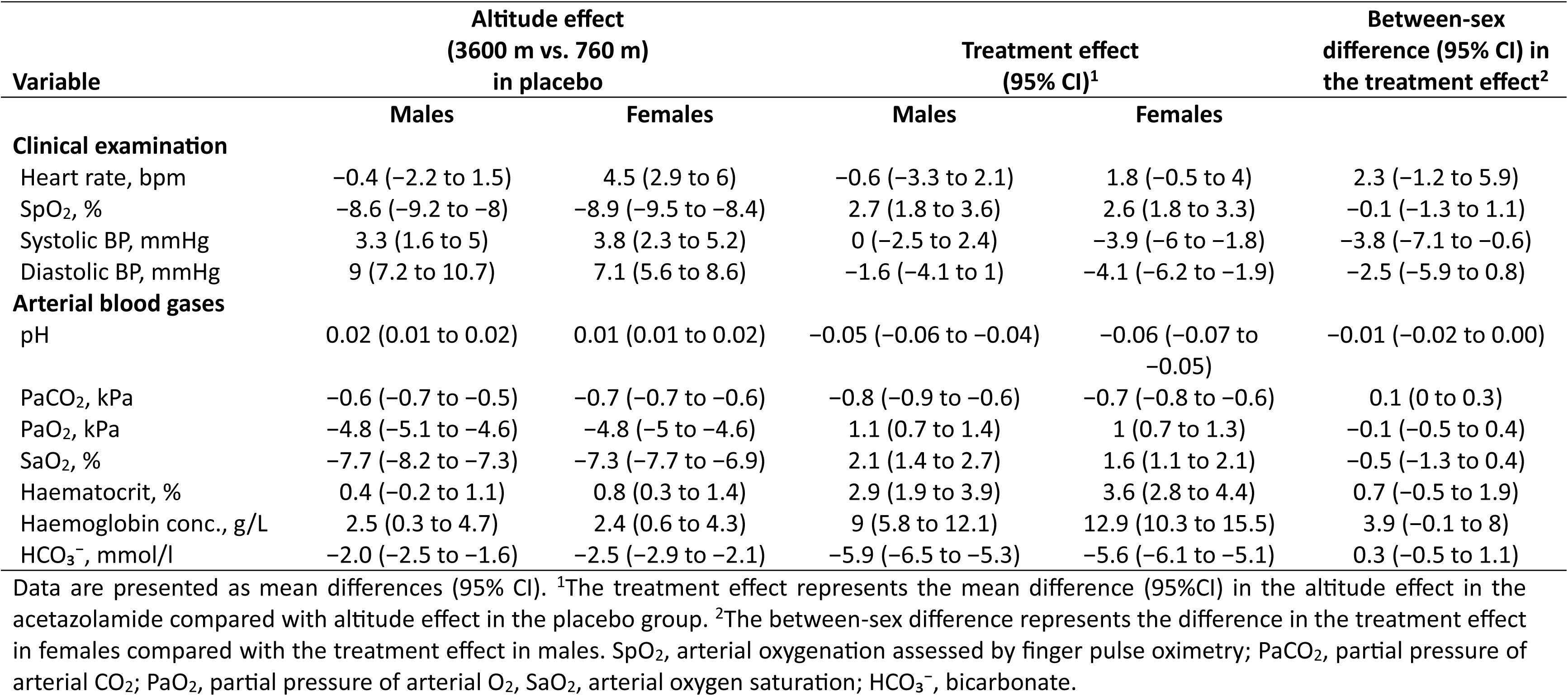
Altitude and treatment effects of secondary outcomes of the per-protocol analysis

## Discussion

In this randomised, double-blind, placebo-controlled parallel-group trial designed and powered to assess sex-specific efficacy of standard dose 250 mg/day acetazolamide for AMS prevention at 3600 m, we report three main findings. First, contrary to our primary hypothesis, the pre-specified 20% greater relative risk reduction of acetazolamide in females compared with males, for which the trial was powered at 90%, was not observed; the point estimate directionally favoured males over females. Second, premenopausal females not using hormonal contraception experienced a consistently higher incidence of AMS than males. Third, female participants self-reported nearly 5-fold more often moderate-to-severe drug-related side effects compared with males. Taken together, these findings indicate that the pre-hypothesized sex-related advantage of acetazolamide in females was not observed, whereas females showed markedly higher AMS susceptibility and a substantially greater burden of drug-related side-effects. These sex-related differences warrant explicit pre-travel counselling and support further trials of sex-adjusted acetazolamide dosing to optimise the benefit–risk balance in this population.

Acetazolamide remains the gold standard for pharmacological AMS prophylaxis. The Cochrane review and meta-analysis by Nieto Estrada et al., the most comprehensive synthesis of randomised trials to date, confirmed that acetazolamide significantly reduces AMS incidence with a risk reduction (RR) of 0.60 (95%CI, 0.39 to 0.94), and supports its use as first-line preventive strategy.^7^ This RR is consistent with our detected RRR of 31%, corresponding to a RR of 0.69. Previous meta-analyses have consistently reported an approximately 48% RRR with acetazolamide compared with placebo, without a clear dose-response relationship across doses ranging from 250 to 750 mg/day,^7,14^ supporting the use of the lowest effective dose in order to improve drug tolerability.

Our findings are consistent with these aggregate estimates. The comparable ARR between sexes is clinically important, as ARR directly reflects the number of individuals benefiting from treatment. Our results are also generally consistent with the recent post hoc analysis from Häfliger et al., which used 375 mg/day acetazolamide in adults older than 40 years and similarly did not identify a significant sex-by-treatment interaction for AMS incidence.^8^ However, the higher acetazolamide dose and older study population including post-menopausal women limit direct comparison.

Although the RRR appeared numerically lower in females (27%) than in males (42%), the wide confidence intervals and absence of a statistically significant interaction preclude definitive conclusions regarding sex-specific differences in treatment efficacy. These findings do not support our primary hypothesis that females might derive greater benefit from acetazolamide because of suspected higher plasma-concentration due to a smaller blood volume relative to body weight. Broader pharmacokinetic literature suggests that females often exhibit higher plasma drug concentrations and prolonged elimination times for many medications, contributing to female-biased adverse drug reactions.^9^ The higher frequency of moderate-to-severe paraesthesia and polyuria observed in female participants receiving acetazolamide in our study is consistent with this concept and aligns with previous reports showing that acetazolamide side effects are common but generally mild.^7,14^ These findings are intriguing since the definition of AMS depends on subjective symptoms that strongly overlap with known acetazolamide-related side effects (e.g., nausea, fatigue, dizziness). Items contained in the LLS itself, particularly fatigue and dizziness, may be inflated by acetazolamide side effects, especially in females who experience these side effects more frequently. This contamination of the AMS outcome by drug toxicity could mask a true sex-specific efficacy advantage of acetazolamide on the cardinal AMS symptom of headache. Further trials dedicated to sex-specific pharmacokinetic and dose-dependency of acetazolamide under hypoxic conditions are warranted.

The higher susceptibility to AMS observed in female participants was consistent across multiple AMS definitions used in this study and aligns with part of the existing literature suggesting increased AMS risk in females.^8,15^ Similarly, observational data from Nepalese pilgrims ascending to 4,380 m showed that females were substantially more frequently affected than males (RR 1.57, 95% CI 1.23–2.00).^16^ Additional support comes from a post-hoc analysis of a recent randomised controlled trial in lowlanders older than 40 years ascending to 3100 m. In the placebo group, 25% of females developed AMS compared with only 8% of males (p = 0.009).^8^ Despite such findings, a recent scoping review by the International Climbing and Mountaineering Federation Medical Commission on sex differences in AMS concluded that because of limited sex-specific data, general prevention and treatment strategies for AMS should be applied regardless of sex.^17^ The authors highlighted several methodological limitations of prior studies, including retrospective or cross-sectional designs, inadequate control for hormonal status and contraceptive use, underrepresentation of female participants, and reliance on older AMS definitions.^17^ Our trial addressed many of these limitations through its randomised, double-blind, placebo-controlled design, the inclusion of only premenopausal females without hormonal contraceptive use, and the use of the revised 2018 LLS. As such, our findings provide robust evidence supporting a sex difference in AMS incidence. The physiological mechanisms underlying the increased susceptibility to AMS in females remain incompletely understood, and are beyond the scope of the present analysis.

Several limitations should be acknowledged. First, the study included young, healthy individuals, which may limit generalisability of the findings to experienced mountaineers, older adults or individuals with comorbidities. Second, AMS diagnosis intrinsically relied on subjective symptoms, which may introduce reporting bias. However, all participants were instructed to report symptoms at 3600 m, and the absence of differences in baseline symptoms between males and females at 760 m (**TableS5**) argues against a simple reporting bias explaining the higher AMS scores in females. Finally, although the study was powered for the primary outcome of between-sex differences in RRR, within-sex effects were secondary endpoints, and the trial was not primarily powered for within-sex efficacy.

## Conclusions

This randomised, placebo-controlled trial was designed and powered to assess sex-specific efficacy of acetazolamide for AMS prevention. The pre-specified 20% greater relative risk reduction in females compared with males at the currently recommended dose of 250 mg/day was not observed, despite 90% power to detect this difference. Acetazolamide reduced AMS incidence overall, consistent with prior evidence. However, following rapid ascent to 3600 m, female participants had a 1.8-fold higher AMS incidence under placebo and a nearly 5-fold higher risk of moderate-to-severe drug-related side effects than males. These sex-related differences in AMS susceptibility and acetazolamide tolerability warrant explicit pre-travel counselling and support further investigation of sex-adjusted dosing regimens to optimise the benefit–risk balance.

## Data Availability

Individual de-identified participant data underlying the results reported in this article will be made available on reasonable request to the corresponding author, beginning 12 months after publication of the secondary outcomes. Data will be shared with researchers who provide a methodologically sound proposal and a signed data access agreement. Proposals should be directed to.

## Author contribution statement

M.F. designed the study. A.T., M.M., K.M., A.B., and T.M.S. conducted study procedures at high altitude. J.R., A.H., D.H., T.L., K.v.G., A.V., S.Z.-U., B.C., J.M., and M.L. contributed to data acquisition and processing. K.v.G., A.T. and M.F. performed the statistical analysis. M.F. and A.T. wrote the first draft. All authors contributed to data interpretation, critically revised the manuscript, and approved the final version.

