## Supplementary Appendix 1 for "Sex-specific efficacy of acetazolamide for acute mountain sickness. A randomised clinical trial"

**Supplement to**

### Sex-specific efficacy and safety of preventive acetazolamide for acute mountain sickness in healthy lowlanders: a randomised, double-blind, placebo-controlled trial.

*Aijan Taalaibekova[1], Johanna Roche[2], Alina Häfliger[2], Maamed Mademilov[1], Kamila Magdieva[1], Azat Bolotbek[1], Dinah Hertig[2], Taomei Li[2], Kay von Grünigen[2], Alessandro Vella[2], Stefanie Zahner-Ulrich[2], Benoit Champigneulle[3], Julian Müller[2], Mona Lichtblau[2], Konrad E. Bloch[2], Silvia Ulrich[2], Talant M. Sooronbaev[1], and Michael Furian[2]*

*[1]National Center of Cardiology and Internal Medicine, Pulmonology Department, Bishkek, Kyrgyzstan; [2]University Hospital Zurich, Pulmonology Department, Zurich, Switzerland; [3]Grenoble Alpes University Hospital, Department of Anaesthesia and Intensive Care, Grenoble, France.*

**This supplement contains the following items:**

1. **Study protocol**
    a. List of study protocol versions and summary of changes
    b. Original study protocol submitted as grant application
    c. Protocol submitted to the Ethics Committee

2. **Statistical analysis plan (SAP)**
    a. List of SAP versions and summary of changes
    b. Original SAP, version 1.0
    c. Revised SAP, version 1.1
    d. Final SAP, version 1.2

1. **Study protocol**
    a. **Summary of protocol changes**

| Date | Version | Explanation |
|---|---|---|
| 30.09.2023 | 1.0 | Protocol submitted to the Ethics Committee |
| 10.06.2024 | 1.1 | The assessment section has been modified but no other changes related to the methodology were made |
| 03.02.2025 | 1.2 | The assessment section has been modified.<br>The target sample size has been increased, based on the blinded interim analysis findings. |

### SEX-SPECIFIC EFFICACY AND SAFETY OF PREVENTIVE ACETAZOLAMIDE FOR ACUTE MOUNTAIN SICKNESS IN HEALTHY LOWLANDERS: A RANDOMISED, DOUBLE-BLIND, PLACEBO-CONTROLLED TRIAL.

**Principal Investigator**

- Dr. sc. ETH Michael Furian, University Hospital Zurich, Pulmonology Department, Zurich, Switzerland

**Swiss Project Partners**

- Prof. Dr. med. Silvia Ulrich, MD, Dept. of Respiratory Medicine, University Hospital Zurich, Zurich, Switzerland
- Prof. Dr. med. Konrad Bloch, MD, Dept. of Respiratory Medicine, University Hospital Zurich, Zurich, Switzerland
- PD Dr. med. Cornelia Betschart, MD, Gynaecology Department, University Hospital Zurich, Switzerland

**International Project Partners**

- Prof. Dr. med. Talant Sooronbaev, MD, Dept. of Respiratory Medicine, National Center of Cardiology and Internal Medicine, Bishkek, Kyrgyzstan
- Dr. sc. hum, MSc. med. Biometry & Statistics Nicola Benjamin, Center for pulmonary hypertension, University Clinic Heidelberg, Germany
- Research Consortium of EXALT – Centre d'Expertise sur l'Altitude, France:
  Prof. Dr. sc. Samuel Vergès (Université de Grenoble), Prof. Dr. Julien Brugniaux (Université de Grenoble), Dr. Benoit Champigneulle (Université de Grenoble), Prof. Dr. Aurélien Pichon (Université de Poitiers), Prof. Dr. Paul Robach (Ecole Nationale de Ski et d'Alpinisme), Dr. Emeric Stauffer (Hospoces Civils de Lyon)

**Contact**

Dr. sc. ETH Michael Furian

University Hospital Zurich

Pulmonology Department

Raemistrasse 100

CH-8091 Zurich, Switzerland

30.09.2023, Version 1.0

### 1. Summary of the research plan

**Background and rationale:** Millions of people travel to high altitude for work or leisure activities and are exposed to reduced inspiratory oxygen partial pressure and hypoxemia that may lead to altitude illness, among which the most common form is acute mountain sickness (AMS). The main AMS symptoms are headache, malaise, weakness, and fatigue. Prospective studies have shown that 20–60% of newcomers at 2500–4000m develop AMS requiring them to take medications, while, at very high altitudes, AMS may progress to high altitude cerebral oedema. Whether females are more susceptible to AMS remains insufficiently understood since no prospective study controlled for sex hormones, use of hormone contraception or assessed menstrual cycle phase (MCP) at altitude. Therefore, females remain underrepresented and poorly characterized in high altitude studies. In addition, the efficacy and safety of 250 mg/day acetazolamide, the standard recommendation for AMS prevention, has never been compared between sexes, although, females have presumably higher acetazolamide plasma concentration due to lower blood volume. Given the known dose-dependent preventive but also side effects of acetazolamide and equal proportion of females and males among mountain travellers, there is an urgent need to conclusively quantify the efficacy and safety of preventive acetazolamide therapy against AMS in females compared to men.

**Overall objectives and specific aims:** The primary objective is to compare the efficacy of preventive acetazolamide treatment against AMS in females compared to men. Additional objectives are to investigate sex- and MCP-related differences in altitude tolerance including AMS incidence as well as efficacy and safety of acetazolamide compared to placebo.

**Methods:** In this randomized, placebo-controlled, double-blinded parallel trial, healthy, premenopausal females and males aged 18 to 44 years, living <1000m will be recruited. Baseline measurements will take place at the Bishkek University Hospital (760m), Kyrgyz Republic; altitude measurements after ascending within 6h by car to 3600m and during a subsequent stay for 2 days and nights. The primary outcome will be the absolute difference in the acetazolamide treatment effect to reduce AMS incidence in females compared to males during the 2-day stay at 3600m. AMS will be defined as a Lake Louise questionnaire score of ≥3 including headache. Secondary outcomes will be derived from respiratory sleep studies, exercise testing, cerebrovascular reactivity assessments and urine and blood drawings. Starting the day before ascent and daily in the following 30 days, females will perform self-measurements of morning urine concentrations of estrone-1-glucuronide, pregnanediol-3-alpha-glucuronide and luteinizing hormone. From these, the Cycle Days and MCP of participants while at 3600m will be derived. The primary analysis will be on the intention-to-treat population using the Cochran Mantel-Haenszel test. Secondary outcomes and the influence of MCP will be analysed by mixed linear or logistical regression models. To detect a clinically relevant absolute 20% efficacy difference of acetazolamide between sexes, using a ratio of 2:1, a power of 90%, two-sided alpha level of 0.05 and accounting for dropouts, a total of 180 females and 90 males are required.

**Expected results:** This project will conclusively assess the efficacy and safety of preventive acetazolamide in premenopausal females compared to males at a representative altitude for many touristic destinations worldwide. Additionally, this study will provide unique insights into clinical and physiological sex- and MCP-related differences in altitude tolerance.

**Impact for the field**: The detailed characterization of females traveling to altitude and using acetazolamide will represent a milestone in high altitude medicine and physiology and will substantially contribute to our understanding of sex-related altitude tolerance and the effect of acetazolamide. Insights about MCP-dependent susceptibility to hypoxia and AMS symptoms will translate to our physiological and mechanistical understanding of the human body.

### 2. Research Plan

#### 2.1. Current State of Research in the Field

On September 27th 2022, the Swiss National Council promoted research and therapy of specific females's diseases.[1] Consequently, the Swiss Confederation has just now commissioned the SNSF to establish four new National Research Programmes, among them "gender medicine".[2] There are numerous diseases that affect females exclusively or in the majority, however, the Committee for Social Security and Health of the National Council writes that research into treatment options is lagging. It is therefore essential that females-specific diseases are identified as such and researched more broadly, like through research programs of the Swiss National Science Foundation (SNF). In addition, guidelines for diagnosis, indication and therapy are needed, as well as a clear definition for quality outcome measurements in females. Too little is known about females' diseases, they remain undiagnosed for a long time and cause unnecessary suffering. As indicated in the literature, acute mountain sickness (AMS) might be one of the diseases affecting females more than men.

##### 2.1.1. Acute altitude-related illnesses in healthy individuals

Mountain tourism accounts for 15% to 20% of the annual global tourism revenue (approximately 296 billion US dollars in 2019),[3] highlighting the popularity of trips to mountainous regions. However, mountain travel exposes the human body to lower barometric pressures and reduced arterial blood oxygenation (hypoxaemia), which requires numerous physiological adaptations to protect the body and organs against hypoxaemia-related dysfunction and damage. However, moderate hypoxaemia can trigger the development of acute mountain sickness (AMS)[4] and other conditions that compromise a stay at altitude, e.g., poor sleep quality and exercise intolerance. AMS is the most important acute altitude-related illness, affecting 20-60% of unacclimatized lowlanders staying overnight at an altitude between 2500 and 4000 m.[5] The main symptoms of AMS that may emerge within 6 hours to 1-2 days after ascent to high altitudes are headache accompanied by malaise, weakness, and fatigue, which often resolves after 48 hours at altitude.[4] However, AMS can force people to take medications or prematurely terminate their stay at altitude. At altitudes >4000 m, AMS might become life-threatening by progressing to high altitude cerebral oedema.[4] The growing number of publications including in the highest-ranked medical journals (i.e. NEJM), emphasize the importance of understanding AMS and other medical conditions at high altitude.[6-8] Despite the growing work, the pathophysiology of AMS remains elusive and consequently, the diagnosis still relies on a pattern of subjective symptoms rather than on objective findings.[9] AMS may develop due to inadequate physiological responses to hypoxaemia such as decreased hypoxic ventilatory response,[10,11] increased sympathoadrenal activity, and insufficient early hypoxic diuresis.[12,13] A history of migraine, a rapid ascent, and previous episodes of AMS are the most robust predictors of AMS. In 2018, analysis of the AMS literature and based on the early onset of AMS symptoms, an international consensus committee composed of 83 international mountain medicine experts concluded that disturbed sleep might be a direct consequence of hypoxia rather than a symptom of AMS, causing a change in the Lake Louise score (LLS),[14] the most common questionnaire used to diagnose AMS. The revised questionnaire now uses the four items, "headache", "gastrointestinal symptoms", "fatigue/weakness", and "dizziness/light-headedness" to diagnose AMS, each rated with a score from 0 (none) to 3 (severe). In the presence of headache, a total score of ≥3 is required to diagnose AMS.

##### 2.1.2. AMS in females

Whether the prevalence of AMS differs between females and males is still being debated and since the modified definition of AMS in 2018, unknown.[15] However, according to the original AMS definition by LLS, a 2019 meta-analysis of 7669 subjects (2639 females) concluded that females have a 1.24-fold risk (95% CI 1.09 to 1.41) of developing AMS at a minimum altitude of 2500 m compared with men.[16] This finding seems logical, since it is known that females do suffer already at low altitude from more headache episodes than men,[17] however, the female physiology under hypoxia remains not at all understood. Important to note, the conclusion of the meta-analysis is debatable since none of the included studies of this meta-analysis assessed the influence of sex hormones (oestrogen, luteinizing hormone, progesterone), menstrual cycle phases (MCP), pre/postmenopausal status or contraceptive use. However, some uncontrolled studies provide important physiological assumptions for sex-related differences in the AMS incidence. Simplified, the sex-related difference in AMS is assumed to be related to sexual hormones and their impact on the physiological response to hypoxia.[16] In accordance with this assumption, progesterone, which is upregulated during the luteal phase, is a well-known respiratory stimulant associated with elevated minute ventilation and decreased $PaCO_2$ during the luteal compared with the follicular phase.[18] Recently, a large study of 336 premenopausal females confirmed these findings and reported lower arterial oxygen saturation ($SaO_2$) and lower hypoxic ventilatory responses in the follicular compared with the early/mid-luteal phase during normobaric hypoxia exposure ($FiO_2$ 0.115, equivalent to 4800 m).[19] These findings are intriguing, since lower $SaO_2$ and lower hypoxic ventilatory response under exercise are risk factors for AMS.[4,19] Another sex-related risk factor for AMS may be low iron stores[20] in females during the follicular phase just after the menstrual bleeding compared to the luteal phase and compared to men.[21,22] Again, no prospective study has investigated the sex-related difference in iron stores on AMS.

##### 2.1.3. Prevention of AMS

Apart from moderate ascent rate and low sleeping altitude, current guidelines recommend acetazolamide, or dexamethasone starting 24 hours before travel to altitude as preventative AMS measures.[23] However, dexamethasone, a glucocorticoid, is mainly recommended for high-risk situations (emergency rescues, military missions) due to its numerous side-effects including hyperglycaemia, immune suppression, and altered mood. Acetazolamide, a carbonic anhydrase inhibitor, is the main recommended and scientifically-proven, pharmacological prophylaxis against AMS.[24] The current recommended dose of 250 mg/day reduces the relative risk of developing AMS by 44%, with a number needed to prevent one case of AMS of 7.[23,24] However, side effects of acetazolamide have been shown to be dose-dependent and include paraesthesia, dysgeusia, polyuria and fatigue.[25] A systematic literature search in PubMed from database inception to September 2023 identified 8 randomized, placebo-controlled, double-blind trials of acetazolamide for prevention of AMS (defined as the primary outcome) using the Lake Louise questionnaire. The studies are summarized in **table 1**. From the 8 identified clinical trials,[7,26-32] only 6 included females.[7,26,27,29,31,32] From the 6 studies including females, only our own trial reported sex-dependent acetazolamide effects on AMS.[7] However, our trial used 375 mg/day instead of 250 mg/Day acetazolamide; was conducted in healthy aged older than 40years and contains a post-hoc analysis. So, in conclusion, none of the only 8 identified clinical trials aimed to investigate sex-differences in the AMS prevention; appropriately characterized the female population, nor used the 2018 version of the LLS to diagnose AMS. Based on the only available trial from our research group in 349 healthy lowlanders, we observed a clear tendency towards a stronger acetazolamide effect in females compared to males (relative risk reduction of 31% in females compared to 21% in men, **figure 1)**.

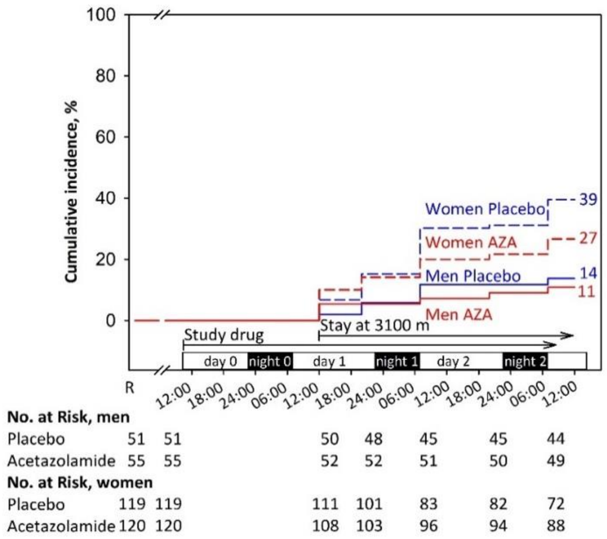

***Figure 1: Incidence of acute mountain sickness and the effect of preventive 375 mg/day acetazolamide therapy starting 24 hours before and during a 2-day stay at 3100 m in healthy females and males older than 40 years (Furian et al., 2022, NEJM Evidence).*** *This post-hoc analysis showed that females were more prone to acute mountain sickness than men; however, no information regarding menopause, menstrual cycle phases, contraceptive use, or hormone supplementation were assessed. Our study also indicated that preventive acetazolamide therapy before ascending to 3100 m might be of relevance for females, but less for men.*

Whether the acetazolamide efficacy against AMS is in fact sex-dependent has not been studied and represent an important gap of knowledge in the field of altitude medicine and physiology. Physiologically, there are arguments supporting such differences in the efficacy, so, lower blood volume in females compared to males is likely to contribute towards higher acetazolamide plasma concentration when prescribing a standard dose of acetazolamide.[33] Higher plasma concentrations are likely causing a dose-dependent preventive acetazolamide effect on AMS but might also induce more side effects.[25,34] The physiological pathway might be that the higher acetazolamide plasma concentration in females versus males promote a stronger metabolic acidosis resulting in a more pronounced hyperventilation, consequently, better mitigating altitude-induced hypoxia – the main driving factor for AMS. Another sex-related factor influencing the acetazolamide efficacy may be related to the increased hypoxic chemosensitivity in males compared to females. Thus, males are more prone to high altitude periodic breathing,[35] whereas this type of sleep-disordered breathing has been shown to cause superficial sleep and worse subjective sleep quality. Acetazolamide has been shown to be highly effective in preventing altitude-induced sleep-disordered breathing and to improve subjective sleep quality – especially in men.[35] So, males were presumably more likely to rate their sleep as “improved” under acetazolamide when using the original Lake Louise questionnaire including the question related to “impaired sleep”. Consequently, the elimination of the item “impaired sleep” in 2018 is likely reducing the preventive acetazolamide effect against AMS in males whereas in females this change in AMS definition has no impact.

**Table 1. Systematic literature analysis of randomized, placebo-controlled, double-blind trials with primary outcome AMS using the Lake Louise questionnaire (all trials cited in PubMed from database inception to September 2023 are included)**

| Item | Design and setting | Participants | Main findings | Limitations of the study |
|---|---|---|---|---|
| **Females included, analysed separately for acetazolamide effect** | | | | |
| Furian et al.[7] 2022 | Placebo-controlled RCT of 375 mg/day AZA on AMS (LLscore ≥3 including headache).<br>LLS at baseline at 760 m; Altitude at 3100 m for 48 hours. | 349 (69% females) healthy lowlanders aged 40 years or more, residing <800 m. | Females<br>39.0% AMS with placebo; 27.0% AMS with AZA; RR of 30.8%, NNT of 8.3.<br>Men<br>14.0% AMS with placebo; 11.0% AMS with AZA RR of 21.4%, NNT of 33. | • **Females not characterized[1]**<br>• **375 mg/day instead of 250 mg/day of AZA**<br>• **1993 LLS version for AMS definition[2]**<br>• **Post-hoc analysis of sex-dependent AZA effect in females ≥40 years.** |
| **Females included, not analysed separately or underpowered for detecting sex-related differences** | | | | |
| Lipman et al.[26] 2018 | Placebo-controlled RCT of 250 mg/day AZA or budesonide on AMS (LLscore ≥3 including headache).<br>LLS at baseline at 1240 m; Altitude at 3810 m for 24 hours. | 70 (47.1% females) healthy adult lowlanders residing <1240 m. | 63.0% AMS with placebo; 43.0% AMS with AZA; RR of 32%, NNT of 5. | • **No sex-dependent AZA effect reported**<br>• **Females not characterized[1]**<br>• **1993 LLS version for AMS definition[2]**<br>• **AZA prevention started on day of ascent**<br>• **Assessment <48 hours at target altitude** |
| Van Patot et al.[27] 2008 | Placebo-controlled RCT of 250 mg/day AZA on AMS (LLscore ≥3 including headache).<br>LLS at baseline at 2000 m; Altitude at 4300 m for 24 hours. | 44 (47.7% females) subjects residing <1600 m | 77.3% AMS with placebo; 31.8% AMS with AZA; RR of 58.9%, NNT of 2.2 | • **No sex-dependent AZA effect reported**<br>• **Females not characterized1**<br>• **No sex-dependent AZA effect reported**<br>• **1993 LLS version for AMS definition[2]**<br>• **Assessment <48 hours at target altitude** |
| Basnyat et al.[29] 2003 | Placebo-controlled RCT of 250 mg/day AZA on AMS (LLS headache + another symptom).<br>LLS at baseline at 4243 m; Altitude at 4937 m. | 155 (32.9% females) trekkers. | 24.7% AMS with placebo; 12.2% AMS with AZA; RR of 50.6%, NNT of 8 | • **No sex-dependent AZA effect reported**<br>• **Females not characterized[1]**<br>• **No baseline <1500 m**<br>• **Trekkers already acclimatized**<br>• **No sex-dependent AZA effect reported**<br>• **Modified AMS definition[2]**<br>• **Assessment <48 hours at target altitude** |
| Chow et al.[31] 2005 | Placebo-controlled RCT of 500 mg/day AZA or Ginkgo biloba on AMS (LLscore ≥3 including headache; also in combination with the clinical assessment score).<br>LLS at baseline at 1230 m; Altitude at 3800 m for 24 hours. | 47 (44.7% females) adult lowlanders residing <1200 m. | 60.0% AMS with placebo; 30.0% AMS with AZA; RR of 50%, NNT of 3.3<br>1 female under placebo developed HAPE. | • **No sex-dependent AZA effect reported**<br>• **Females not characterized[1]**<br>• **No sex-dependent AZA effect reported**<br>• **Modified AMS definition[2]**<br>• **Assessment <48 hours at target altitude** |
| Gertsch et al.[32] 2004 | Placebo-controlled RCT of 500 mg/day AZA or Ginkgo biloba or both on AMS (LLscore ≥3 including headache).<br>LLS at baseline at 4280; Altitude at 4358 m after the first night. | 303 (29.3% females) healthy trekkers. | 34% AMS with placebo; 12% AMS with AZA. RR of 64.7%, NNT of 4.5. | • **No sex-dependent AZA effect reported**<br>• **Females not characterized[1]**<br>• **Dropout rate of 33%**<br>• **No baseline <1500 m**<br>• **Trekkers already acclimatized**<br>• **No sex-dependent AZA effect reported**<br>• **1993 LLS version for AMS definition[2]**<br>• **Assessment <48 hours at target altitude** |
| **No females included** | | | | |
| Hillenbrand et al.[28] 2006 | Placebo-controlled RCT of 250 mg/day AZA on AMS (LLscore ≥3 including headache).<br>LLS at baseline 3440 m; Altitude at 4930 m on arrival. | 400 (0% females) male Nepali porters | 11.1% AMS with placebo; 12.7% AMS with AZA; | • **No females included**<br>• **Dropout rate of 69%**<br>• **No baseline <1500 m**<br>• **Porters already acclimatized**<br>• **1993 LLS version for AMS definition[2]**<br>• **Assessment <48 hours at target altitude** |
| Moraga et al.[30] 2007 | Placebo-controlled RCT of 500 mg/day AZA or Ginkgo on AMS (LLscore ≥3 including headache).<br>LLS at baseline at 0 m; Altitude at 3696 m for 72 hours. | 36 (0% females) healthy adults residing <1000 m. | 56.0% AMS with placebo; 36.0% AMS with AZA; RR of 35.7%, NNT of 5. | • **No females included**<br>• **1993 LLS version for AMS definition[2]** |

[1] Females were not characterized in terms of their reproductive status (pre- / peri- or postmenopausal), use of contraception), hormonal supplementation, or menstrual cycle phase during the stay at altitude. [2]According to the 2018 revised recommendations for defining AMS, a Lake Louise Questionnaire score of at least 3 points including at least mild headache and at least one other symptom of nausea and vomiting, fatigue and/or weakness, dizziness

and/or light-headedness within the first 48 hours at high altitude, is defined as AMS.[15] RR, risk reduction; NNT, number needed to treat; AMS, acute mountain sickness; AZA, acetazolamide; RCT, randomized clinical trial; LLS, Lake Louise Questionnaire.

#### 2.2. Current state of own research

**The current project partners** have successfully collaborated in various research projects in the field of high-altitude medicine and physiology, including the SNF-funded projects (32003B, 192048, 172980, 143875, 122081, IZK0Z3_168254). They have an outstanding expertise in carrying out major clinical trials in healthy and patients with respiratory disease at high altitude places worldwide. To strengthen their collaboration, they launched the "Swiss-Kyrgyz High Altitude Medicine and Research Initiative" endorsed by the University Hospital and the University of Zurich, the Kyrgyz Ministry of Health and the National Center of Cardiology and Internal Medicine (NCCIM) in Bishkek, Kyrgyz Republic.[51] The partners established several research facilities in Kyrgyzstan offering excellent infrastructure for clinical trials: the National Center of Cardiology and Internal Medicine (Bishkek, 760 m), the Tuja Ashu High Altitude Clinic (Tuja Ashu pass, 3'100 m), the Kumtor Gold Mine Operation Facility (Issyk Kul Oblast, 3600 ), and at the Aksay Health Post (3'200 m).

**Dr. sc. ETH Michael Furian** This proposal consolidates and exploits many years of my own experience and expertise in conducting and leading major international studies and clinical trials in Switzerland, Chile, Peru, France, Ecuador, Antarctica and Kyrgyzstan on high-altitude physiology and medicine. This proposal aims to foster my independency in the field of high-altitude medicine and physiology. Until today, I have investigated AMS,[36,37] sleep-disordered breathing,[38,39] exercise performance,[40-42] cerebrovascular reactivity,[43,44] and the effects of acclimatisation and repeated altitude exposure[45] in healthy lowlanders, patients with chronic obstructive pulmonary disease (COPD), and in highlanders permanently living at high altitude.[43,46] I led several randomised placebo-controlled trials as principal investigator investigating the preventive efficacy of nocturnal oxygen therapy and dexamethasone in patients with COPD staying at altitude.[38,39,44,47,48] In a land-mark trial in 185 patients with COPD and 345 healthy subjects ≥40 years staying for 2 days at 3100 m, we recently showed that prophylactic acetazolamide therapy reduces altitude-related adverse health effects in COPD patients and AMS in healthy individuals.[7] This trial further suggested that females were at increased risk of altitude-related illnesses and that the efficacy of acetazolamide might be sex-dependent.

**Prof. Dr. med. Konrad Bloch (KEB)** KEB has a longstanding experience in performing high altitude field studies including in Switzerland (Capanna Regina Margherita, Jungfraujoch, Davos, St. Moritz) and abroad. This research on effects of altitude on cardio-respiratory function,[49,50] exercise, sleep,[51-54] cognitive performance,[55,56] acclimatization and altitude-related illness in healthy individuals[51,57,58] is internationally well recognized. KEB was among the first to perform randomized, placebo-controlled trials in patients with respiratory conditions going to high altitude. These studies established current treatment recommendations for patients with obstructive sleep apnea travelling to altitude.[59-61] KEB has also performed physiological and clinical studies in COPD patients at low and high altitude.[40,41,62] His team performed the first randomized, placebo-controlled trials evaluating prevention of ARAHE in COPD patients with dexamethasone and AZA.[7,36,63-68]

**Prof. Dr. med. Silvia Ulrich (SU)** is director of the Dept. of Respiratory Medicine, University Hospital of Zurich. She has extensive clinical experience in respiratory and internal medicine and in particular in the invasive and non-invasive diagnosis of pulmonary hypertension (PH), treatment of PH, exercise and training enhancing effects of oxygen in cardiorespiratory

diseases and effects of hypoxia in patients with cardiorespiratory diseases and highlanders[63,68-91]. SU conducted several investigator-initiated studies on the pathogenesis and treatment of PH including several placebo-controlled trials on oxygen therapy in PH-patients,[66-68] exercise hemodynamics and exercise limiting factors in PH,[69,70] and the association of sleep disordered breathing with PH, [66] and acute and subacute effects of acetazolamide on hemodynamics and clinical outcomes in PH patients.[78,89,91,92] SU has performed extensive research in high altitude physiology and medicine with a special focus on the pulmonary circulation at rest and during exercise and patients with pulmonary vascular disease going to altitude and extensive collaborative studies with the applicant and collaborators.[69,70,85,86,93-95]

**Prof. Dr. med. Talant Sooronbaev (TS)** is director of the National Center of Cardiology and Internal Medicine (NCCIM) and head of the Dept. of Respiratory Medicine in Bishkek, Kyrgyzstan. He is Chief Pulmonologist in the Ministry of Health of the Kyrgyz Republic and member of the National Medical Research Council. TS directs the research facilities operated by the "Swiss-Kyrgyz High Altitude and Medicine Initiative". He performed various studies on cardio-respiratory disease in Kyrgyz highlanders[96-98] and is a main collaborator in various altitude studies conducted by our project team. TS has also studied the prevalence of COPD in high altitude residents in the Burden of Obstructive Lung Disease study (www.boldstudy.org) and in the FRESH AIR study http://www.theipcrg.org/freshair.

**PD Dr. med. Cornelia Betschart (CB)** is a gynecologist and obstetrician at the University Hospital Zurich since 2008 and board-certified urogynecologist since 2016. She is interested in AMS as a long-standing member of the Swiss Alpine Club. Scientifically she conducts several clinical multicenter observational, and prospective randomised placebo-controlled trials as principal investigator, also granted by the SNF and private foundations. She promotes inter-professional and inter-disciplinary research as founding member of the International Collaboration for Harmonising Outcomes, Research, and Standards in Urogynaecology and Women' Health (CHORUS), as president of the Swiss Urogynecological Association, steering-board member of Gender medicine USZ, co-founder and leader of Pelvic Floor Center USZ and deputy chairwoman of the Department of Gynecology USZ.

**Dr. sc. hum, MSc. med. Biometry & Statistics Nicola Benjamin (NB)** is the Head of study coordination and scientific project management of the Center for pulmonary hypertension at the University Clinic of Heidelberg, Germany. NB has a broad and extensive knowledge on clinical trial design and statistical analyses in the field of Pulmonology (including hypoxia and hypoxemia).[99-101] She contributed to the trial design and statistical considerations of this application, furthermore, NB will supervise the creation of the statistical analysis plan, the interim analysis and the final statistical analyses of this project.

**EXALT Research Consortium** is composed of outstanding high-altitude researchers situated in France. These researchers, namely Prof. Dr. Samuel Vergès (Université Grenoble Alpes), Prof. Dr. Julien Brugniaux (Université Grenoble Alpes), Dr. Benoit Champigneulle (Université Grenoble Alpes), Prof. Dr. Aurélien Pichon (Université de Poitiers), Prof. Dr. Paul Robach (Ecole Nationale de Ski et d'Alpinisme), Dr. Emeric Stauffer (Hospoces Civils de Lyon), have conducted landmark studies in France and in Peru (Expedition 5300 m).[102-106] They are specialized in comprehensive physiological and basic research at altitudes up to 5300 m. Due to SNF Postdoc.Mobility grant in 2020, the main applicant joined this team in Peru, La Rinconada in 2021 and invited them to research studies in Kyrgyzstan.[107,108] They will strongly contribute to the physiological project aims to understand physiological and mechanistic sex-differences in altitude tolerance and mechanisms of action of acetazolamide.

##### 2.2.1. Insights from two preceding pilot studies in females

Assessing sex-related differences at high altitude require meticulous planning, translational knowledge in interdisciplinary research fields and established high altitude research facilities. Therefore, to gain important insights into the expected challenges of high-altitude studies in females, and to enhance the feasibility of the proposed project, the main applicant of this proposal conducted two pilot studies in overall 42 healthy premenopausal females in 2022. All females were instructed to monitor their urinary hormone concentrations for 30 consecutive days by the *Full Cycle Hormone Insights Kit* (Proov, MFB Fertility Inc., CO, US) starting the day before traveling to 3100 m or 3600 m and staying there for 2 days/nights. These pilot studies revealed excellent hormone monitoring adherence, with 1201 of 1230 (97.6%) successful hormone analyses, providing a robust and reliable hormone measurement technique. The averaged and smoothed hormone profile illustrated in **Figure 2, Panel A** shows that ascending to high altitude, independent of the MCP, results in the expected random distribution of participants throughout the cycle days (**Figure 2, Panel B**). This important finding confirms that any large-scale randomized clinical trial does not require to schedule altitude ascents based on the cycle day or MCP. The equal distribution of the participants throughout the Cycle Days are a clear strength of the study design, since it avoids any biased conclusion based on comparisons of a few isolated Cycle Days in the luteal compared to follicular MCP (in case randomization is based on specific Cycle Days or MCP).

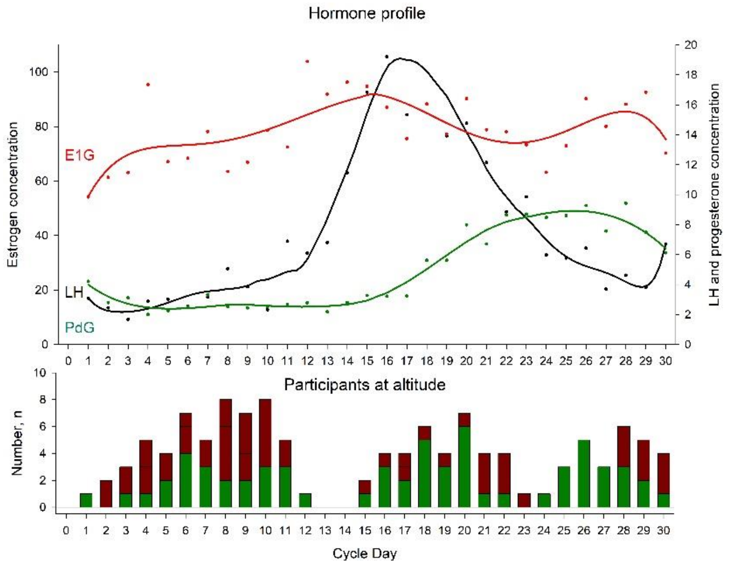

***Figure 2: Urinary hormone profile from 42 females travelling to high altitudes. Panel A:*** *Females daily monitored their urinary hormones E1G, LH and PdG for 30 consecutive days. Based on their first day of the menstruation, the individual Cycle Day at altitude ascent was calculated. Hormone concentrations were averaged for each Cycle Day and a curve was fitted for each hormone.*

***Panel B:*** *The total number of participants staying at altitude and in relation to their individual Cycle Day. The green proportion of the bars represent females without AMS; red proportion of the bars represent females with diagnosed AMS.*

In addition, when comparing the findings at 3100 to 3600 m (**Table 2**), the pilot studies provide important quantitative data related to AMS incidences. Therefore, an AMS incidence of 57% can be expected in premenopausal females staying for 2 days/nights at 3600 m.

**Table 2. Main findings of the two pilot studies.**

| | Group 1 (n = 21) | | | Group 2 (n = 21) | | | Mean difference in altitude effect (3600 m vs 3100 m) (95% CI) |
|---|---|---|---|---|---|---|---|
| | **760 m** | **3100 m** | **Mean altitude effect (95% CI)** | **760 m** | **3600 m** | **Mean altitude effect (95% CI)** | |
| **Clinical examination** | | | | | | | |

| | | | | | | | |
|---|---|---|---|---|---|---|---|
| Systolic BP, mmHg | 106 ± 2 | 106 ± 2 | 0 (-3 to 3) | 104 ± 2 | 105 ± 2 | 1 (-2 to 5) | 1 (-3 to 6) |
| Diastolic BP, mmHg | 73 ± 2 | 74 ± 2 | 1 (-3 to 5) | 70 ± 2 | 76 ± 2 | 6 (2 to 10)* | 5 (0 to 10) |
| Heart rate, bpm | 85 ± 2 | 87 ± 2 | 2 (-3 to 7) | 78 ± 2 | 95 ± 2 | 17 (12 to 22)* | 15 (8 to 23)* |
| $SpO_2$, % | 96.1 ± 0.4 | 93.0 ± 0.4 | -3.1 (-4.2 to -2.1)* | 97.3 ± 0.4 | 86.8 ± 0.4 | -10.5 (-11.6 to -9.4)* | -7.3 (-8.8 to -5.8)* |
| Acute mountain sickness | | | | | | | |
| 2018 Lake Louise Score | 0.0 ± 0.3 | 0.3 ± 0.3 | 0.3 (-0.4 to 1.0) | 0.0 ± 0.3 | 1.9 ± 0.3 | 1.9 (1.2 to 2.6)* | 1.6 (0.6 to 2.6)* |
| 2018 AMS incidence, % | 0% | 43% | | 0% | 57% | | |

BP, blood pressure; $SpO_2$, arterial oxygen saturation assessed by finger pulse oximetry.

#### 2.3. Detailed research plan

##### 2.3.1. Purpose

The primary purpose of this project is to compare the efficacy of preventive acetazolamide treatment against AMS in females compared to men. An important secondary purpose will be to compare the AMS incidence in females versus males at 3600 m. Additional purposes are to investigate sex- and MCP-related differences in altitude tolerance and efficacy of acetazolamide prevention and to underpin clinical findings with various physiological measures providing important mechanistic insights. Findings will improve our understanding of AMS pathophysiology and will provide scientific evidence whether sex-related recommendations on altitude travel and prevention of altitude illness are necessary.

##### 2.3.2. Primary Hypothesis

The primary hypothesis will be that, during the time course of 2 days/nights at 3600 m, preventive 250 mg/day acetazolamide therapy starting 24 hours before ascending, will reduce the AMS incidence significantly more in females compared to men. AMS will be defined as a Lake Louise questionnaire score of ≥3 points including headache.[14]

##### 2.3.3. Additional Hypotheses

Altitude effects in females

1) Females under placebo staying at 3600 m experience altitude-induced exercise intolerance after arrival at 3600 m compared to 760 m assessed by a maximal cardiopulmonary bicycle exercise test.

Altitude effects between females and men

2) Females under placebo have a higher AMS incidence compared to males under placebo during the time course of 2 days and nights at 3600 m.
3) Females under placebo have less sleep-disordered breathing during the first night at 3600 m compared to males under placebo assessed by respiratory polygraphy.

Acetazolamide effects in females

4) Acetazolamide therapy mitigates altitude-induced exercise intolerance compared to placebo.

Acetazolamide effects between females and men

5) The acetazolamide plasma concentration in the morning after the first night at 3600 m is higher in females compared to men.

6) Perceived acetazolamide-related side effects are higher in females compared to males during the time course of 2 days and nights at 3600 m.

MCP-related differences

7) In females under placebo staying at 3600 m during the luteal MCP, the AMS incidence; altitude-induced exercise intolerance and cardiac repolarization disturbances are lower compared to females during the follicular MCP.

##### 2.3.4. Study design and setting

This is a randomized, placebo-controlled, double-blind, parallel trial (**Figure 3**) conducted over 2 consecutive summer seasons. Healthy females and males (according to physician assigned sex at birth) will perform measurements at the National Center of Cardiology and Internal Medicine, Department of Respiratory Medicine, Bishkek (760 m), Kyrgyzstan and in the Kumtor High Altitude Facility (3600 m), Kyrgyzstan. After written consent of female participants, they will be randomized to either acetazolamide or placebo and will start the sexual hormone monitoring in the morning urine as described in the **section 2.3.6** below. Baseline measurements at 760 m will be conducted 24 hours before ascending to 3600 m and altitude measurements will be performed while staying for 2 days and nights at 3600 m. Male participants will undergo the same measurements at 760 and 3600 m but will not monitor hormones. Transfers between locations will be performed by minibus within 6 hours.

##### 2.3.5. Participants

Inclusion criteria for female participants are premenopausal, eumenorrheic, non-smoking, healthy females with a BMI >18 kg/m$^2$ and <30 kg/m$^2$, aged 18 to 44 years, and who live at altitudes <1000 m. Exclusion criteria are any pre-existing diseases, regular intake of medication (including oral contraceptives), other types of contraceptives (hormonal intrauterine device, vaginal ring, subcutaneous injections or implants, among others), pregnancy or nursing, anaemic (haemoglobin concentration <10g/dl), and any altitude trip <4 weeks before the study. Additionally, all included females require a mobile phone compatible with the hormone monitoring app *Proov*. Male participants fulfilling the above-mentioned, male-applicable inclusion and exclusion criteria will be recruited. Participants will be recruited by several teams advertising the study at various universities in Bishkek and in the surrounding villages. This recruitment procedure has been applied in several previous randomized clinical trials and has been proven to be extremely successful. The protocol will be submitted to the Ethic Committee

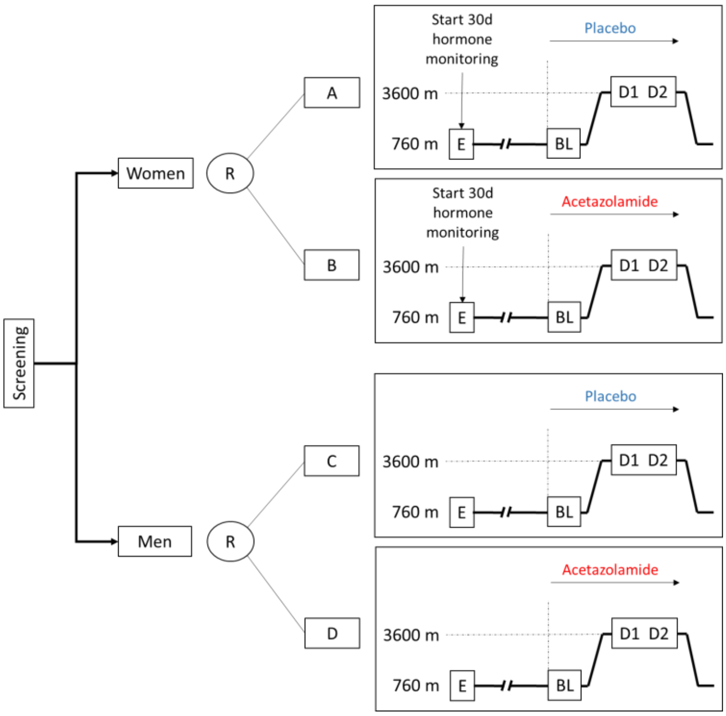

***Figure 3: Study design.*** *Preventive 250 mg/day acetazolamide will be administered 24 hours before travelling and staying for 2 days/ nights at 3600 m. AMS incidence will be assessed at 3600 m. Female participants will monitor urinary levels of sexual hormones for 30 days allowing to determine the MCP during the stay at 3600 m. E, study entry; BL, baseline; R, randomization; D1 – 2, day 1 to 2.*

of the National Center of Cardiology and Internal Medicine, Bishkek, Kyrgyzstan and to the Cantonal Ethics Committee of Zurich, Zurich Switzerland. Participants will be asked to provide written informed consent.

##### 2.3.6. Measurements during study visits

The schedule for the measurements is illustrated in **Table 3**.

**Table 3. Assessment schedule**

| | Screening | BL, Day 1, 760 m | BL, Day 2, 760 m | Day of ascent, 760 m / 3600 m | Day 1, 3600 m | Day 2, 3600 m |
|---|---|---|---|---|---|---|
| **Time before ascent to 3600 m** | **up to -5 month** | **-1 week** | **-1 day** | **0** | **+ 1 day** | **+2 days** |
| **Assessment** | | | | | | |
| Informed consent | x | | | | | |
| In-/exclusion criteria | x | | | | | |
| Reproductive status | x | | | | | |
| Clinical examination | x | x | x | x | x | x |
| AMS questionnaires | | x | x | x | x | x |
| Respiratory polygraphy | | x | | x | x | |
| Blood drawing | | x | | | x | |
| Hypoxic ventilatory response | | x | | | | |
| Cerebrovascular reactivity | | x | | | x | |
| 12-lead ECG | | x | | x | | |
| Maximal CPET | | x | | x | | |
| Fluid homeostasis | | | | x | x | x |
| Hemorheology | | x | | | x | |
| OpCo CO-rebreathing | | x | | | x | |
| Medical intervention | | | → | → | → | → |
| Urinary hormone testing | | → | → | → | → | → |

BL, baseline; ECG, electrocardiogram; CPET, cardiopulmonary exercise testing.

Screening visit

After providing written informed consent, a complete medical history will be obtained. A reproductive status questionnaire will assess regularity of menstrual periods, menstrual cycle length, menarche, pregnancy and breastfeeding, use of contraception, use of other hormone supplementation and known gynecologic or other medical conditions. Inclusion and exclusion criteria will be applied. Physical examination will include weight, height, blood pressure, heart rate, and cardiac and pulmonary auscultation. A pregnancy test will be performed.

History, symptoms, and clinical examination

Repeated physical examinations will include weight, height, blood pressure, heart rate, and cardiac and pulmonary auscultation. AMS will be assessed using the current and the 1993 version of the Lake Louise score[14,109] and the environmental symptoms questionnaire (AMS-c).[110] Subjective sleepiness and sleep quality will be assessed using the Karolinska Sleepiness scale and a 100-mm visual analogue scale. A standardised headache diary including information about the date, time, intensity (1 to 10), preceding symptoms, triggers and relief of headache will be distributed. This diary will be incorporated in the 30-day urinary hormone monitoring and will also be distributed to male participants.

Urinary hormone monitoring

Females will monitor the urinary hormone concentrations of estrone-1-glucuronide (E1G), pregnanediol-3-alpha-glucuronide (PdG) and luteinizing hormone (LH) for 30 consecutive days by the FDA approved *Full Cycle Hormone Insights Kit* (proov, MFB Fertility Inc., CO, US) starting before traveling to 3600 m. The *proov* hormone monitoring kit has been used in previous research studies and has been validated against other methods for hormone monitoring.[111] Measurements will be standardized in the morning, after awakening and before drinking or eating breakfast. The *proov* multi-hormone test strip lateral flow assay uses gold nanoparticles and buffered sample pads designed to adjust for pH and hydration levels, filters unwanted particulates and binds contaminants in urine that may interfere with the accuracy of the test. The strip contains three test lines and one control line, corresponding to E1G, PdG and LH (beta subunit). After waiting for 10 minutes, the user provides a photo of her urine test strip with the *proov* app. The application server uses machine learning specifically designed to analyse photographed images of the test strip, control for variations in camera, lightning, and operating system, check for input or output irregularities, and mathematically derive the associated hormone levels.[111] In case of a failure of the analysis due to loss of internet, then an offline analysis by the company is feasibly by providing the time, day and picture of the test strip of the participant. After the completion of the 30-day urine hormone monitoring and in consideration the first day of the last menstrual bleeding, Cycle Days and the participants' MCP will be defined by visually inspecting the 30-day hormone profile. The follicular phase will be defined from the Cycle Day 1 (first day of menstrual bleeding) until the LH peak; the luteal phase will be defined as the day after the LH peak until the day before the next menstrual bleeding.

Hypoxic-ventilatory response test

The hypoxic ventilatory response at exercise (HVRe) has been suggested to be a good predictor for AMS.[19,112] Especially in premenopausal females, it has been suggested that HVRe is higher in the luteal compared to the follicular phase, providing

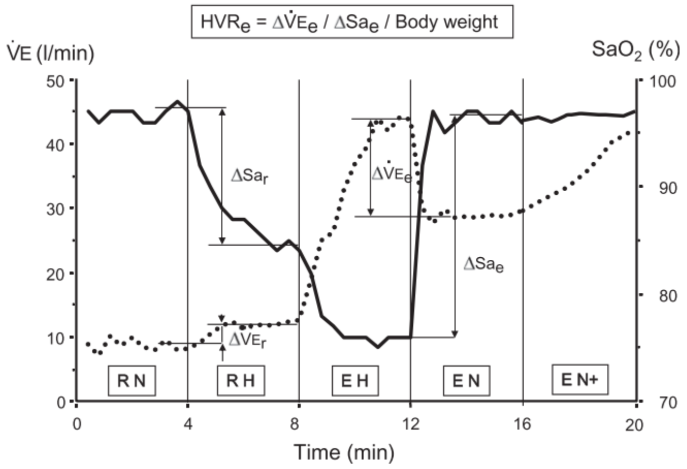

***Figure 4: Hypoxic exercise test.*** *Figure 1 from Richalet et al. 2012.*[104] *RN, RH, EH, and EN represent rest in normoxia, rest in hypoxia, exercise in hypoxia, and exercise in normoxia, respectively. Hypoxia will be produced by breathing normobaric hypoxic gas (fraction of inspired oxygen of 0.115). Exercise will be performed at 30% of maximal exercise capacity in phases EH and EN. In phase EN+, the exercise intensity will be adjusted so that the heart rate reaches the same value as in EH. HVRe, ventilatory response to hypoxia during exercise;* $SaO_2$*, arterial oxygen saturation assessed by finger pulse oximetry;* $Sa_e$*, arterial oxygen saturation during exercise;* $Sa_r$*, arterial oxygen saturation at rest; V'E, minute ventilation;* $V'E_e$*, minute ventilation during exercise. Dotted line represents V'E; solid line represents* $SaO_2$*.*

a potential underlying factor explaining MCP-related differences in AMS susceptibility. Therefore, subjects will perform an exercise test consisting of 5 consecutive phases: 1 – rest in normoxia; 2 – rest in hypoxia (FiO2 of 0.115 equivalent to 4800 m); 3 – exercise in hypoxia (EH) at 30% of maximal normoxic maximal exercise capacity (Ergoselect 200; Ergoline GmbH, Bitz, Germany); 4 – exercise in normoxia (EN) and 5 – EN with the same heart rate achieved during EH (EN+). Arterial oxygenation by finger pulse oximetry and respiratory gas concentrations, tidal volume, and respiratory rate will be measured breath-by-breath by a metabolic unit (Ergostick; Geratherm Medical AG, Gschwenda, Germany) to compute minute ventilation (V'E) and other outcomes. HVRe will be calculated as previously described and illustrated in **figure 4**.

Haemoglobin mass ($Hb_{mass}$) and intravascular volumes

Plasma volume (PV) contraction is an early mechanism allowing an increase in haemoglobin concentration ([Hb]) (and hence, in arterial oxygen content) after an acute high-altitude exposure, and thus, counteracting the decrease in oxygen availability.[113] Experimental studies distinctly conducted in both male and female voluntaries confirmed that this decrease in PV occurred early (*i.e.*, in the first 12-24-hours) and with a similar magnitude between sex, mainly through a fluid-redistribution mechanism from the intra to the extravascular compartment, rather than due to a hypoxic diuresis.[114,115] Beneficial effects of a preventive acetazolamide uptake on AMS are assumed to be mostly driven by the consecutive hyperventilation balancing the induced metabolic acidosis and, thus, increasing the arterial oxygen content; however, even at a such low dose, acetazolamide might have beneficial effect on arterial oxygen content, through its diuretic effect, by further decreasing the PV.[116] To investigate PV changes, an $Hb_{mass}$ measurement using the CO-rebreathing method, allowing the computation of PV, will be conducted at 760 m and after 12 hours staying at 3600 m. Briefly, the CO-rebreathing test will be conducted as previously descripted, using an automated system (OpCo, Detalo Health, Copenhagen, Danmark), after a 20-min rest period in supine position.[117] From a capillary blood sample, [Hb], pre-test percentage of carboxyhaemoglobin (Rapidpoint 500, Siemens AG, Zürich) and haematocrit (Hct, microcentrifuge method) will be determinate in quadruplicate. Next, while the participant will breathe a 100% mixture of oxygen in a rebreathing circuit, a bolus dose of 99.997% pure CO gas (1 mL per kg body mass for men, 0.8 mL per kg body mass for females) will be automatically administered by the Opco system. After a 6-min rebreathing period, the participant will be disconnected from the rebreathing circuit and the residual CO concentration in the circuit will be measured. Then, a secondary capillary blood sampling will be performed at 10 min, to measure, in quadruplicate, the post-test %HbCO ($\%HbCO_{POST}$). From the post-pre-test difference in %HbCO and the amount of absorbed CO, the $Hb_{mass}$ value will be calculated and the derived intravascular volumes (PV, red blood cell volume and total blood volume) will be calculated.[117] Previous studies conducted in the field in high-altitude environment have confirmed both the feasibility and the safety of the CO-rebreathing method.[105,118,119]

Hemorheological analysis

Potential effects of a prophylaxis acetazolamide uptake in order to prevent AMS on blood viscosity could be leaded by an expected decrease in PV or by a direct effect on red blood cell (RBC) deformability due to a direct inhibition of RBC carbonic anhydrase,[113] albeit hemorheological evaluations have never been performed in this context. To explore the potential hemorheological effect of acetazolamide prophylaxis administration, and the potential difference of effect between males and females, we will plan to perform blood viscosity and RBC's deformability measurements at 760 m and after the first night at 3600 m. At both altitudes, from a 3 mL venous sample collected in an EDTA tube, the following analyses will be performed, in accordance with the guidelines for hemorheological laboratory techniques:[120]

- Blood viscosity will be measured at native and corrected Hct (40%, after dilution using autologous plasma) at several shear rates (22.5 $s^{-1}$, 45 $s^{-1}$, 90·$s^{-1}$).
- RBC's deformability will be measured by ektacytometry using the Laser Optical Rotational Red Cell Analyzer (LORRCA, RR Mechatronics, Hoorn, The Netherlands) at 3 and 30 Pa and the extent of RBC aggregation and the strength of RBC aggregates will be measured by syllectometry using the LORRCA (RR Mechatronics, Hoorn, The Netherlands) after adjustment of the Hct to 40% with autologous plasma.

Arterial and venous blood sampling

Effects of altitude and acetazolamide on arterial blood gases will be assessed in a radial artery sample after 15 minutes of quiet and awake supine position in the morning after awakening. As previously described, arterial blood will be analysed for pH, $PaO_2$, $PaCO_2$, $SaO_2$, haematocrit and electrolytes (Rapidpoint 500, Siemens AG, Zürich).[38] To quantify and confirm MCPs, absolute hormone concentrations and plasma concentration of acetazolamide and to provide mechanistic explanations to PV changes, a 20 mL peripheral venous blood sample in the morning after awakening will be withdrawn, centrifuged, and the plasma/serum immediately frozen, stored, and transported at -20°C for further analyses. Analysis at 760 m will include iron metabolism (ferritin, TSAT, hepcidin, erythroferrone, sTfR1). At both altitudes, circulating reproductive hormone concentrations (oestradiol, progesterone, LH) , plasma renin activity, aldosterone, copeptin (as a surrogate of the antidiuretic hormone) and midregional proANP (the precursor of the atrial natriuretic peptide) will be analysed.

Cardiopulmonary exercise testing and electrocardiogram

As previously performed by our project team, a resting 12-lead ECG will be performed after resting for 10 minutes in a supine, awake position which will give insights into cardiac repolarization disturbances at 3600 m.[70] Thereafter, a bicycle exercise test with a progressive ramp protocol will be performed until exhaustion (Ergoselect 200, Ergoline GmbH, Bitz, Germany) as previously performed in patients with COPD and healthy;[40,67] and according to international guidelines to assess alterations in physical exercise performance at altitude and under acetazolamide.[42,121] Participants will rest on the bicycle for 10 minutes, followed by a 10 – 20 W/min work-load increase starting at 20 W. Respiratory gas concentrations, tidal volume, and respiratory rate will be measured breath-by-breath (Ergostick, Geratherm Medical AG, Gschwenda, Germany). Arterial oxygenation (by finger oximetry) and a 12-lead ECG will be applied. Before and at peak exercise, dyspnoea sensation and leg fatigue will be assessed with the BORG CR10 scale.[122]

Cardiorespiratory sleep studies

Continuous nocturnal measurements by portable devices (Alice PDX, Philips Respironics, Zofingen, Switzerland) will include nasal pressure swings, chest wall excursion, ECG, and pulse oximetry. Mean oxygen saturation and percentage time with $SpO_2$ <90% and <80% will be computed. The apnoea/hypopnea index and oxygen desaturation index ($SpO_2$ dips >3%) will be computed as mean number of events/h.[52] These measurements will give valuable insights into sleep-disordered breathing often seen at altitude and the effect of acetazolamide. This measurement has been successfully implemented by our project team in patients with COPD and healthy lowlanders and highlanders.[7,123]

Cerebrovascular reactivity assessment

Participants will undergo transcranial near-infrared spectroscopy (NIRS) measurements of the prefrontal cortex, finger pulse oximetry, transcranial Doppler ultrasound (TCD), electrocardiography (ECG), measurement of continuous blood pressure, end-expiratory $PCO_2$ measurement and breathing pattern while the participant is sitting in a comfortable position for 10 minutes breathing room air. After baseline measurements, the participant will be asked to hyperventilate and reduce their expired $PCO_2$ by 10 mmHg or do an isometric exercise by pressing a predefined load in the dominant hand by a hand grip dynamometer for 1 minute, in random order. The TCD probe will be used to insonate the middle cerebral artery on the same side as the hand grip maneuver is performed to eliminate neurovascular activity contamination. Between interventions a 5-minute wash-out phase will be implemented to avoid any carry-over effect. The last 2 minutes of quiet breathing room air will be used as baseline measurement and will be compared with hyperventilation in participants under placebo compared to participants under acetazolamide. Furthermore, blood flow response in the middle cerebral artery due to blood pressure alterations will be compared between altitudes, drug and sex.

To complement TCD's assessment of regional cerebral blood flow (CBF) velocity through the middle cerebral artery, global CBF will be assessed during the same procedures using Duplex ultrasound of both the right internal carotid and the right vertebral arteries (ICA and VA, respectively). Combined diameter and blood velocity measurements provide the necessary data to calculate blood flow through these vessels. Owing the ICA and VA are bilateral, the calculated blood flow will then be multiplied by a factor of 2 to obtain a value for total CBF.

Biobeat Wrist Monitor

The non-invasive biobeat Wrist Monitor continuously monitors vital parameters including heart rate, breathing frequency, arterial oxygenation and systolic and diastolic blood pressure. The patented algorithm is based on artificial intelligence and machine-learning and has been used in other scientific settings.[124] A unique feature is that the vital parameters are not displayed on the Wrist Monitor, instead, are uploaded to the cloud and are live accessible by the physician through a remote patient monitoring system. This approach minimizes confounding the participant and allows high-quality remote monitoring of the participants. Moreover, vital parameters will be continuously recorded and might allow in the future to use an early warning system to prevent AMS. During this study, each participant will wear a biobeat Wrist Monitor during the complete duration of the altitude sojourn. The continuous data recording will be modelled and analysed by partners of the ETH Zurich.

Lung ultrasonography

Using a phased array probe, bilateral lung ultrasound will be performed in a supine from the second to fourth or fifth intercostal space (left and right hemithorax, respectively) down the parasternal, mid-clavicular, anterior axillary and mid-axillary lines, resulting in 28 total windows of interest (left: 12; right: 16). A B line will be defined as one echogenic, continuous, wedge-shaped, signal arising from the uninterrupted pleural interface with a narrow origin in the near field of the image. The number of B-lines will be counted for each lung field and totalled.[76]

Echocardiography

Cardiac morphological and functional parameters will be assessed by standard two-dimensional Doppler echocardiography. Doppler imaging of the tricuspid annulus and the pulmonary outflow tract will be assessed according to standard methods. This will allow us to assess the following parameters: systolic pulmonary artery pressure ($RV_{sys}$, calculated from the tricuspid regurgitation velocity by using the modified Bernoulli equation: $RV_{sys} = 4x\ (v_{max})$ where $v_{max}$ is the maximum of the regurgitation velocity jet measured over the tricuspid valve) and the right ventricular outflow tract acceleration time; the right ventricular fractional area change [%] and the tricuspid annular plain systolic excursion [mm]. The fractional area change of the right ventricle will be determined. The tricuspid annular plane systolic excursion (TAPSE) will be measured in M-mode. The stroke volume will be measured based on the left ventricular outflow tract diameter and the velocity time integral over the aortic valve in the apical 5- chamber view or the apical long axis view.

##### 2.3.7. Outcomes

The primary outcome of this study is the absolute difference in efficacy of acetazolamide to reduce AMS incidence in females compared to men. AMS will be defined as a 2018 Lake Louise questionnaire score of ≥3 points including headache.[15] If a participant scores ≥3 points while staying at 3600 m, the participant will be deemed to have AMS.

Secondary outcomes will be derived from urinary sexual hormone concentrations, AMS severity, AMS symptom components, and AMS defined by different questionnaires and cut-off scores.[109,110] Vital parameters, parameters from arterial and venous blood sampling, hypoxic ventilatory response during exercise, cerebrovascular reactivity and respiratory sleep studies will be obtained and compared between sexes, between acetazolamide and placebo and between MCPs. Additionally, ECG morphology, cardiac repolarization disturbances, sleep-disordered breathing and exercise performance and exercise-limiting factors will be obtained and compared. Mechanistic insights of the prophylactic effect of acetazolamide for AMS will be derived from intravascular volume measurements, $Hb_{mass}$, blood viscosity and hemorheological assessments.

##### 2.3.8. Randomisation, intervention and blinding

Stratified randomization will be conducted for females and males using a computer generated schedule (MinimPy 0.3), stratifying for sex in a females-to-males ratio of 2:1.[125] Within sex, participants will be 1:1 randomized to A: 250 mg/day acetazolamide (125 mg in the morning with breakfast, 125 mg in the evening with dinner) starting 24 hours before arriving and while staying for 2 days and nights at 3600 m (total of 6 capsules of 125 mg acetazolamide) or B: identically looking, organoleptic placebo capsules. An independent pharmacist will prepare identically looking verum and placebo capsules labelled with secret codes. The list of codes will be kept confidential to investigators and participants until data acquisition and analysis has been completed.

##### 2.3.9. Sample Size Estimation

Sample size estimation was performed using the Cochran Mantel-Haenszel test with continuity correction. The previously described preventive effect of acetazolamide against AMS has been suggested to be 44%.[34] To detect a clinically meaningful efficacy difference of 20% of acetazolamide therapy against AMS in females compared to males (i.e. treatment effect of 50% in females and 30% in men) with a group allocation ratio of 2:1, power of 90%, two-sided alpha level of 0.05 and accounting for dropouts, a total of 180 females and 90 males will be recruited. The females-to-males ratio of 2:1 was chosen to enable conclusive testing of the additional hypotheses within females (see **section 2.3.3**). The project team has successfully recruited even higher numbers of COPD patients and healthy participants in previous randomized clinical trials at high altitude. As an example, in 2015, 124 COPD patients were recruited within 1 summer expedition and included in the final analysis[36]. In another trial of acetazolamide, 186 COPD patients and 345 healthy subjects were recruited and included in the final analysis within 3 summer expeditions.[7] Since the complex recruitment of these large numbers of chronically ill patients was successful, the anticipated number of healthy females and males required for this study is well feasible and within the capacity of the project team.

##### 2.3.10. Data Analysis and Statistics

In accordance with international guidelines for prospective clinical trials,[126] a statistical analysis plan, similarly to the studies published by the applicant in *NEJM Evidence,*[7] will be created before any statistical analyses. This study will be pre-registered at ClinicalTrials.gov and results reporting will follow the CONsolidated Standards Of Reporting Trials (CONSORT) guidelines.[127] Data will be summarised by numbers and proportions and means (SD) and medians (quartiles) for normally and non-normally distributed data, respectively. Altitude-induced differences will be calculated by mean or median differences (95% confidence intervals). The primary analysis will be performed on the intention-to-treat population including all randomized participants using the Cochran Mantel-Haenszel test comparing the sex-related difference in the acetazolamide effect against AMS.[128] In case of missing values in the primary outcome (AMS, binary), a sensitivity analysis with best / worst case scenario will be applied. Kaplan-Meier curves for AMS will be plotted for the four groups representing the combination of sex and treatment assignment: 1) males randomly assigned to receive acetazolamide; 2) males randomly assigned to receive placebo; 3) females randomly assigned to acetazolamide; 4) females randomly assigned to placebo. Comparisons will be performed using a log-rank test. Secondary, a multivariable Cox proportional-hazards analysis will be conducted to determine whether the interaction between sex and acetazolamide therapy is independent of other baseline factors.

Secondary outcomes will be analysed on the per-protocol population defined as participants with available data. Continuous variables will be analysed using mixed linear regression models, with the variable of interest as the dependent variable and drug, location, sex and MCP as fixed effects, including the interaction term drug*location*sex*MCP.

To proactively minimize any unforeseen deviations from the anticipated study progression, such as extreme benefit or harm of acetazolamide or futility and unexpected AMS incidences, a planned interim analysis will be conducted after the completion of the first study year. The planned interim analysis will be performed by an independent statistician blinded to drug. The statistician will report the blinded results to the project steering committee (composed by the applicant and the co-applicants), who will decide on continuation, termination, or on any appropriate protocol adaptation (e.g., sample size re-estimation). Termination of the study is defined by symmetric stopping boundaries at $p<0.001$ (Peto approach).[129] In the final analysis, a $p<0.05$ will be considered statistically significant.

##### 2.3.11. Further considerations

As in previous studies over the past few years, the applicant has access to a dedicated and experienced Swiss and Kyrgyz research team composed of assistant physicians and medical students, as well as access to pivotal medical devices needed for the described measurements such as ergometers, respiratory polygraphy, computers, and other hardware. This equipment is available from the *Swiss-Kyrgyz High Altitude Medicine and Research Initiative*. Medical devices and consumables will be transported to Kyrgyzstan for the study duration. During data acquisition periods, up to 6 medical bachelors' students from ETH Zürich will perform their 6-week research internship in Kyrgyzstan. Prof. Silvia Ulrich will dedicate additional Swiss staff for preparing and conducting this project during the years 2024 and 2025. The Kyrgyz team led by Prof. Sooronbaev (The Kyrgyz director of the Swiss-Kyrgyz High Altitude Research Center and Director of the University Hospital in Bishkek) will contribute the required personnel to recruit participants and for data acquisition, facility maintenances at low and high altitude, and transportation.

In regard of the health and safety of the participants at 3600 m, the Kumtor Gold mine Facility owns a fully equipped medical station including supplemental oxygen, emergency medications, medical staff and ambulances to quickly evacuate to lower altitudes.

#### 2.4. SCHEDULE AND MILESTONES

The schedule and milestones of the project are outlined in **Figure 5**. The project partners will meet several times per year to decide on scientific, logistic and administrative aspects and other questions that might arise. The study preparations will be started on January 2024 and will recruit 140 participants in the summer season 2024. Milestone 1.1 (M1.1) represents the successful completion of the first expedition. In 2025, the second expedition with another 140 participants will take place (M1.2). Data analyses will be conducted in the following months, and manuscript submission can be expected thereafter (M1.3). In 2026, analysis of secondary outcomes such as pooled blood samples, exercise, cerebrovascular and ventilatory responses will be scheduled (M1.4).

| 01.01.2024 to 31.12.2026 (3 years) | 2024 | | | | | | | | | | | | 2025 | | | | | | | | | | | | 2026 | | | | | | | | | | | |
|---|---|---|---|---|---|---|---|---|---|---|---|---|---|---|---|---|---|---|---|---|---|---|---|---|---|---|---|---|---|---|---|---|---|---|---|---|
| | J | F | M | A | M | J | J | A | S | O | N | D | J | F | M | A | M | J | J | A | S | O | N | D | J | F | M | A | M | J | J | A | S | O | N | D |
| Preparation | | | | | | | | | | | | | | | | | | | | | | | | | | | | | | | | | | | | |
| Data Collection | | | | n = 140 | | | | | | M1.1 | | | | | | n = 140 | | | | | | M1.2 | | | | | | | | | | | | | | |
| Interim Analysis | | | | | | | | | | | | | | | | | | | | | | | | | | | | | | | | | | | | |
| Primary data analysis | | | | | | | | | | | | | | | | | | | | | | | | | | | | M1.3 | | | | | | | | |
| Secondary data analysis | | | | | | | | | | | | | | | | | | | | | | | | | | | | | | | | | | | M1.4 | |
| | J | F | M | A | M | J | J | A | S | O | N | D | J | F | M | A | M | J | J | A | S | O | N | D | J | F | M | A | M | J | J | A | S | O | N | D |
| | 2024 | | | | | | | | | | | | 2025 | | | | | | | | | | | | 2026 | | | | | | | | | | | |

***Figure 5. Schedule and Milestones of the proposed study.*** *Grey areas indicate periods of scheduled work packages, Letters represent months of the years 2024-2026.*

#### 2.5. RELEVANCE AND IMPACT

##### 2.5.1. SCIENTIFIC RELEVANCE

Previous research has been conducted to elucidate AMS susceptibility, prevention, and treatments, but females have only occasionally been included and have never been appropriately characterized. No randomized clinical trial has been conducted to investigate and compare the efficacy of acetazolamide against AMS incidence in females compared to men, despite the knowledge that females are more prone to develop AMS compared to men. Differences in drug efficacy or side-effects based on sex-related differences are indicated in the literature and are likely due to lower blood volume and consequently higher acetazolamide plasma concentration, due to differences in the hypoxic chemosensitivity or other factors between females and men. The current gaps in our knowledge are mainly due to the demanding logistical and organisational challenges in conducting large-scale mountain medicine and physiology research, especially in females. The applicant and his partners have acquired the knowledge and study setting to overcome these challenges. Therefore, this study will provide conclusive results on the efficacy and side-effect of preventive acetazolamide against AMS in females compared to males and whether AMS susceptibility depends on female sex hormones or MCPs. Moreover, physiological, urine and blood analyses will provide comprehensive insights into physiological and clinical differences between females and males under hypoxic conditions. Findings from this large-scale project are expected to become milestones in the research history of high-altitude medicine, opening up new avenues for mechanistic research related to sex- and hormone-dependent tolerance to hypoxia.

##### 2.5.2. Broader Impact

Females are substantially underrepresented in heart failure, acute coronary syndrome and pulmonary disease studies, with females <55 years accounting for <10% of the study population and in case of an incident females present with a worse outcome.[30,130,131] The literature shows that many lung diseases are more commonly found in females and present with higher degree of severity, exacerbation rate, hospitalizations and mortality than in men.[132] In this regard, insights from this altitude study investigating hypoxia- and hypoxemia-related physiological and clinical adaptations in females compared to males might enhance our understanding of the known sex-related differences in lung diseases at low altitude. Furthermore, to date, there is limited performance of hormonal and cell immunological diagnostic endeavour and low inclusion of female participants in clinical trials on AMS prevention. Our findings are expected to have a large and broad impact, since a considerable number of summer and estimated 400 million annual winter visits will eventually be affected by altitude-related health impairments.[5,133] Based on the expected results, female athletes might be able to improve their high altitude performance or live-high, train-low training schedules based on the MCP. Exercise training based on the MCP is already common in top athletes, therefore, sports in the mountains are equally affected. Understanding sex differences in altitude tolerance may finally result in less accidents and emergency evacuations, as well as improved work efficiency in employees temporarily working at high altitudes. Moreover, translational knowledge transfer of these findings from hypoxemic conditions might contribute to a better understanding of acute cardiopulmonary diseases associated with hypoxemia (i.e. COVID), their progression, their therapy, and any related sex differences as well as to millions of highlanders living at altitudes >2500 m, who are chronically hypoxemic.

10.1378/chest.14-0495

112. Richalet JP, Larmignat P, Poitrine E, Letournel M, Canoui-Poitrine F. Physiological risk factors for severe high-altitude illness: a prospective cohort study. *Am J Respir Crit Care Med*. Jan 15 2012;185(2):192-8. doi:10.1164/rccm.201108-1396OC
113. Siebenmann C, Robach P, Lundby C. Regulation of blood volume in lowlanders exposed to high altitude. *J Appl Physiol (1985)*. Oct 1 2017;123(4):957-966. doi:10.1152/japplphysiol.00118.2017
114. Schlittler M, Gatterer H, Turner R, et al. Regulation of plasma volume in male lowlanders during 4 days of exposure to hypobaric hypoxia equivalent to 3500 m altitude. *J Physiol*. Feb 2021;599(4):1083-1096. doi:10.1113/jp280601
115. Roche J, Rasmussen P, Gatterer H, et al. Hypoxia briefly increases diuresis but reduces plasma volume by fluid redistribution in women. *Am J Physiol Heart Circ Physiol*. Dec 1 2022;323(6):H1068-h1079. doi:10.1152/ajpheart.00394.2022
116. Swenson ER. Pharmacology of acute mountain sickness: old drugs and newer thinking. *Journal of Applied Physiology*. 2016;120(2):204-215. doi:10.1152/japplphysiol.00443.2015
117. Siebenmann C, Keiser S, Robach P, Lundby C. CORP: The assessment of total hemoglobin mass by carbon monoxide rebreathing. *J Appl Physiol (1985)*. Sep 1 2017;123(3):645-654. doi:10.1152/japplphysiol.00185.2017
118. Champigneulle B, Stauffer E, Robach P, et al. Early effects of acetazolamide on hemoglobin mass and plasma volume in chronic mountain sickness at 5100 m. *Pulmonology*. May 30 2023;doi:10.1016/j.pulmoe.2023.05.006
119. Stembridge M, Williams AM, Gasho C, et al. The overlooked significance of plasma volume for successful adaptation to high altitude in Sherpa and Andean natives. *Proc Natl Acad Sci U S A*. Aug 13 2019;116(33):16177-16179. doi:10.1073/pnas.1909002116
120. Baskurt OK, Boynard M, Cokelet GC, et al. New guidelines for hemorheological laboratory techniques. *Clinical hemorheology and microcirculation*. 2009;42(2):75-97. doi:10.3233/ch-2009-1202
121. Ross RM. ATS/ACCP statement on cardiopulmonary exercise testing. *Am J Respir Crit Care Med*. May 15 2003;167(10):1451; author reply 1451. doi:10.1164/ajrccm.167.10.950
122. Borg G. *Borg's perceived exertion and pain scales*. Human kinetics; 1998.
123. Sooronbaev T, Osmonov B, Furian M, et al. Time course of pulmonary artery pressure and clinical characteristics in Kyrgyz highlanders. *Eur Respir J*. 2015;46(suppl 59):PA3761. doi:10.1183/13993003.congress-2015.PA3761 %J European Respiratory Journal
124. Nachman D, Gilan A, Goldstein N, et al. Twenty-Four-Hour Ambulatory Blood Pressure Measurement Using a Novel Noninvasive, Cuffless, Wireless Device. *Am J Hypertens*. Nov 20 2021;34(11):1171-1180. doi:10.1093/ajh/hpab095
125. Saghaei M. An overview of randomization and minimization programs for randomized clinical trials. *J Med Signals Sens*. Jan 2011;1(1):55-61.
126. Gamble C, Krishan A, Stocken D, et al. Guidelines for the Content of Statistical Analysis Plans in Clinical Trials. *JAMA*. 2017;318(23):2337-2343. doi:10.1001/jama.2017.18556
127. Schulz KF, Altman DG, Moher D, Group C. CONSORT 2010 statement: updated guidelines for reporting parallel group randomised trials. *BMJ*. Mar 23 2010;340:c332. doi:10.1136/bmj.c332
128. Rathore SS, Wang Y, Krumholz HM. Sex-Based Differences in the Effect of Digoxin for the Treatment of Heart Failure. *New England Journal of Medicine*. 2002;347(18):1403-1411. doi:10.1056/NEJMoa021266
129. Pocock SJ. When to stop a clinical trial. *Bmj*. Jul 25 1992;305(6847):235-40. doi:10.1136/bmj.305.6847.235
130. Pilote L, Raparelli V. Participation of Women in Clinical Trials: Not Yet Time to Rest on Our Laurels. *J Am Coll Cardiol*. May 8 2018;71(18):1970-1972. doi:10.1016/j.jacc.2018.02.069
131. Arora S, Stouffer GA, Kucharska-Newton AM, et al. Twenty Year Trends and Sex Differences in Young Adults Hospitalized With Acute Myocardial Infarction. *Circulation*. Feb 19 2019;139(8):1047-1056. doi:10.1161/CIRCULATIONAHA.118.037137
132. Silveyra P, Fuentes N, Rodriguez Bauza DE. Sex and Gender Differences in Lung Disease. *Adv Exp Med Biol*. 2021;1304:227-258. doi:10.1007/978-3-030-68748-9_14
133. Vanat L. *2019 International Report on Snow & Mountain Tourism. Overview of the key industry figures for ski resorts.* Vol. 11. 2019. April 2019. Accessed 12.11.2020. https://www.vanat.ch/RM-world-report-2019.pdf

### Sex-specific efficacy and safety of preventive acetazolamide for acute mountain sickness in healthy lowlanders: a randomised, double-blind, placebo-controlled trial.

**Principal Investigator**

- Dr. sc. ETH Michael Furian, University Hospital Zurich, Pulmonology Department, Zurich, Switzerland

**Swiss Project Partners**

- Prof. Dr. med. Silvia Ulrich, MD, Dept. of Respiratory Medicine, University Hospital Zurich, Zurich, Switzerland
- Prof. Dr. med. Konrad Bloch, MD, Dept. of Respiratory Medicine, University Hospital Zurich, Zurich, Switzerland
- PD Dr. med. Cornelia Betschart, MD, Gynaecology Department, University Hospital Zurich, Switzerland

**International Project Partners**

- Prof. Dr. med. Talant Sooronbaev, MD, Dept. of Respiratory Medicine, National Center of Cardiology and Internal Medicine, Bishkek, Kyrgyzstan
- Dr. sc. hum, MSc. med. Biometry & Statistics Nicola Benjamin, Center for pulmonary hypertension, University Clinic Heidelberg, Germany
- Research Consortium of EXALT – Centre d'Expertise sur l'Altitude, France:
  Prof. Dr. sc. Samuel Vergès (Université de Grenoble), Prof. Dr. Julien Brugniaux (Université de Grenoble), Dr. Benoit Champigneulle (Université de Grenoble), Prof. Dr. Aurélien Pichon (Université de Poitiers), Prof. Dr. Paul Robach (Ecole Nationale de Ski et d'Alpinisme), Dr. Emeric Stauffer (Hospices Civils de Lyon)

**Contact**

Dr. sc. ETH Michael Furian

University Hospital Zurich

Pulmonology Department

Raemistrasse 100

CH-8091 Zurich, Switzerland

10.06.2024, Version 1.1

### 1. Summary of the research plan

**Background and rationale:** Millions of people travel to high altitude for work or leisure activities and are exposed to reduced inspiratory oxygen partial pressure and hypoxemia that may lead to altitude illness, among which the most common form is acute mountain sickness (AMS). The main AMS symptoms are headache, malaise, weakness, and fatigue. Prospective studies have shown that 20–60% of newcomers at 2500–4000m develop AMS requiring them to take medications, while, at very high altitudes, AMS may progress to high altitude cerebral oedema. Whether females are more susceptible to AMS remains insufficiently understood since no prospective study controlled for sex hormones, use of hormone contraception or assessed menstrual cycle phase (MCP) at altitude. Therefore, females remain underrepresented and poorly characterized in high altitude studies. In addition, the efficacy and safety of 250 mg/day acetazolamide, the standard recommendation for AMS prevention, has never been compared between sexes, although, females have presumably higher acetazolamide plasma concentration due to lower blood volume. Given the known dose-dependent preventive but also side effects of acetazolamide and equal proportion of females and males among mountain travellers, there is an urgent need to conclusively quantify the efficacy and safety of preventive acetazolamide therapy against AMS in females compared to men.

**Overall objectives and specific aims:** The primary objective is to compare the efficacy of preventive acetazolamide treatment against AMS in females compared to men. Additional objectives are to investigate sex- and MCP-related differences in altitude tolerance including AMS incidence as well as efficacy and safety of acetazolamide compared to placebo.

**Methods:** In this randomized, placebo-controlled, double-blinded parallel trial, healthy, premenopausal females and males aged 18 to 44 years, living <1000m will be recruited. Baseline measurements will take place at the Bishkek University Hospital (760m), Kyrgyz Republic; altitude measurements after ascending within 6h by car to 3600m and during a subsequent stay for 2 days and nights. The primary outcome will be the absolute difference in the acetazolamide treatment effect to reduce AMS incidence in females compared to males during the 2-day stay at 3600m. AMS will be defined as a Lake Louise questionnaire score of ≥3 including headache. Secondary outcomes will be derived from respiratory sleep studies, exercise testing, cerebrovascular reactivity assessments and urine and blood drawings.

Starting the day before ascent and daily in the following 30 days, females will perform self-measurements of morning urine concentrations of estrone-1-glucuronide, pregnanediol-3-alpha-glucuronide and luteinizing hormone. From these, the Cycle Days and MCP of participants while at 3600m will be derived. The primary analysis will be on the intention-to-treat population using the Cochran Mantel-Haenszel test. Secondary outcomes and the influence of MCP will be analysed by mixed linear or logistical regression models. To detect a clinically relevant absolute 20% efficacy difference of acetazolamide between sexes, using a ratio of 2:1, a power of 90%, two-sided alpha level of 0.05 and accounting for dropouts, a total of 180 females and 90 males are required.

**Expected results:** This project will conclusively assess the efficacy and safety of preventive acetazolamide in premenopausal females compared to males at a representative altitude for many touristic destinations worldwide. Additionally, this study will provide unique insights into clinical and physiological sex- and MCP-related differences in altitude tolerance.

**Impact for the field**: The detailed characterization of females traveling to altitude and using acetazolamide will represent a milestone in high altitude medicine and physiology and will substantially contribute to our understanding of sex-related altitude tolerance and the effect of acetazolamide. Insights about MCP-dependent susceptibility to hypoxia and AMS symptoms will translate to our physiological and mechanistical understanding of the human body.

### 2. Research Plan

#### 2.1. Current State of Research in the Field

On September 27th 2022, the Swiss National Council promoted research and therapy of specific females' diseases.[1] Consequently, the Swiss Confederation has just now commissioned the SNSF to establish four new National Research Programmes, among them "gender medicine".[2] There are numerous diseases that affect females exclusively or in the majority, however, the Committee for Social Security and Health of the National Council writes that research into treatment options is lagging. It is therefore essential that females-specific diseases are identified as such and researched more broadly, like through research programs of the Swiss National Science Foundation (SNF). In addition, guidelines for diagnosis, indication and therapy are needed, as well as a clear definition for quality outcome measurements in females. Too little is known about females' diseases, they remain undiagnosed for a long time and cause unnecessary suffering. As indicated in the literature, acute mountain sickness (AMS) might be one of the diseases affecting females more than men.

##### 2.1.1. Acute altitude-related illnesses in healthy individuals

Mountain tourism accounts for 15% to 20% of the annual global tourism revenue (approximately 296 billion US dollars in 2019),[3] highlighting the popularity of trips to mountainous regions. However, mountain travel exposes the human body to lower barometric pressures and reduced arterial blood oxygenation (hypoxaemia), which requires numerous physiological adaptations to protect the body and organs against hypoxaemia-related dysfunction and damage. However, moderate hypoxaemia can trigger the development of acute mountain sickness (AMS)[4] and other conditions that compromise a stay at altitude, e.g., poor sleep quality and exercise intolerance. AMS is the most important acute altitude-related illness, affecting 20-60% of unacclimatized lowlanders staying overnight at an altitude between 2500 and 4000 m.[5] The main symptoms of AMS that may emerge within 6 hours to 1-2 days after ascent to high altitudes are headache accompanied by malaise, weakness, and fatigue, which often resolves after 48 hours at altitude.[4] However, AMS can force people to take medications or prematurely terminate their stay at altitude. At altitudes >4000 m, AMS might become life-threatening by progressing to high altitude cerebral oedema.[4] The growing number of publications including in the highest-ranked medical journals (i.e. NEJM), emphasize the importance of understanding AMS and other medical conditions at high altitude.[6-8] Despite the growing work, the pathophysiology of AMS remains elusive and consequently, the diagnosis still relies on a pattern of subjective symptoms rather than on objective findings.[9] AMS may develop due to inadequate physiological responses to hypoxaemia such as decreased hypoxic ventilatory response,[10,11] increased sympathoadrenal activity, and insufficient early hypoxic diuresis.[12,13] A history of migraine, a rapid ascent, and previous episodes of AMS are the most robust predictors of AMS. In 2018, analysis of the AMS literature and based on the early onset of AMS symptoms, an international consensus committee composed of 83 international mountain medicine experts concluded that disturbed sleep might be a direct consequence of hypoxia rather than a symptom of AMS, causing a change in the Lake Louise score (LLS),[14] the most common questionnaire used to diagnose AMS. The revised questionnaire now uses the four items, "headache", "gastrointestinal symptoms", "fatigue/weakness", and "dizziness/light-headedness" to diagnose AMS, each rated with a score from 0 (none) to 3 (severe). In the presence of headache, a total score of ≥3 is required to diagnose AMS.

##### 2.1.2. AMS in females

Whether the prevalence of AMS differs between females and males is still being debated and since the modified definition of AMS in 2018, unknown.[15] However, according to the original AMS definition by LLS, a 2019 meta-analysis of 7669 subjects (2639 females) concluded that females have a 1.24-fold risk (95% CI 1.09 to 1.41) of developing AMS at a minimum altitude of 2500 m compared with men.[16] This finding seems logical, since it is known that females do suffer already at low altitude from more headache episodes than men,[17] however, the female physiology under hypoxia remains not at all understood. Important to note, the conclusion of the meta-analysis is debatable since none of the included studies of this meta-analysis assessed the influence of sex hormones (oestrogen, luteinizing hormone, progesterone), menstrual cycle phases (MCP), pre/postmenopausal status or contraceptive use. However, some uncontrolled studies provide important physiological assumptions for sex-related differences in the AMS incidence. Simplified, the sex-related difference in AMS is assumed to be related to sexual hormones and their impact on the physiological response to hypoxia.[16] In accordance with this assumption, progesterone, which is upregulated during the luteal phase, is a well-known respiratory stimulant associated with elevated minute ventilation and decreased $PaCO_2$ during the luteal compared with the follicular phase.[18] Recently, a large study of 336 premenopausal females confirmed these findings and reported lower arterial oxygen saturation ($SaO_2$) and lower hypoxic ventilatory responses in the follicular compared with the early/mid-luteal phase during normobaric hypoxia exposure ($FiO_2$ 0.115, equivalent to 4800 m).[19] These findings are intriguing, since lower $SaO_2$ and lower hypoxic ventilatory response under exercise are risk factors for AMS.[4,19] Another sex-related risk factor for AMS may be low iron stores[20] in females during the follicular phase just after the menstrual bleeding compared to the luteal phase and compared to men.[21,22] Again, no prospective study has investigated the sex-related difference in iron stores on AMS.

##### 2.1.3. Prevention of AMS

Apart from moderate ascent rate and low sleeping altitude, current guidelines recommend acetazolamide, or dexamethasone starting 24 hours before travel to altitude as preventative AMS measures.[23] However, dexamethasone, a glucocorticoid, is mainly recommended for high-risk situations (emergency rescues, military missions) due to its numerous side-effects including hyperglycaemia, immune suppression, and altered mood. Acetazolamide, a carbonic anhydrase inhibitor, is the main recommended and scientifically-proven, pharmacological prophylaxis against AMS.[24] The current recommended dose of 250 mg/day reduces the relative risk of developing AMS by 44%, with a number needed to prevent one case of AMS of 7.[23,24] However, side effects of acetazolamide have been shown to be dose-dependent and include paraesthesia, dysgeusia, polyuria and fatigue.[25] A systematic literature search in PubMed from database inception to September 2023 identified 8 randomized, placebo-controlled, double-blind trials of acetazolamide for prevention of AMS (defined as the primary outcome) using the Lake Louise questionnaire. The studies are summarized in **table 1**. From the 8 identified clinical trials,[7,26-32] only 6 included females.[7,26,27,29,31,32] From the 6 studies including females, only our own trial reported sex-dependent acetazolamide effects on AMS.[7] However, our trial used 375 mg/day instead of 250 mg/Day acetazolamide; was conducted in healthy aged older than 40years and contains a post-hoc analysis. So, in conclusion, none of the only 8 identified clinical trials aimed to investigate sex-differences in the AMS prevention; appropriately characterized the female population, nor used the

2018 version of the LLS to diagnose AMS. Based on the only available trial from our research group in 349 healthy lowlanders, we observed a clear tendency towards a stronger acetazolamide effect in females compared to males (relative risk reduction of 31% in females compared to 21% in men, **figure 1)**.

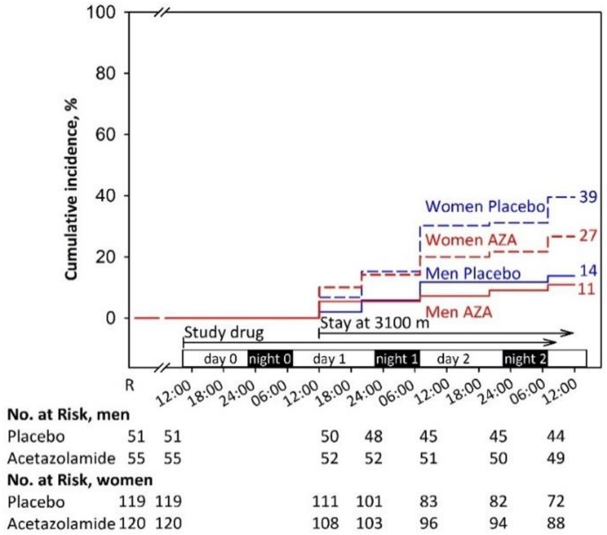

***Figure 1: Incidence of acute mountain sickness and the effect of preventive 375 mg/day acetazolamide therapy starting 24 hours before and during a 2-day stay at 3100 m in healthy females and males older than 40 years (Furian et al., 2022, NEJM Evidence).*** *This post-hoc analysis showed that females were more prone to acute mountain sickness than men; however, no information regarding menopause, menstrual cycle phases, contraceptive use, or hormone supplementation were assessed. Our study also indicated that preventive acetazolamide therapy before ascending to 3100 m might be of relevance for females, but less for men.*

Whether the acetazolamide efficacy against AMS is actually sex-dependent has not been studied and represent an important gap of knowledge in the field of altitude medicine and physiology. Physiologically, there are arguments supporting such differences in the efficacy, so, lower blood volume in females compared to males is likely to contribute towards higher acetazolamide plasma concentration when prescribing a standard dose of acetazolamide.[33] Higher plasma concentrations are likely causing a dose-dependent preventive acetazolamide effect on AMS but might also induce more side effects.[25,34] The physiological pathway might be that the higher acetazolamide plasma concentration in females versus males promote a stronger metabolic acidosis resulting in a more pronounced hyperventilation, consequently, better mitigating altitude-induced hypoxia – the main driving factor for AMS. Another sex-related factor influencing the acetazolamide efficacy may be related to the increased hypoxic chemosensitivity in males compared to females. Thus, males are more prone to high altitude periodic breathing,[35] whereas this type of sleep-disordered breathing has been shown to cause superficial sleep and worse subjective sleep quality. Acetazolamide has been shown to be highly effective in preventing altitude-induced sleep-disordered breathing and to improve subjective sleep quality – especially in men.[35] So, males were presumably more likely to rate their sleep as "improved" under acetazolamide when using the original Lake Louise questionnaire including the question related to "impaired sleep". Consequently, the elimination of the item "impaired sleep" in 2018 is likely reducing the preventive acetazolamide effect against AMS in males whereas in females this change in AMS definition has no impact.

**Table 1. Systematic literature analysis of randomized, placebo-controlled, double-blind trials with primary outcome AMS using the Lake Louise questionnaire (all trials cited in PubMed from database inception to September 2023 are included)**

| Item | Design and Setting | Participants | Main findings | Limitations of the study |
|---|---|---|---|---|
| **Females included, analysed separately for acetazolamide effect** | | | | |
| Furian et al.[7] 2022 | Placebo-controlled RCT of 375 mg/day AZA on AMS (LLscore ≥3 including headache).<br>LLS at baseline at 760 m; Altitude at 3100 m during 48 hours. | 349 (69% females) healthy lowlanders aged 40 years or more, residing <800 m. | Females<br>39.0% AMS with placebo; 27.0% AMS with AZA; RR of 30.8%, NNT of 8.3.<br>Men<br>14.0% AMS with placebo; 11.0% AMS with AZA RR of 21.4%, NNT of 33. | • **Females not characterized[1]**<br>• **375 mg/day instead of 250 mg/day of AZA**<br>• **1993 LLS version for AMS definition[2]**<br>• **Post-hoc analysis of sex-dependent AZA effect in females ≥40 years.** |
| **Females included, not analysed separately or underpowered for detecting sex-related differences** | | | | |
| Lipman et al.[26] 2018 | Placebo-controlled RCT of 250 mg/day AZA or budesonide on AMS (LLscore ≥3 including headache).<br>LLS at baseline at 1240 m: Altitude at 3810 m during 24 hours. | 70 (47.1% females) healthy adult lowlanders residing <1240 m. | 63.0% AMS with placebo; 43.0% AMS with AZA; RR of 32%, NNT of 5. | • **No sex-dependent AZA effect reported**<br>• **Females not characterized[1]**<br>• **1993 LLS version for AMS definition[2]**<br>• **AZA prevention started on day of ascent**<br>• **Assessment <48 hours at target altitude** |
| Van Patot et al.[27] 2008 | Placebo-controlled RCT of 250 mg/day AZA on AMS (LLscore ≥3 including headache).<br>LLS at baseline at 2000 m; Altitude at 4300 m during 24 hours. | 44 (47.7% females) subject residing <1600 m | 77.3% AMS with placebo; 31.8% AMS with AZA; RR of 58.9%, NNT of 2.2 | • **No sex-dependent AZA effect reported**<br>• **Females not characterized1**<br>• **No sex-dependent AZA effect reported**<br>• **1993 LLS version for AMS definition[2]**<br>• **Assessment <48 hours at target altitude** |
| Basnyat et al.[29] 2003 | Placebo-controlled RCT of 250 mg/day AZA on AMS (LLS headache + 1 other symptom).<br>LLS at baseline at 4243 m; Altitude at 4937 m. | 155 (32.9% females) trekkers. | 24.7% AMS with placebo; 12.2% AMS with AZA; RR of 50.6%, NNT of 8 | • **No sex-dependent AZA effect reported**<br>• **Females not characterized[1]**<br>• **No baseline <1500 m**<br>• **Trekkers already acclimatized**<br>• **No sex-dependent AZA effect reported**<br>• **Modified AMS definition[2]**<br>• **Assessment <48 hours at target altitude** |
| Chow et al.[31] 2005 | Placebo-controlled RCT of 500 mg/day AZA or Ginkgo biloba on AMS (LLscore ≥3 including headache; also in combination with the clinical assessment score).<br>LLS at baseline at 1230 m; Altitude at 3800 m during 24 hours. | 47 (44.7% females) adult lowlanders residing <1200 m. | 60.0% AMS with placebo; 30.0% AMS with AZA; RR of 50%, NNT of 3.3<br>1 female under placebo developed HAPE. | • **No sex-dependent AZA effect reported**<br>• **Females not characterized[1]**<br>• **No sex-dependent AZA effect reported**<br>• **Modified AMS definition[2]**<br>• **Assessment <48 hours at target altitude** |
| Gertsch et al.[32] 2004 | Placebo-controlled RCT of 500 mg/day AZA or Ginkgo biloba or both on AMS (LLscore ≥3 including headache).<br>LLS at baseline at 4280; Altitude at 4358 m after the first night. | 303 (29.3% females) healthy trekkers. | 34% AMS with placebo; 12% AMS with AZA. RR of 64.7%, NNT of 4.5. | • **No sex-dependent AZA effect reported**<br>• **Females not characterized[1]**<br>• **Dropout rate of 33%**<br>• **No baseline <1500 m**<br>• **Trekkers already acclimatized**<br>• **No sex-dependent AZA effect reported**<br>• **1993 LLS version for AMS definition[2]**<br>• **Assessment <48 hours at target altitude** |
| **No females included** | | | | |
| Hillenbrand et al.[28] 2006 | Placebo-controlled RCT of 250 mg/day AZA on AMS (LLscore ≥3 including headache).<br>LLS at baseline 3440 m; Altitude at 4930 m on arrival. | 400 (0% females) male Nepali porters | 11.1% AMS with placebo; 12.7% AMS with AZA; | • **No females included**<br>• **Dropout rate of 69%**<br>• **No baseline <1500 m**<br>• **Porters already acclimatized**<br>• **1993 LLS version for AMS definition[2]**<br>• **Assessment <48 hours at target altitude** |
| Moraga et al.[30] 2007 | Placebo-controlled RCT of 500 mg/day AZA or Ginkgo on AMS (LLscore ≥3 including headache).<br>LLS at baseline at 0 m; Altitude at 3696 m during 72 hours. | 36 (0% females) healthy adults residing <1000 m. | 56.0% AMS with placebo; 36.0% AMS with AZA; RR of 35.7%, NNT of 5. | • **No females included**<br>• **1993 LLS version for AMS definition[2]** |

[1] Females were not characterized in terms of their reproductive status (pre- / peri- or postmenopausal), use of contraception), hormonal supplementation, or menstrual cycle phase during the stay at altitude. [2]According to the 2018 revised recommendations for defining AMS, a Lake Louise Questionnaire score of at least 3 points including at least mild headache and at least one other symptom of nausea and vomiting, fatigue and/or weakness, dizziness

and/or light-headedness within the first 48 hours at high altitude, is defined as AMS.[15] RR, risk reduction; NNT, number needed to treat; AMS, acute mountain sickness; AZA, acetazolamide; RCT, randomized clinical trial; LLS, Lake Louise Questionnaire.

#### 2.2. Current state of own research

**The current project partners** have successfully collaborated in various research projects in the field of high altitude medicine and physiology, including the SNF-funded projects (32003B, 192048, 172980, 143875, 122081, IZK0Z3_168254). They have an outstanding expertise in carrying out major clinical trials in healthy and patients with respiratory disease at high altitude places worldwide. To strengthen their collaboration, they launched the "Swiss-Kyrgyz High Altitude Medicine and Research Initiative" endorsed by the University Hospital and the University of Zurich, the Kyrgyz Ministry of Health and the National Center of Cardiology and Internal Medicine (NCCIM) in Bishkek, Kyrgyz Republic.[51] The partners established several research facilities in Kyrgyzstan offering excellent infrastructure for clinical trials: the National Center of Cardiology and Internal Medicine (Bishkek, 760 m), the Tuja Ashu High Altitude Clinic (Tuja Ashu pass, 3'100 m), the Kumtor Gold Mine Operation Facility (Issyk Kul Oblast, 3600 ), and at the Aksay Health Post (3'200 m).

**Dr. sc. ETH Michael Furian** This proposal consolidates and exploits many years of my own experience and expertise in conducting and leading major international studies and clinical trials in Switzerland, Chile, Peru, France, Ecuador, Antarctica and Kyrgyzstan on high-altitude physiology and medicine. This proposal aims to foster my independency in the field of high altitude medicine and physiology. Until today, I have investigated AMS,[36,37] sleep-disordered breathing,[38,39] exercise performance,[40-42] cerebrovascular reactivity,[43,44] and the effects of acclimatisation and repeated altitude exposure[45] in healthy lowlanders, patients with chronic obstructive pulmonary disease (COPD), and in highlanders permanently living at high altitude.[43,46] I led several randomised placebo-controlled trials as principal investigator investigating the preventive efficacy of nocturnal oxygen therapy and dexamethasone in patients with COPD staying at altitude.[38,39,44,47,48] In a land-mark trial in 185 patients with COPD and 345 healthy subjects ≥40 years staying for 2 days at 3100 m, we recently showed that prophylactic acetazolamide therapy reduces altitude-related adverse health effects in COPD patients and AMS in healthy individuals.[7] This trial further suggested that females were at increased risk of altitude-related illnesses and that the efficacy of acetazolamide might be sex-dependent.

**Prof. Dr. med. Konrad Bloch (KEB)** KEB has a longstanding experience in performing high altitude field studies including in Switzerland (Capanna Regina Margherita, Jungfrauoch, Davos, St. Moritz) and abroad. This research on effects of altitude on cardio-respiratory function,[49,50] exercise, sleep,[51-54] cognitive performance,[55,56] acclimatization and altitude-related illness in healthy individuals[51,57,58] is internationally well recognized. KEB was among the first to perform randomized, placebo-controlled trials in patients with respiratory conditions going to high altitude. These studies established current treatment recommendations for patients with obstructive sleep apnea travelling to altitude.[59-61] KEB has also performed physiological and clinical studies in COPD patients at low and high altitude.[40,41,62] His team performed the first randomized, placebo-controlled trials evaluating prevention of ARAHE in COPD patients with dexamethasone and AZA.[7,36,63-68]

**Prof. Dr. med. Silvia Ulrich (SU)** is director of the Dept. of Respiratory Medicine, University Hospital of Zurich. She has extensive clinical experience in respiratory and internal medicine and in particular in the invasive and noninvasive diagnosis of pulmonary hypertension (PH), treatment of PH, exercise and training enhancing effects of oxygen in cardiorespiratory

diseases and effects of hypoxia in patients with cardiorespiratory diseases and highlanders[63,68-91]. SU conducted several investigator-initiated studies on the pathogenesis and treatment of PH including several placebo-controlled trials on oxygen therapy in PH-patients,[66-68] exercise hemodynamics and exercise limiting factors in PH,[69,70] and the association of sleep disordered breathing with PH, [66] and acute and subacute effects of acetazolamide on hemodynamics and clinical outcomes in PH patients.[78,89,91,92] SU has performed extensive research in high altitude physiology and medicine with a special focus on the pulmonary circulation at rest and during exercise and patients with pulmonary vascular disease going to altitude and extensive collaborative studies with the applicant and collaborators.[69,70,85,86,93-95]

**Prof. Dr. med. Talant Sooronbaev (TS)** is director of the National Center of Cardiology and Internal Medicine (NCCIM) and head of the Dept. of Respiratory Medicine in Bishkek, Kyrgyzstan. He is Chief Pulmonologist in the Ministry of Health of the Kyrgyz Republic and member of the National Medical Research Council. TS directs the research facilities operated by the "Swiss-Kyrgyz High Altitude and Medicine Initiative". He performed various studies on cardio-respiratory disease in Kyrgyz highlanders[96-98] and is a main collaborator in various altitude studies conducted by our project team. TS has also studied the prevalence of COPD in high altitude residents in the Burden of Obstructive Lung Disease study (www.boldstudy.org) and in the FRESH AIR study http://www.theipcrg.org/freshair.

**PD Dr. med. Cornelia Betschart (CB)** is a gynecologist and obstetrician at the University Hospital Zurich since 2008 and board-certified urogynecologist since 2016. She is interested in AMS as a long-standing member of the Swiss Alpine Club. Scientifically she conducts several clinical multicenter observational, and prospective randomised placebo-controlled trials as principal investigator, also granted by the SNF and private foundations. She promotes inter-professional and inter-disciplinary research as founding member of the International Collaboration for Harmonising Outcomes, Research, and Standards in Urogynaecology and Women's Health (CHORUS), as president of the Swiss Urogynecological Association, steering-board member of Gender medicine USZ, co-founder and leader of Pelvic Floor Center USZ and deputy chairwoman of the Department of Gynecology USZ.

**Dr. sc. hum, MSc. med. Biometry & Statistics Nicola Benjamin (NB)** is the Head of study coordination and scientific project management of the Center for pulmonary hypertension at the University Clinic of Heidelberg, Germany. NB has a broad and extensive knowledge on clinical trial design and statistical analyses in the field of Pulmonology (including hypoxia and hypoxemia).[99-101] She contributed to the trial design and statistical considerations of this application, furthermore, NB will supervise the creation of the statistical analysis plan, the interim analysis and the final statistical analyses of this project.

**EXALT Research Consortium** is composed of outstanding high altitude researchers situated in France. These researchers, namely Prof. Dr. Samuel Vergès (Université Grenoble Alpes), Prof. Dr. Julien Brugniaux (Université Grenoble Alpes), Dr. Benoit Champigneulle (Université Grenoble Alpes), Prof. Dr. Aurélien Pichon (Université de Poitiers), Prof. Dr. Paul Robach (Ecole Nationale de Ski et d'Alpinisme), Dr. Emeric Stauffer (Hospoces Civils de Lyon), have conducted landmark studies in France and in Peru (Expedition 5300 m).[102-106] They are specialized in comprehensive physiological and basic research at altitudes up to 5300 m. Due to SNF Postdoc.Mobility grant in 2020, the main applicant joined this team in Peru, La Rinconada in 2021 and invited them to research studies in Kyrgyzstan.[107,108] They will strongly contribute to the physiological project aims to understand physiological and mechanistic sex-differences in altitude tolerance and mechanisms of action of acetazolamide.

#### 2.2.1. Insights from two preceding pilot studies in females

Assessing sex-related differences at high altitude require meticulous planning, translational knowledge in interdisciplinary research fields and established high altitude research facilities. Therefore, to gain important insights into the expected challenges of high altitude studies in females, and to enhance the feasibility of the proposed project, the main applicant of this proposal conducted two pilot studies in overall 42 healthy premenopausal females in 2022. All females were instructed to monitor their urinary hormone concentrations for 30 consecutive days by the *Full Cycle Hormone Insights Kit* (proov, MFB Fertility Inc., CO, US) starting the day before traveling to 3100 m or 3600 m and staying there for 2 days/nights. These pilot studies revealed excellent hormone monitoring adherence, with 1201 of 1230 (97.6%) successful hormone analyses, providing a robust and reliable hormone measurement technique. The averaged and smoothed hormone profile illustrated in **Figure 2, Panel A** shows that ascending to high altitude, independent of the MCP, results in the expected random distribution of participants throughout the Cycle Days (**Figure 2, Panel B**). This important finding confirms that any large-scale randomized clinical trial does not require to schedule altitude ascents based on the Cycle Day or MCP. The equal distribution of the participants throughout the Cycle Days are a clear strength of the study design, since it avoids any biased conclusion based on comparisons of a few isolated Cycle Days in the luteal compared to follicular MCP (in case randomization is based on specific Cycle Days or MCP).

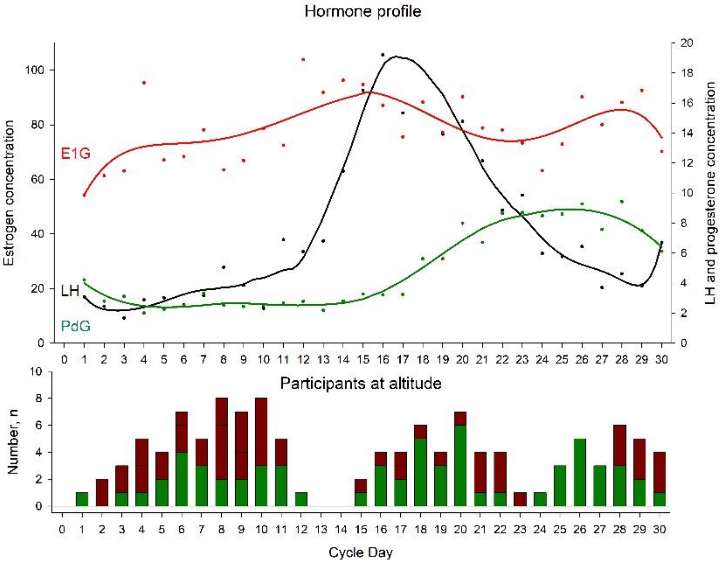

***Figure 2: Urinary hormone profile from 42 females travelling to high altitudes. Panel A:*** *Females daily monitored their urinary hormones E1G, LH and PdG for 30 consecutive days. Based on their first day of the menstruation, the individual Cycle Day at altitude ascent was calculated. Hormone concentrations were averaged for each Cycle Day and a curve was fitted for each hormone.*

***Panel B:*** *The total number of participants staying at altitude and in relation to their individual Cycle Day. The green proportion of the bars represent females without AMS; red proportion of the bars represent females with diagnosed AMS.*

In addition, when comparing the findings at 3100 to 3600 m (**Table 2**), the pilot studies provide important quantitative data related to AMS incidences. Therefore, an AMS incidence of 57% can be expected in premenopausal females staying for 2 days/nights at 3600 m.

**Table 2. Main findings of the two pilot studies.**

| | Group 1 (n = 21) | | | Group 2 (n = 21) | | | Mean difference in altitude effect (3600 m vs 3100 m) (95% CI) |
|---|---|---|---|---|---|---|---|
| | 760 m | 3100 m | Mean altitude effect (95% CI) | 760 m | 3600 m | Mean altitude effect (95% CI) | |
| **Clinical examination** | | | | | | | |
| Systolic BP, mmHg | 106 ± 2 | 106 ± 2 | 0 (-3 to 3) | 104 ± 2 | 105 ± 2 | 1 (-2 to 5) | 1 (-3 to 6) |
| Diastolic BP, mmHg | 73 ± 2 | 74 ± 2 | 1 (-3 to 5) | 70 ± 2 | 76 ± 2 | 6 (2 to 10)* | 5 (0 to 10) |
| Heart rate, bpm | 85 ± 2 | 87 ± 2 | 2 (-3 to 7) | 78 ± 2 | 95 ± 2 | 17 (12 to 22)* | 15 (8 to 23)* |
| $SpO_2$, % | 96.1 ± 0.4 | 93.0 ± 0.4 | -3.1 (-4.2 to -2.1)* | 97.3 ± 0.4 | 86.8 ± 0.4 | -10.5 (-11.6 to -9.4)* | -7.3 (-8.8 to -5.8)* |
| Acute mountain sickness | | | | | | | |

| | | | | | | | |
|---|---|---|---|---|---|---|---|
| 2018 Lake Louise Score | 0.0 ± 0.3 | 0.3 ± 0.3 | 0.3 (-0.4 to 1.0) | 0.0 ± 0.3 | 1.9 ± 0.3 | 1.9 (1.2 to 2.6)* | 1.6 (0.6 to 2.6)* |
| 2018 AMS incidence, % | 0% | 43% | | 0% | 57% | | |

BP, blood pressure; $SpO_2$, arterial oxygen saturation assessed by finger pulse oximetry.

#### 2.3. Detailed research plan

##### 2.3.1. Purpose

The primary purpose of this project is to compare the efficacy of preventive acetazolamide treatment against AMS in females compared to men. An important secondary purpose will be to compare the AMS incidence in females versus males at 3600 m. Additional purposes are to investigate sex- and MCP-related differences in altitude tolerance and efficacy of acetazolamide prevention and to underpin clinical findings with various physiological measures providing important mechanistic insights. Findings will improve our understanding of AMS pathophysiology and will provide scientific evidence whether sex-related recommendations on altitude travel and prevention of altitude illness are necessary.

##### 2.3.2. Primary Hypothesis

The primary hypothesis will be that, during the time course of 2 days/nights at 3600 m, preventive 250 mg/day acetazolamide therapy starting 24 hours before ascending, will reduce the AMS incidence significantly more in females compared to men. AMS will be defined as a Lake Louise questionnaire score of ≥3 points including headache.[14]

##### 2.3.3. Additional Hypotheses

Altitude effects in females

1) Females under placebo staying at 3600 m experience altitude-induced exercise intolerance after arrival at 3600 m compared to 760 m assessed by a maximal cardiopulmonary bicycle exercise test.

Altitude effects between females and men

2) Females under placebo have a higher AMS incidence compared to males under placebo during the time course of 2 days and nights at 3600 m.
3) Females under placebo have less sleep-disordered breathing during the first night at 3600 m compared to males under placebo assessed by respiratory polygraphy.

Acetazolamide effects in females

4) Acetazolamide therapy mitigates altitude-induced exercise intolerance compared to placebo.

Acetazolamide effects between females and men

5) The acetazolamide plasma concentration in the morning after the first night at 3600 m is higher in females compared to men.
6) Perceived acetazolamide-related side effect are higher in females compared to males during the time course of 2 days and nights at 3600 m.

MCP-related differences

7) In females under placebo staying at 3600 m during the luteal MCP, the AMS incidence; altitude-induced exercise intolerance and cardiac repolarization disturbances are lower compared to females during the follicular MCP.

##### 2.3.4. STUDY DESIGN AND SETTING

This is a randomized, placebo-controlled, double-blind, parallel trial (**Figure 3**) conducted over 2 consecutive summer seasons. Healthy females and males (according to physician assigned sex at birth) will perform measurements at the National Center of Cardiology and Internal Medicine, Department of Respiratory Medicine, Bishkek (760 m), Kyrgyzstan and in the Kumtor High Altitude Facility (3600 m), Kyrgyzstan. After written consent of female participants, they will be randomized to either acetazolamide or placebo and will start the sexual hormone monitoring in the morning urine as described in the **section 2.3.6** below. Baseline measurements at 760 m will be conducted 24 hours before ascending to 3600 m and altitude measurements will be performed while staying for 2 days and nights at 3600 m. Male participants will undergo the same measurements at 760 and 3600 m but will not monitor hormones. Transfers between locations will be performed by minibus within 6 hours.

##### 2.3.5. PARTICIPANTS

Inclusion criteria for female participants are premenopausal, eumenorrheic, non-smoking, healthy females with a BMI >18 kg/m$^2$ and <30 kg/m$^2$, aged 18 to 44 years, and who live at altitudes <1000 m. Exclusion criteria are any pre-existing diseases, regular intake of medication (including oral contraceptives), other types of contraceptives (hormonal intrauterine device, vaginal ring, subcutaneous injections or implants, among others), pregnancy or nursing, anaemic (haemoglobin concentration <10g/dl), and any altitude trip <4 weeks before the study. Additionally, all included females require a mobile phone compatible with the hormone monitoring app *proov*. Male participants fulfilling the above-mentioned, male-applicable inclusion and exclusion criteria will be recruited. Participants will be recruited by several teams advertising the study at various universities in Bishkek and in the surrounding villages. This recruitment procedure has been applied in several previous randomized clinical trials and has been proven to be extremely successful. The protocol will be submitted to the Ethic Committee of the National Center of Cardiology and Internal Medicine, Bishkek, Kyrgyzstan and to the Cantonal Ethics Committee of Zurich, Zurich Switzerland. Participants will be asked to provide written informed consent.

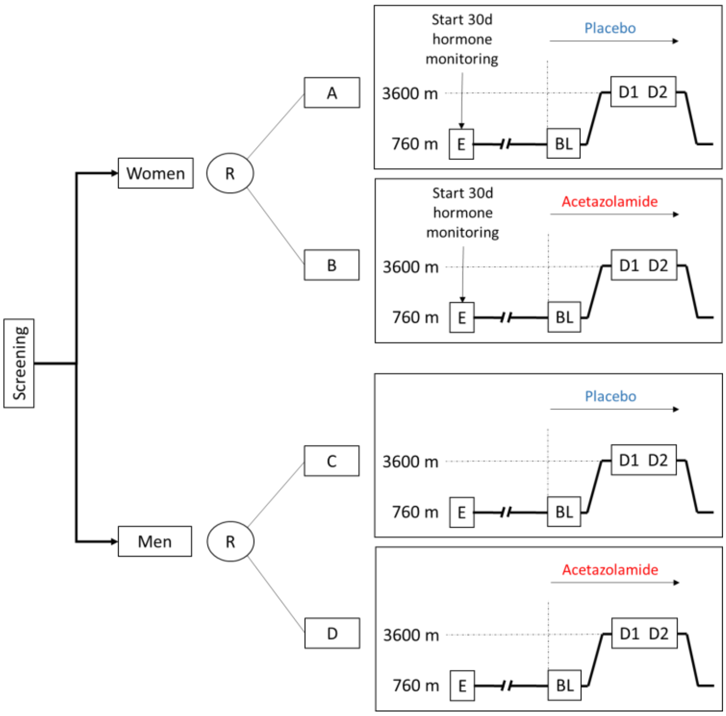

***Figure 3: Study design.*** *Preventive 250 mg/day acetazolamide will be administered 24 hours before travelling and staying for 2 days/ nights at 3600 m. AMS incidence will be assessed at 3600 m. Female participants will monitor urinary levels of sexual hormones for 30 days allowing to determine the MCP during the stay at 3600 m. E, study entry; BL, baseline; R, randomization; D1 – 2, day 1 to 2.*

##### 2.3.6. MEASUREMENTS DURING STUDY VISITS

The schedule for the measurements is illustrated in **Table 3**.

| Table 3. Assessment schedule | | | | | | |
|---|---|---|---|---|---|---|
| | Screening | BL, Day 1, 760 m | BL, Day 2, 760 m | Day of ascent, 760 m / 3600 m | Day 1, 3600 m | Day 2, 3600 m |
| Time before ascent to 3600 m | up to -5 month | -1 week | -1 day | 0 | + 1 day | +2 days |
| Assessment | | | | | | |
| Informed consent (NCCIM) | x | | | | | |
| In-/exclusion criteria (NCCIM) | x | | | | | |
| Reproductive status (NCCIM) | x | | | | | |
| Clinical examination (USZ) | x | x | x | x | x | x |
| AMS questionnaires / Visualiz. (USZ) | | x | x | x | x | x |
| Respiratory polygraphy (USZ) | | x | | x | x | |
| Iron status (USZ, France) | | x | x | | x | |
| Hypoxic ventilatory response (USZ) | | x | | | | |
| Cerebrovascular reactivity (France) | | x | | | x | |
| Echocardiography (USZ) | | x | | | x | |
| Hemorheology (France) | | x | | | x | |
| OpCo CO-rebreathing (France) | | x | | | x | |
| Fundoscopy, Pupillometry (USZ, France) | | x | | | x | |
| Acetazolamide concentration (Italy) | | x | x | | x | |
| Urinary hormone testing (USZ) | | 30 consecutive days by urine hormone sampling | | | | |

Screening visit

After providing written informed consent, a complete medical history will be obtained. A reproductive status questionnaire will assess regularity of menstrual periods, menstrual cycle length, menarche, pregnancy and breastfeeding, use of contraception, use of other hormone supplementation and known gynecologic or other medical conditions. Inclusion and exclusion criteria will be applied. Physical examination will include weight, height, blood pressure, heart rate, and cardiac and pulmonary auscultation. A pregnancy test will be performed.

History, symptoms, and clinical examination

Repeated physical examinations will include weight, height, blood pressure, heart rate, and cardiac and pulmonary auscultation. AMS will be assessed using the current and the 1993 version of the Lake Louise score[14,109] and the environmental symptoms questionnaire (AMS-c).[110] Subjective sleepiness and sleep quality will be assessed using the Karolinska Sleepiness scale and a 100-mm visual analogue scale. A standardised headache diary including information about the date, time, intensity (1 to 10), preceding symptoms, triggers and relief of headache will be distributed. This diary will be incorporated in the 30-day urinary hormone monitoring and will also be distributed to male participants.

Urinary hormone monitoring

Females will monitor the urinary hormone concentrations of estrone-1-glucuronide (E1G), pregnanediol-3-alpha-glucuronide (PdG) and luteinizing hormone (LH) for 30 consecutive days by the FDA approved *Full Cycle Hormone Insights Kit* (proov, MFB Fertility Inc., CO, US) starting before traveling to 3600 m. The *proov* hormone monitoring kit has been used in previous research studies and has been validated against other methods for hormone monitoring.[111] Measurements will be standardized in the morning, after awakening and before drinking or eating breakfast. The *proov* multi-hormone test strip lateral flow assay uses gold nanoparticles and buffered sample pads designed to adjust for pH and hydration levels, filters unwanted particulates and binds contaminants in urine that may interfere with the accuracy of the test. The strip contains three test lines and one control line, corresponding to E1G, PdG and LH (beta subunit). After waiting for 10 minutes, the user provides a photo of her urine test strip with the *proov* app. The application server uses machine learning specifically designed to analyse photographed images of the test strip, control for variations in camera, lightning, and operating system, check for input or output irregularities, and mathematically derive the associated hormone levels.[111] In case of a failure of the analysis due to loss of internet, then an offline analysis by the company is feasibly by providing the time, day and picture of the test strip of the participant. After the completion of the 30-day urine hormone monitoring and in consideration the first day of the last menstrual bleeding, Cycle Days and the participants' MCP will be defined by visually inspecting the 30-day hormone profile. The follicular phase will be defined from the Cycle Day 1 (first day of menstrual bleeding) until the LH peak; the luteal phase will be defined as the day after the LH peak until the day before the next menstrual bleeding.

<u>Hypoxic-ventilatory response test</u>

The hypoxic ventilatory response at exercise (HVRe) has been suggested to be a good predictor for AMS.[19,112] Especially in premenopausal females, it has been suggested that HVRe is higher in the luteal compared to the follicular phase, providing

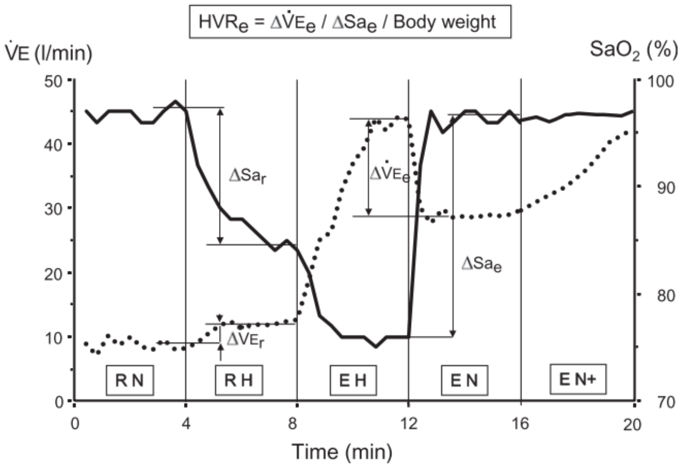

***Figure 4: Hypoxic exercise test.*** *Figure 1 from Richalet et al. 2012.*[104] *RN, RH, EH, and EN represent rest in normoxia, rest in hypoxia, exercise in hypoxia, and exercise in normoxia, respectively. Hypoxia will be produced by breathing normobaric hypoxic gas (fraction of inspired oxygen of 0.115). Exercise will be performed at 30% of maximal exercise capacity in phases EH and EN. In phase EN+, the exercise intensity will be adjusted so that the heart rate reaches the same value as in EH. HVRe, ventilatory response to hypoxia during exercise; $SaO_2$, arterial oxygen saturation assessed by finger pulse oximetry; $Sa_e$, arterial oxygen saturation during exercise; $Sa_r$, arterial oxygen saturation at rest; V'E, minute ventilation; $V'E_e$, minute ventilation during exercise. Dotted line represents V'E; solid line represents $SaO_2$.*

a potential underlying factor explaining MCP-related differences in AMS susceptibility. Therefore, subjects will perform an exercise test consisting of 5 consecutive phases: 1 – rest in normoxia; 2 – rest in hypoxia (FiO2 of 0.115 equivalent to 4800 m); 3 – exercise in hypoxia (EH) at 30% of maximal normoxic maximal exercise capacity (Ergoselect 200; Ergoline GmbH, Bitz, Germany); 4 – exercise in normoxia (EN) and 5 – EN with the same heart rate achieved during EH (EN+). Arterial oxygenation by finger pulse oximetry and respiratory gas concentrations, tidal volume, and respiratory rate will be measured breath-by-breath by a metabolic unit (Ergostick; Geratherm Medical AG, Gschwenda, Germany) to compute minute ventilation (V'E) and other outcomes. HVRe will be calculated as previously described and illustrated in **figure 4**.

<u>Haemoglobin mass ($Hb_{mass}$) and intravascular volumes</u>

Plasma volume (PV) contraction is an early mechanism allowing an increase in haemoglobin concentration ([Hb]) (and hence, in arterial oxygen content) after an acute high-altitude exposure, and thus, counteracting the decrease in oxygen availability.[113] Experimental studies distinctly conducted in both male and female voluntaries confirmed that this decrease in PV occurred early (*i.e.*, in the first 12-24-hours) and with a similar magnitude between sex, mainly through a fluid-redistribution mechanism from the intra to the extravascular compartment, rather than due to a hypoxic diuresis.[114,115] Beneficial effects of a preventive acetazolamide uptake on AMS are assumed to be mostly driven by the consecutive hyperventilation balancing the induced metabolic acidosis and, thus, increasing the arterial oxygen content; however, even at a such low dose, acetazolamide might have beneficial effect on arterial oxygen content, through its diuretic effect, by further decreasing the PV.[116] To investigate PV changes, an $Hb_{mass}$ measurement using the CO-rebreathing method, allowing the computation of PV, will be conducted at 760 m and after 12 hours staying at 3600 m. Briefly, the CO-rebreathing test will be conducted as previously described, using an automated system (OpCo, Detalo Health, Copenhagen, Danemark), after a 20-min rest period in supine position.[117] From a capillary blood sample, [Hb], pre-test percentage of carboxyhaemoglobin (Rapidpoint 500, Siemens AG, Zürich) and haematocrit (Hct, microcentrifuge method) will be determinate in quadruplicate. Next, while the participant will breathe a 100% mixture of oxygen in a rebreathing circuit, a bolus dose of 99.997% pure CO gas (1 mL per kg body mass for men, 0.8 mL per kg body mass for females) will be automatically administered by the Opco system. After a 6-min rebreathing period, the participant will be disconnected from the rebreathing circuit and the residual CO concentration in the circuit will be measured. Then, a secondary capillary blood sampling will be performed at 10 min, to measure, in quadruplicate, the post-test %HbCO (%$HbCO_{POST}$). From the post-pre-test difference in %HbCO and the amount of absorbed CO, the $Hb_{mass}$ value will be calculated and the derived intravascular volumes (PV, red blood cell volume and total blood volume) will be calculated.[117] Previous studies conducted in the field in high-altitude environment have confirmed both the feasibility and the safety of the CO-rebreathing method.[105,118,119]

Hemorheological analysis

Potential effects of a prophylaxis acetazolamide uptake in order to prevent AMS on blood viscosity could be leaded by an expected decrease in PV or by a direct effect on red blood cell (RBC) deformability due to a direct inhibition of RBC carbonic anhydrase,[113] albeit hemorheological evaluations have never been performed in this context. To explore the potential hemorheological effect of acetazolamide prophylaxis administration, and the potential difference of effect between males and females, we will plan to perform blood viscosity and RBC's deformability measurements at 760 m and after the first night at 3600 m. At both altitudes, from a 3 mL venous sample collected in an EDTA tube, the following analyses will be performed, in accordance with the guidelines for hemorheological laboratory techniques:[120]

- Blood viscosity will be measured at native and corrected Hct (40%, after dilution using autologous plasma) at several shear rates (22.5 $s^{-1}$, 45 $s^{-1}$, 90·$s^{-1}$).
- RBC's deformability will be measured by ektacytometry using the Laser Optical Rotational Red Cell Analyzer (LORRCA, RR Mechatronics, Hoorn, The Netherlands) at 3 and 30 Pa and the extent of RBC aggregation and the strength of RBC aggregates will be measured by syllectometry using the LORRCA (RR Mechatronics, Hoorn, The Netherlands) after adjustment of the Hct to 40% with autologous plasma.

Arterial and venous blood sampling

Effects of altitude and acetazolamide on arterial blood gases will be assessed in a radial artery sample after 15 minutes of quiet and awake supine position in the morning after awakening. As previously described, arterial blood will be analysed for pH, $PaO_2$, $PaCO_2$, $SaO_2$, haematocrit and electrolytes (Rapidpoint 500, Siemens AG, Zürich).[38] To quantify and confirm MCPs, absolute hormone concentrations and plasma concentration of acetazolamide and to provide mechanistic explanations to PV changes, a 20 mL peripheral venous blood sample in the morning after awakening will be withdrawn, centrifuged, and the plasma/serum immediately frozen, stored, and transported at -20°C for further analyses. Analysis at 760 m will include iron metabolism (ferritin, TSAT, hepcidin, erythroferrone, sTfR1). At both altitudes, circulating reproductive hormone concentrations (oestradiol, progesterone, LH) , plasma renin activity, aldosterone, copeptin (as a surrogate of the antidiuretic hormone) and midregional proANP (the precursor of the atrial natriuretic peptide) will be analysed. The plasma concentration of acetazolamide will be assessed using dried blood spots.[121] Therefore, 80-100 µl venous blood from the sample will be pipetted on to four spots on the dried blood spot card and analysed as previously described.

Cardiopulmonary exercise testing and electrocardiogram

As previously performed by our project team, a resting 12-lead ECG will be performed after resting for 10 minutes in a supine, awake position which will give insights into cardiac repolarization disturbances at 3600 m.[70] Thereafter, a bicycle exercise test with a progressive ramp protocol will be performed until exhaustion (Ergoselect 200, Ergoline GmbH, Bitz, Germany) as previously performed in patients with COPD and healthy;[40,67] and according to international guidelines to assess alterations in physical exercise performance at altitude and under acetazolamide.[42,122] Participants will rest on the bicycle for 10 minutes, followed by a 10 – 20 W/min work-load increase starting at 20 W. Respiratory gas concentrations, tidal volume, and respiratory rate will be measured breath-by-breath (Ergostick, Geratherm Medical AG, Gschwenda, Germany). Arterial oxygenation (by finger oximetry) and a 12-lead ECG will be applied. Before and at peak exercise, dyspnoea sensation and leg fatigue will be assessed with the BORG CR10 scale.[123]

Cardiorespiratory sleep studies

Continuous nocturnal measurements by portable devices (Alice PDX, Philips Respironics, Zofingen, Switzerland) will include nasal pressure swings, chest wall excursion, ECG, and pulse oximetry. Mean oxygen saturation and percentage time with $SpO_2$ <90% and <80% will be computed. The apnoea/hypopnea index and oxygen desaturation index ($SpO_2$ dips >3%) will be computed as mean number of events/h.[52] These measurements will give valuable insights into sleep-disordered breathing often seen at altitude and the effect of acetazolamide. This measurement has been successfully implemented by our project team in patients with COPD and healthy lowlanders and highlanders.[7,124]

Cerebrovascular reactivity assessment

Participants will undergo transcranial near-infrared spectroscopy (NIRS) measurements of the prefrontal cortex, finger pulse oximetry, transcranial Doppler ultrasound (TCD), electrocardiography (ECG), measurement of continuous blood pressure, end-expiratory $PCO_2$ measurement and breathing pattern while the participant is sitting in a comfortable position for 10 minutes breathing room air. After baseline measurements, the participant will be asked to perform squat jumps, hyperventilate and reduce their expired $PCO_2$ by 10 mmHg or do an isometric exercise by pressing a predefined load in the dominant hand by a hand grip dynamometer for 1 minute, in random order. The TCD probe will be used to insonate the middle cerebral artery on the same side as the hand grip maneuver is performed to eliminate neurovascular activity contamination. Between interventions a 5-minute wash-out phase will be implemented to avoid any carry-over effect. The last 2 minutes of quiet breathing room air will be used as baseline measurement and will be compared with hyperventilation in participants under placebo compared to participants under acetazolamide. Furthermore, blood flow response in the middle cerebral artery due to blood pressure alterations will be compared between altitudes, drug and sex.

To complement TCD's assessment of regional cerebral blood flow (CBF) velocity through the middle cerebral artery, global CBF will be assessed during the same procedures using Duplex ultrasound of both the right internal carotid and the right vertebral arteries (ICA and VA, respectively). Combined diameter and blood velocity measurements provide the necessary data to calculate blood flow through these vessels. Owing the ICA and VA are bilateral, the calculated blood flow will then be multiplied by a factor of 2 to obtain a value for total CBF.

Biobeat Wrist Monitor

The non-invasive biobeat Wrist Monitor continuously monitors vital parameters including heart rate, breathing frequency, arterial oxygenation and systolic and diastolic blood pressure. The patented algorithm is based on artificial intelligence and machine-learning and has been used in other scientific settings.[125] A unique feature is that the vital parameters are not displayed on the Wrist Monitor, instead, are uploaded to the cloud and are live accessible by the physician through a remote patient monitoring system. This approach minimizes confounding the participant and allows high-quality remote monitoring of the participants. Moreover, vital parameters will be continuously recorded and might allow in the future to use an early warning system to prevent AMS. During this study, each participant will wear a biobeat Wrist Monitor during the complete duration of the altitude sojourn. The continuous data recording will be modelled and analysed by partners of the ETH Zurich.

Lung ultrasonography

Using a phased array probe, bilateral lung ultrasound will be performed in a supine from the second to fourth or fifth intercostal space (left and right hemithorax, respectively) down the parasternal, mid-clavicular, anterior axillary and mid-axillary lines, resulting in 28 total windows of interest (left: 12; right: 16). A B line will be defined as one echogenic, continuous, wedge-shaped, signal arising from the uninterrupted pleural interface with a narrow origin in the near field of the image. The number of B-lines will be counted for each lung field and totalled.[76]

Echocardiography

Cardiac morphological and functional parameters will be assessed by standard two-dimensional Doppler echocardiography. Doppler imaging of the tricuspid annulus and the pulmonary outflow tract will be assessed according to standard methods. This will allow us to assess the following parameters: systolic pulmonary artery pressure ($RV_{sys}$, calculated from the tricuspid regurgitation velocity by using the modified Bernoulli equation: $RV_{sys}$ = 4x ($v_{max}$) where $v_{max}$ is the maximum of the regurgitation velocity jet measured over the tricuspid valve) and the right ventricular outflow tract acceleration time; the right ventricular fractional area change [%] and the tricuspid annular plain systolic excursion [mm]. The fractional area change of the right ventricle will be determined. The tricuspid annular plane systolic excursion (TAPSE) will be measured in M-mode. The stroke volume will be measured based on the left ventricular outflow tract diameter and the velocity time integral over the aortic valve in the apical 5- chamber view or the apical long axis view.

Optic nerve sheath diameter (ONSD)

ONSD measurement will be done with a 10 Mhz ultrasound probe using a portable ultrasound machine (uSmart 3300, Terason, USA). Briefly, a thick layer of gel will be applied on the upper closed eyelid before the transducer will be positioned on it, avoiding any excessive pressure, on the temporal side of the eye. Positioning the transducer in the transverse axis, a first measurement will be done in triplicate; then, the transducer will be positioned in the longitudinal axis, to perform three other measurements. Measurements will be done 3 mm behind the ocular globe. The average value obtained for each eye from the six measured values (three in the transversal and three in the longitudinal axis) will be used for the further calculations. To reduce the measurement time, the ultrasound motions will be captured, and the measurements will be conducted in a second time. Duration of the measurement is estimated to one minute by eye.

Automated Pupillometry (AP)

Quantitative pupillometry will be performed using the NeurOptics® pupillometer (NeurOptics, Irvine, CA, USA) on both eyes (right, then left), following the ONSD measurement. The NeuOptics® pupillometer delivers a standardized light stimulation of fixed intensity (1000 Lux), allowing a rapid and precise measurement (0.05 mm limit) of the pupil size changes.[126] The following pupil parameters, measured by the NeurOptics® pupillometer will be recorded : pupil size (mm) before and after light stimulation, percentage of constriction (%), latency of constriction (s), constriction velocity (mm·s-1) and dilatation velocity (mm·s-1) and NPi (neurological pupil index, a computed parameter provides by the device and based on the previously cited parameters). A special attention will be paid to obtain standardized ambient light conditions (i.e., dark conditions) between the two places to avoid a potential confounder between the LA and the HA measurements, as ambient light level has been shown to impact pupil parameters, even in healthy subjects, excepted for latency time.[127]

Funduscopy

We conduct fundus photography based on handheld retinal imaging system (VistaView™ by Volk optics) in healthy lowlanders ascending to 3600 meters above sea-level. We hope to gain insight in the feasibility and usefulness of retinal imaging for detection of papilledema as an early sign of increased intracerebral pressure. To ensure optimal recording conditions, mydriasis will be induced by topical application of atropine drops.

Pain visualization with SMaRT App

Pain sensation of AMS and any other pain-related symptoms will be recorded using the Sensation Mapping and Reporting Tool (SMaRT) app on a tablet.[128] This app allows the participants to describe their sensations by sketching it inside the image of a body. This will help to assess the intensity, characterization and location of sensations. Whenever the participants fill out the Lake Louis questionnaire, they will also use the SMaRT app to assess their sensations. Recording these additional symptoms or sensations will help to give a more holistic view of AMS in the participants.

Traditional Chinese Medicine (TCM) tongue diagnosis

The TCM syndrome diagnostics are of interest in the context of AMS, given that the main symptom of AMS is headache. Acupuncture is known to be a non-pharmacological treatment option for headache and TCM syndromes are mainly based on the investigation of the tongue's shape, color and coating. However, before an interventional study with acupuncture can be planned, more information on the effect of hypoxia and AMS on the tongue appearance is required. In this study, we therefore take pictures of the participant's tongue at baseline and at high altitude. These pictures will then be analysed using machine learning algorithms analyzing the pictures in relation to hypoxia and AMS.

##### 2.3.7. Outcomes

The primary outcome of this study is the absolute difference in efficacy of acetazolamide to reduce AMS incidence in females compared to men. AMS will be defined as a 2018 Lake Louise questionnaire score of ≥3 points including headache.[15] If a participant scores ≥3 points while staying at 3600 m, the participant will be deemed to have AMS.

Secondary outcomes will be derived from urinary sexual hormone concentrations, AMS severity, AMS symptom components, and AMS defined by different questionnaires and cut-off scores.[109,110] Vital parameters, parameters from arterial and venous blood sampling, hypoxic ventilatory response during exercise, cerebrovascular reactivity and respiratory sleep studies will be obtained and compared between sexes, between acetazolamide and placebo and between MCPs. Additionally, ECG morphology, cardiac repolarization disturbances, sleep-disordered breathing and exercise performance and exercise-limiting factors will be obtained and compared. Mechanistic insights of the prophylactic effect of acetazolamide for AMS will be derived from intravascular volume measurements, $Hb_{mass}$, blood viscosity and hemorheological assessments.

##### 2.3.8. Randomisation, intervention and blinding

Stratified randomization will be conducted for females and males using a computer generated schedule (MinimPy 0.3), stratifying for sex in a females-to-males ratio of 2:1.[129] Within sex, participants will be 1:1 randomized to A: 250 mg/day acetazolamide (125 mg in the morning with breakfast, 125 mg in the evening with dinner) starting 24 hours before arriving and while staying for 2 days and nights at 3600 m (total of 6 capsules of 125 mg acetazolamide) or B: identically looking, organoleptic placebo capsules. An independent pharmacist will prepare identically looking verum and placebo capsules labelled with secret codes. The list of codes will be kept confidential to investigators and participants until data acquisition and analysis has been completed.

##### 2.3.9. Sample Size Estimation

Sample size estimation was performed using the Cochran Mantel-Haenszel test with continuity correction. The previously described preventive effect of acetazolamide against AMS has been suggested to be 44%.[34] To detect a clinically meaningful efficacy difference of 20% of acetazolamide therapy against AMS in females compared to males (i.e. treatment effect of 50% in females and 30% in men) with a group allocation ratio of 2:1, power of 90%, two-sided alpha level of 0.05 and accounting for dropouts, a total of 180 females and 90 males will be recruited. The females-to-males ratio of 2:1 was chosen to enable conclusive testing of the additional hypotheses within females (see **section 2.3.3**). The project team has successfully recruited even higher numbers of COPD patients and healthy participants in previous randomized clinical trials at high altitude. As an example, in 2015, 124 COPD patients were recruited within 1 summer expedition and included in the final analysis[36]. In another trial of acetazolamide, 186 COPD patients and 345 healthy subjects were recruited and included in the final analysis within 3 summer expeditions.[7] Since the complex recruitment of these large numbers of chronically ill patients was successful, the anticipated number of healthy females and males required for this study is well feasible and within the capacity of the project team.

##### 2.3.10. Data Analysis and Statistics

In accordance with international guidelines for prospective clinical trials,[130] a statistical analysis plan, similarly to the studies published by the applicant in *NEJM Evidence,*[7] will be created before any statistical analyses. This study will be pre-registered at ClinicalTrials.gov and results reporting will follow the CONsolidated Standards Of Reporting Trials (CONSORT) guidelines.[131]

Data will be summarised by numbers and proportions and means (SD) and medians (quartiles) for normally and non-normally distributed data, respectively. Altitude-induced differences will be calculated by mean or median differences (95% confidence intervals). The primary analysis will be performed on the intention-to-treat population including all randomized participants using the Cochran Mantel-Haenszel test comparing the sex-related difference in the acetazolamide effect against AMS.[132] In case of missing values in the primary outcome (AMS, binary), a sensitivity analysis with best / worst case scenario will be applied. Kaplan-Meier curves for AMS will be plotted for the four groups representing the combination of sex and treatment assignment: 1) males randomly assigned to receive acetazolamide; 2) males randomly assigned to receive placebo; 3) females randomly assigned to acetazolamide; 4) females randomly assigned to placebo. Comparisons will be performed using a log-rank test. Secondary, a multivariable Cox proportional-hazards analysis will be conducted to determine whether the interaction between sex and acetazolamide therapy is independent of other baseline factors.

Secondary outcomes will be analysed on the per-protocol population defined as participants with available data. Continuous variables will be analysed using mixed linear regression models, with the variable of interest as the dependent variable and drug, location, sex and MCP as fixed effects, including the interaction term drug*location*sex*MCP.

To proactively minimize any unforeseen deviations from the anticipated study progression, such as extreme benefit or harm of acetazolamide or futility and unexpected AMS incidences, a planned interim analysis will be conducted after the completion of the first study year. The planned interim analysis will be performed by an independent statistician blinded to drug. The statistician will report the blinded results to the project steering committee (composed by the applicant and the co-applicants), who will decide on continuation, termination, or on any appropriate protocol adaptation (e.g., sample size re-estimation). Termination of the study is defined by symmetric stopping boundaries at $p<0.001$ (Peto approach).[133] In the final analysis, a $p<0.05$ will be considered statistically significant.

##### 2.3.11. Further considerations

As in previous studies over the past few years, the applicant has access to a dedicated and experienced Swiss and Kyrgyz research team composed of assistant physicians and medical students, as well as access to pivotal medical devices needed for the described measurements such as ergometers, respiratory polygraphy, computers, and other hardware. This equipment is available from the *Swiss-Kyrgyz High Altitude Medicine and Research Initiative*. Medical devices and consumables will be transported to Kyrgyzstan for the study duration. During data acquisition periods, up to 6 medical bachelors' students from ETH Zürich will perform their 6-week research internship in Kyrgyzstan. Prof. Silvia Ulrich will dedicate additional Swiss staff for preparing and conducting this project during the years 2024 and 2025. The Kyrgyz team led by Prof. Sooronbaev (The Kyrgyz director of the Swiss-Kyrgyz High Altitude Research Center and Director of the University Hospital in Bishkek) will contribute the required personnel to recruit participants and for data acquisition, facility maintenances at low and high altitude, and transportation.

In regard of the health and safety of the participants at 3600 m, the Kumtor Gold mine Facility owns a fully equipped medical station including supplemental oxygen, emergency medications, medical staff and ambulances to quickly evacuate to lower altitudes.

#### 2.4. Schedule and milestones

The schedule and milestones of the project are outlined in **Figure 5**. The project partners will meet several times per year to decide on scientific, logistic and administrative aspects and other questions that might arise. The study preparations will be started on January 2024 and will recruit 140 participants in the summer season 2024. Milestone 1.1 (M1.1) represents the successful completion of the first expedition. In 2025, the second expedition with another 140 participants will take place (M1.2). Data analyses will be conducted in the following months and manuscript submission can be expected thereafter (M1.3). In 2026, analysis of secondary outcomes such as pooled blood samples, exercise, cerebrovascular and ventilatory responses will be scheduled (M1.4).

| **01.01.2024 to 31.12.2026 (3 years)** | **2024** | | | | | | | | | | | | **2025** | | | | | | | | | | | | **2026** | | | | | | | | | | | |
|---|---|---|---|---|---|---|---|---|---|---|---|---|---|---|---|---|---|---|---|---|---|---|---|---|---|---|---|---|---|---|---|---|---|---|---|---|
| | J | F | M | A | M | J | J | A | S | O | N | D | J | F | M | A | M | J | J | A | S | O | N | D | J | F | M | A | M | J | J | A | S | O | N | D |
| Preparation | | | | | | | | | | | | | | | | | | | | | | | | | | | | | | | | | | | | |
| Data Collection | | | | **n = 140** | | | | | | M1.1 | | | | | | **n = 140** | | | | | | M1.2 | | | | | | | | | | | | | | |
| Interim Analysis | | | | | | | | | | | | | | | | | | | | | | | | | | | | | | | | | | | | |
| Primary data analysis | | | | | | | | | | | | | | | | | | | | | | | | | | | | M1.3 | | | | | | | | |
| Secondary data analysis | | | | | | | | | | | | | | | | | | | | | | | | | | | | | | | | | | | M1.4 | |
| | J | F | M | A | M | J | J | A | S | O | N | D | J | F | M | A | M | J | J | A | S | O | N | D | J | F | M | A | M | J | J | A | S | O | N | D |
| | **2024** | | | | | | | | | | | | **2025** | | | | | | | | | | | | **2026** | | | | | | | | | | | |

***Figure 5. Schedule and Milestones of the proposed study.*** *Grey areas indicate periods of scheduled work packages, Letters represent months of the years 2024-2026.*

#### 2.5. Relevance and impact

##### 2.5.1. Scientific relevance

Previous research has been conducted to elucidate AMS susceptibility, prevention, and treatments, but females have only occasionally been included and have never been appropriately characterized. No randomized clinical trial has been conducted to investigate and compare the efficacy of acetazolamide against AMS incidence in females compared to men, despite the knowledge that females are more prone to develop AMS compared to men. Differences in drug efficacy or side-effects based on sex-related differences are indicated in the literature and are likely due to lower blood volume and consequently higher acetazolamide plasma concentration, due to differences in the hypoxic chemosensitivity or other factors between females and men. The current gaps in our knowledge are mainly due to the demanding logistical and organisational challenges in conducting large-scale mountain medicine and physiology research, especially in females. The applicant and his partners have acquired the knowledge and study setting to overcome these challenges. Therefore, this study will provide conclusive results on the efficacy and side-effect of preventive acetazolamide against AMS in females compared to males and whether AMS susceptibility depends on female sex hormones or MCPs. Moreover, physiological, urine and blood analyses will provide comprehensive insights into physiological and clinical differences between females and males under hypoxic conditions. Findings from this large-scale project are expected to become milestones in the research history of high altitude medicine, opening up new avenues for mechanistic research related to sex- and hormone-dependent tolerance to hypoxia.

##### 2.5.2. Broader Impact

Females are substantially underrepresented in heart failure, acute coronary syndrome and pulmonary disease studies, with females <55 years accounting for <10% of the study population and in case of an incident females present with a worse outcome.[30,134,135] The literature shows that many lung diseases are more commonly found in females and present with higher degree of severity, exacerbation rate, hospitalizations and mortality than in men.[136] In this regard, insights from this altitude study investigating hypoxia- and hypoxemia-related physiological and clinical adaptations in females compared to males might enhance our understanding of the known sex-related differences in lung diseases at low altitude. Furthermore, to date, there is limited performance of hormonal and cell immunological diagnostic endeavour and low inclusion of female participants in clinical trials on AMS prevention. Our findings are expected to have a large and broad impact, since a considerable number of summer and estimated 400 million annual winter visits will eventually be affected by altitude-related health impairments.[5,137] Based on the expected results, female athletes might be able to improve their high altitude performance or live-high, train-low training schedules based on the MCP. Exercise training based on the MCP is already common in top athletes, therefore, sports in the mountains are equally affected. Understanding sex differences in altitude tolerance may finally result in less accidents and emergency evacuations, as well as improved work efficiency in employees temporarily working at high altitudes. Moreover, translational knowledge transfer of these findings from hypoxemic conditions might contribute to a better understanding of acute cardiopulmonary diseases associated with hypoxemia (i.e. COVID), their progression, their therapy, and any related sex differences as well as to millions of highlanders living at altitudes >2500 m, who are chronically hypoxemic.

10.1378/chest.14-0495

112. Richalet JP, Larmignat P, Poitrine E, Letournel M, Canoui-Poitrine F. Physiological risk factors for severe high-altitude illness: a prospective cohort study. *Am J Respir Crit Care Med*. Jan 15 2012;185(2):192-8. doi:10.1164/rccm.201108-1396OC
113. Siebenmann C, Robach P, Lundby C. Regulation of blood volume in lowlanders exposed to high altitude. *J Appl Physiol (1985)*. Oct 1 2017;123(4):957-966. doi:10.1152/japplphysiol.00118.2017
114. Schlittler M, Gatterer H, Turner R, et al. Regulation of plasma volume in male lowlanders during 4 days of exposure to hypobaric hypoxia equivalent to 3500 m altitude. *J Physiol*. Feb 2021;599(4):1083-1096. doi:10.1113/jp280601
115. Roche J, Rasmussen P, Gatterer H, et al. Hypoxia briefly increases diuresis but reduces plasma volume by fluid redistribution in women. *Am J Physiol Heart Circ Physiol*. Dec 1 2022;323(6):H1068-h1079. doi:10.1152/ajpheart.00394.2022
116. Swenson ER. Pharmacology of acute mountain sickness: old drugs and newer thinking. *Journal of Applied Physiology*. 2016;120(2):204-215. doi:10.1152/japplphysiol.00443.2015
117. Siebenmann C, Keiser S, Robach P, Lundby C. CORP: The assessment of total hemoglobin mass by carbon monoxide rebreathing. *J Appl Physiol (1985)*. Sep 1 2017;123(3):645-654. doi:10.1152/japplphysiol.00185.2017
118. Champigneulle B, Stauffer E, Robach P, et al. Early effects of acetazolamide on hemoglobin mass and plasma volume in chronic mountain sickness at 5100 m. *Pulmonology*. May 30 2023;doi:10.1016/j.pulmoe.2023.05.006
119. Stembridge M, Williams AM, Gasho C, et al. The overlooked significance of plasma volume for successful adaptation to high altitude in Sherpa and Andean natives. *Proc Natl Acad Sci U S A*. Aug 13 2019;116(33):16177-16179. doi:10.1073/pnas.1909002116
120. Baskurt OK, Boynard M, Cokelet GC, et al. New guidelines for hemorheological laboratory techniques. *Clinical hemorheology and microcirculation*. 2009;42(2):75-97. doi:10.3233/ch-2009-1202
121. Mazzarino M, Di Costanzo L, Comunità F, Stacchini C, de la Torre X, Botrè F. UHPLC–HRMS Method for the Simultaneous Screening of 235 Drugs in Capillary Blood for Doping Control Purpose: Comparative Evaluation of Volumetric and Non-volumetric Dried Blood Spotting Devices. *ACS Omega*. 2022/09/13 2022;7(36):31845-31868. doi:10.1021/acsomega.2c01417
122. Ross RM. ATS/ACCP statement on cardiopulmonary exercise testing. *Am J Respir Crit Care Med*. May 15 2003;167(10):1451; author reply 1451. doi:10.1164/ajrccm.167.10.950
123. Borg G. *Borg's perceived exertion and pain scales*. Human kinetics; 1998.
124. Sooronbaev T, Osmonov B, Furian M, et al. Time course of pulmonary artery pressure and clinical characteristics in Kyrgyz highlanders. *Eur Respir J*. 2015;46(suppl 59):PA3761. doi:10.1183/13993003.congress-2015.PA3761 %J European Respiratory Journal
125. Nachman D, Gilan A, Goldstein N, et al. Twenty-Four-Hour Ambulatory Blood Pressure Measurement Using a Novel Noninvasive, Cuffless, Wireless Device. *Am J Hypertens*. Nov 20 2021;34(11):1171-1180. doi:10.1093/ajh/hpab095
126. Oddo M, Sandroni C, Citerio G, et al. Quantitative versus standard pupillary light reflex for early prognostication in comatose cardiac arrest patients: an international prospective multicenter double-blinded study. *Intensive Care Med*. Dec 2018;44(12):2102-2111. doi:10.1007/s00134-018-5448-6
127. Ong C, Hutch M, Smirnakis S. The Effect of Ambient Light Conditions on Quantitative Pupillometry. *Neurocrit Care*. Apr 2019;30(2):316-321. doi:10.1007/s12028-018-0607-8
128. Shaballout N, Aloumar A, Manuel J, May M, Beissner F. Lateralization and Bodily Patterns of Segmental Signs and Spontaneous Pain in Acute Visceral Disease: Observational Study. *Journal of medical Internet research*. Aug 27 2021;23(8):e27247. doi:10.2196/27247
129. Saghaei M. An overview of randomization and minimization programs for randomized clinical trials. *J Med Signals Sens*. Jan 2011;1(1):55-61.
130. Gamble C, Krishan A, Stocken D, et al. Guidelines for the Content of Statistical Analysis Plans in Clinical Trials. *JAMA*. 2017;318(23):2337-2343. doi:10.1001/jama.2017.18556
131. Schulz KF, Altman DG, Moher D, Group C. CONSORT 2010 statement: updated guidelines for reporting parallel group randomised trials. *BMJ*. Mar 23 2010;340:c332. doi:10.1136/bmj.c332

### SEX-SPECIFIC EFFICACY AND SAFETY OF PREVENTIVE ACETAZOLAMIDE FOR ACUTE MOUNTAIN SICKNESS IN HEALTHY LOWLANDERS: A RANDOMISED, DOUBLE-BLIND, PLACEBO-CONTROLLED TRIAL.

**Principal Investigator**

- Dr. sc. ETH Michael Furian, University Hospital Zurich, Pulmonology Department, Zurich, Switzerland

**Swiss Project Partners**

- Prof. Dr. med. Silvia Ulrich, MD, Dept. of Respiratory Medicine, University Hospital Zurich, Zurich, Switzerland
- Prof. Dr. med. Konrad Bloch, MD, Dept. of Respiratory Medicine, University Hospital Zurich, Zurich, Switzerland
- PD Dr. med. Cornelia Betschart, MD, Gynaecology Department, University Hospital Zurich, Switzerland

**International Project Partners**

- Prof. Dr. med. Talant Sooronbaev, MD, Dept. of Respiratory Medicine, National Center of Cardiology and Internal Medicine, Bishkek, Kyrgyzstan
- Dr. sc. hum, MSc. med. Biometry & Statistics Nicola Benjamin, Center for pulmonary hypertension, University Clinic Heidelberg, Germany
- Research Consortium of EXALT – Centre d'Expertise sur l'Altitude, France:
  Prof. Dr. sc. Samuel Vergès (Université de Grenoble), Prof. Dr. Julien Brugniaux (Université de Grenoble), Dr. Benoit Champigneulle (Université de Grenoble), Prof. Dr. Aurélien Pichon (Université de Poitiers), Prof. Dr. Paul Robach (Ecole Nationale de Ski et d'Alpinisme), Dr. Emeric Stauffer (Hospices Civils de Lyon)
- Prof. Dr. Andrea Olschewski, Anaesthesiology and Intensive Care Medicine, Medical University of Graz, Austria, Prof. Dr. Horst Olschewski, Medical Faculty, Sigmund Freud Private University, Vienna, Austria and Charité University Medicine, Berlin, Germany.

**Contact**

Dr. sc. ETH Michael Furian
University Hospital Zurich
Pulmonology Department
Raemistrasse 100
CH-8091 Zurich, Switzerland


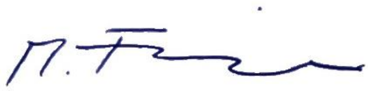

03.02.2025, Version 1.2

### 1. Summary of the research plan

**Background and rationale:** Millions of people travel to high altitude for work or leisure activities and are exposed to reduced inspiratory oxygen partial pressure and hypoxemia that may lead to altitude illness, among which the most common form is acute mountain sickness (AMS). The main AMS symptoms are headache, malaise, weakness, and fatigue. Prospective studies have shown that 20–60% of newcomers at 2500–4000m develop AMS requiring them to take medications, while, at very high altitudes, AMS may progress to high altitude cerebral oedema. Whether females are more susceptible to AMS remains insufficiently understood since no prospective study controlled for sex hormones, use of hormone contraception or assessed menstrual cycle phase (MCP) at altitude. Therefore, females remain underrepresented and poorly characterized in high altitude studies. In addition, the efficacy and safety of 250 mg/day acetazolamide, the standard recommendation for AMS prevention, has never been compared between sexes, although, females have presumably higher acetazolamide plasma concentration due to lower blood volume. Given the known dose-dependent preventive but also side effects of acetazolamide and equal proportion of females and males among mountain travellers, there is an urgent need to conclusively quantify the efficacy and safety of preventive acetazolamide therapy against AMS in females compared to men.

**Overall objectives and specific aims:** The primary objective is to compare the efficacy of preventive acetazolamide treatment against AMS in females compared to men. Additional objectives are to investigate sex- and MCP-related differences in altitude tolerance including AMS incidence as well as efficacy and safety of acetazolamide compared to placebo.

**Methods:** In this randomized, placebo-controlled, double-blinded parallel trial, healthy, premenopausal females and males aged 18 to 44 years, living <1000m will be recruited. Baseline measurements will take place at the Bishkek University Hospital (760m), Kyrgyz Republic; altitude measurements after ascending within 6h by car to 3600m and during a subsequent stay for 2 days and nights. The primary outcome will be the absolute difference in the acetazolamide treatment effect to reduce AMS incidence in females compared to males during the 2-day stay at 3600m. AMS will be defined as a Lake Louise questionnaire score of ≥3 including headache. Secondary outcomes will be derived from respiratory sleep studies, exercise testing, cerebrovascular reactivity assessments and urine and blood drawings.

Starting the day before ascent and daily in the following 30 days, females will perform self-measurements of morning urine concentrations of estrone-1-glucuronide, pregnanediol-3-alpha-glucuronide and luteinizing hormone. From these, the Cycle Days and MCP of participants while at 3600m will be derived. The primary analysis will be on the intention-to-treat population using the Cochran Mantel-Haenszel test. Secondary outcomes and the influence of MCP will be analysed by mixed linear or logistical regression models. To detect a clinically relevant absolute 20% efficacy difference of acetazolamide between sexes, using a ratio of 2:1, a power of 90%, two-sided alpha level of 0.05 and accounting for dropouts, a total of 180 females and 90 males are required.

**Expected results:** This project will conclusively assess the efficacy and safety of preventive acetazolamide in premenopausal females compared to males at a representative altitude for many touristic destinations worldwide. Additionally, this study will provide unique insights into clinical and physiological sex- and MCP-related differences in altitude tolerance.

**Impact for the field**: The detailed characterization of females traveling to altitude and using acetazolamide will represent a milestone in high altitude medicine and physiology and will substantially contribute to our understanding of sex-related altitude tolerance and the effect of acetazolamide. Insights about MCP-dependent susceptibility to hypoxia and AMS symptoms will translate to our physiological and mechanistical understanding of the human body.

### 2. Research Plan

#### 2.1. Current State of Research in the Field

On September 27th 2022, the Swiss National Council promoted research and therapy of specific females' diseases.[1] Consequently, the Swiss Confederation has just now commissioned the SNSF to establish four new National Research Programmes, among them "gender medicine".[2] There are numerous diseases that affect females exclusively or in the majority, however, the Committee for Social Security and Health of the National Council writes that research into treatment options is lagging. It is therefore essential that females-specific diseases are identified as such and researched more broadly, like through research programs of the Swiss National Science Foundation (SNF). In addition, guidelines for diagnosis, indication and therapy are needed, as well as a clear definition for quality outcome measurements in females. Too little is known about females' diseases, they remain undiagnosed for a long time and cause unnecessary suffering. As indicated in the literature, acute mountain sickness (AMS) might be one of the diseases affecting females more than males.

##### 2.1.1. Acute altitude-related illnesses in healthy individuals

Mountain tourism accounts for 15% to 20% of the annual global tourism revenue (approximately 296 billion US dollars in 2019),[3] highlighting the popularity of trips to mountainous regions. However, mountain travel exposes the human body to lower barometric pressures and reduced arterial blood oxygenation (hypoxaemia), which requires numerous physiological adaptations to protect the body and organs against hypoxaemia-related dysfunction and damage. However, moderate hypoxaemia can trigger the development of acute mountain sickness (AMS)[4] and other conditions that compromise a stay at altitude, e.g., poor sleep quality and exercise intolerance. AMS is the most important acute altitude-related illness, affecting 20-60% of unacclimatized lowlanders staying overnight at an altitude between 2500 and 4000 m.[5] The main symptoms of AMS that may emerge within 6 hours to 1-2 days after ascent to high altitudes are headache accompanied by malaise, weakness, and fatigue, which often resolves after 48 hours at altitude.[4] However, AMS can force people to take medications or prematurely terminate their stay at altitude. At altitudes >4000 m, AMS might become life-threatening by progressing to high altitude cerebral oedema.[4] The growing number of publications including in the highest-ranked medical journals (i.e. NEJM), emphasize the importance of understanding AMS and other medical conditions at high altitude.[6-8] Despite the growing work, the pathophysiology of AMS remains elusive and consequently, the diagnosis still relies on a pattern of subjective symptoms rather than on objective findings.[9] AMS may develop due to inadequate physiological responses to hypoxaemia such as decreased hypoxic ventilatory response,[10,11] increased sympathoadrenal activity, and insufficient early hypoxic diuresis.[12,13] A history of migraine, a rapid ascent, and previous episodes of AMS are the most robust predictors of AMS. In 2018, analysis of the AMS literature and based on the early onset of AMS symptoms, an international consensus committee composed of 83 international mountain medicine experts concluded that disturbed sleep might be a direct consequence of hypoxia rather than a symptom of AMS, causing a change in the Lake Louise score (LLS),[14] the most common questionnaire used to diagnose AMS. The revised questionnaire now uses the four items, "headache", "gastrointestinal symptoms", "fatigue/weakness", and "dizziness/light-headedness" to diagnose AMS, each rated with a score from 0 (none) to 3 (severe). In the presence of headache, a total score of ≥3 is required to diagnose AMS.

##### 2.1.2. AMS IN FEMALES

Whether the prevalence of AMS differs between females and males is still being debated and since the modified definition of AMS in 2018, unknown.[15] However, according to the original AMS definition by LLS, a 2019 meta-analysis of 7669 subjects (2639 females) concluded that females have a 1.24-fold risk (95% CI 1.09 to 1.41) of developing AMS at a minimum altitude of 2500 m compared with men.[16] This finding seems logical, since it is known that females do suffer already at low altitude from more headache episodes than men,[17] however, the female physiology under hypoxia remains not at all understood. Important to note, the conclusion of the meta-analysis is debatable since none of the included studies of this meta-analysis assessed the influence of sex hormones (oestrogen, luteinizing hormone, progesterone), menstrual cycle phases (MCP), pre/postmenopausal status or contraceptive use. However, some uncontrolled studies provide important physiological assumptions for sex-related differences in the AMS incidence. Simplified, the sex-related difference in AMS is assumed to be related to sexual hormones and their impact on the physiological response to hypoxia.[16] In accordance with this assumption, progesterone, which is upregulated during the luteal phase, is a well-known respiratory stimulant associated with elevated minute ventilation and decreased $PaCO_2$ during the luteal compared with the follicular phase.[18] Recently, a large study of 336 premenopausal females confirmed these findings and reported lower arterial oxygen saturation ($SaO_2$) and lower hypoxic ventilatory responses in the follicular compared with the early/mid-luteal phase during normobaric hypoxia exposure ($FiO_2$ 0.115, equivalent to 4800 m).[19] These findings are intriguing, since lower $SaO_2$ and lower hypoxic ventilatory response under exercise are risk factors for AMS.[4,19] Another sex-related risk factor for AMS may be low iron stores[20] in females during the follicular phase just after the menstrual bleeding compared to the luteal phase and compared to men.[21,22] Again, no prospective study has investigated the sex-related difference in iron stores on AMS.

##### 2.1.3. PREVENTION OF AMS

Apart from moderate ascent rate and low sleeping altitude, current guidelines recommend acetazolamide, or dexamethasone starting 24 hours before travel to altitude as preventative AMS measures.[23] However, dexamethasone, a glucocorticoid, is mainly recommended for high-risk situations (emergency rescues, military missions) due to its numerous side-effects including hyperglycaemia, immune suppression, and altered mood. Acetazolamide, a carbonic anhydrase inhibitor, is the main recommended and scientifically-proven, pharmacological prophylaxis against AMS.[24] The current recommended dose of 250 mg/day reduces the relative risk of developing AMS by 44%, with a number needed to prevent one case of AMS of 7.[23,24] However, side effects of acetazolamide have been shown to be dose-dependent and include paraesthesia, dysgeusia, polyuria and fatigue.[25] A systematic literature search in PubMed from database inception to September 2023 identified 8 randomized, placebo-controlled, double-blind trials of acetazolamide for prevention of AMS (defined as the primary outcome) using the Lake Louise questionnaire. The studies are summarized in **table 1**. From the 8 identified clinical trials,[7,26-32] only 6 included females.[7,26,27,29,31,32] From the 6 studies including females, only our own trial reported sex-dependent acetazolamide effects on AMS.[7] However, our trial used 375 mg/day instead of 250 mg/Day acetazolamide; was conducted in healthy aged older than 40years and contains a post-hoc analysis, and it remains unclear if AMS incidence is related to the extent of adaptation of tissue perfusion to counteract tissue hypoxia. So, in conclusion, none of the only 8 identified clinical trials aimed to investigate sex-differences in the AMS prevention; appropriately characterized the female population, nor used the 2018 version of the LLS to diagnose AMS. Based on the only available trial from our research group in 349 healthy lowlanders, we observed a clear tendency towards a stronger acetazolamide effect in females compared to males (relative risk reduction of 31% in females compared to 21% in men, **figure 1)**.

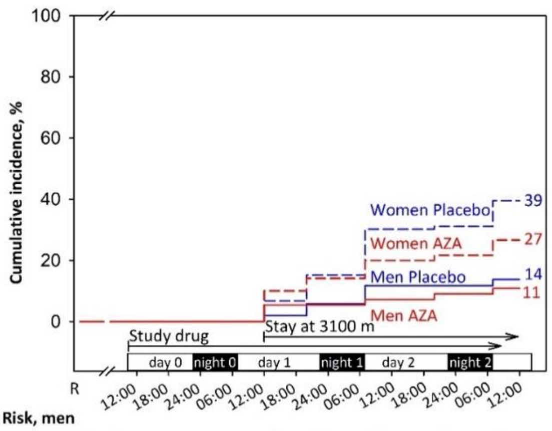

| | | | | | | | |
|---|---|---|---|---|---|---|---|
| **No. at Risk, men** | | | | | | | |
| Placebo | 51 | 51 | 50 | 48 | 45 | 45 | 44 |
| Acetazolamide | 55 | 55 | 52 | 52 | 51 | 50 | 49 |
| **No. at Risk, women** | | | | | | | |
| Placebo | 119 | 119 | 111 | 101 | 83 | 82 | 72 |
| Acetazolamide | 120 | 120 | 108 | 103 | 96 | 94 | 88 |

***Figure 1: Incidence of acute mountain sickness and the effect of preventive 375 mg/day acetazolamide therapy starting 24 hours before and during a 2-day stay at 3100 m in healthy females and males older than 40 years (Furian et al., 2022, NEJM Evidence).*** *This post-hoc analysis showed that females were more prone to acute mountain sickness than men; however, no information regarding menopause, menstrual cycle phases, contraceptive use, or hormone supplementation were assessed. Our study also indicated that preventive acetazolamide therapy before ascending to 3100 m might be of relevance for females, but less for men.*

Whether the acetazolamide efficacy against AMS and its relation to the upregulation of tissue perfusion is in fact sex-dependent has not been studied and represent an important gap of knowledge in the field of altitude medicine and physiology. Physiologically, there are arguments supporting such differences in the efficacy, so, lower blood volume in females compared to males is likely to contribute towards higher acetazolamide plasma concentration when prescribing a standard dose of acetazolamide.[33] Higher plasma concentrations are likely causing a dose-dependent preventive acetazolamide effect on AMS but might also induce more side effects.[25,34] The physiological pathway might be that the higher acetazolamide plasma concentration in females versus males promote a stronger metabolic acidosis resulting in a more pronounced hyperventilation, consequently, better mitigating altitude-induced hypoxia – the main driving factor for AMS. Another sex-related factor influencing the acetazolamide efficacy may be related to the increased hypoxic chemosensitivity in males compared to females. Thus, males are more prone to high altitude periodic breathing,[35] whereas this type of sleep-disordered breathing has been shown to cause superficial sleep and worse subjective sleep quality. Acetazolamide has been shown to be highly effective in preventing altitude-induced sleep-disordered breathing and to improve subjective sleep quality – especially in men.[35] So, males were presumably more likely to rate their sleep as “improved” under acetazolamide when using the original Lake Louise questionnaire including the question related to “impaired sleep”. Consequently, the elimination of the item “impaired sleep” in 2018 is likely reducing the preventive acetazolamide effect against AMS in males whereas in females this change in AMS definition has no impact.

**Table 1. Systematic literature analysis of randomized, placebo-controlled, double-blind trials with primary outcome AMS using the Lake Louise questionnaire (all trials cited in PubMed from database inception to September 2023 are included)**

| Item | Design and setting | Participants | Main findings | Limitations of the study |
|---|---|---|---|---|
| **Females included, analysed separately for acetazolamide effect** | | | | |
| Furian et al.[7] 2022 | Placebo-controlled RCT of 375 mg/day AZA on AMS (LLscore ≥3 including headache).<br>LLS at baseline at 760 m; Altitude at 3100 m for 48 hours. | 349 (69% females) healthy lowlanders aged 40 years or more, residing <800 m. | Females<br>39.0% AMS with placebo; 27.0% AMS with AZA; RR of 30.8%, NNT of 8.3.<br>Men<br>14.0% AMS with placebo; 11.0% AMS with AZA RR of 21.4%, NNT of 33. | • Females not characterized[1]<br>• **375 mg/day instead of 250 mg/day of AZA**<br>• **1993 LLS version for AMS definition[2]**<br>• **Post-hoc analysis of sex-dependent AZA effect in females ≥40 years.** |
| **Females included, not analysed separately or underpowered for detecting sex-related differences** | | | | |
| Lipman et al.[26] 2018 | Placebo-controlled RCT of 250 mg/day AZA or budesonide on AMS (LLscore ≥3 including headache).<br>LLS at baseline at 1240 m; Altitude at 3810 m for 24 hours. | 70 (47.1% females) healthy adult lowlanders residing <1240 m. | 63.0% AMS with placebo; 43.0% AMS with AZA; RR of 32%, NNT of 5. | • **No sex-dependent AZA effect reported**<br>• **Females not characterized[1]**<br>• **1993 LLS version for AMS definition[2]**<br>• **AZA prevention started on day of ascent**<br>• **Assessment <48 hours at target altitude** |
| Van Patot et al.[27] 2008 | Placebo-controlled RCT of 250 mg/day AZA on AMS (LLscore ≥3 including headache).<br>LLS at baseline at 2000 m; Altitude at 4300 m for 24 hours. | 44 (47.7% females) subjects residing <1600 m | 77.3% AMS with placebo; 31.8% AMS with AZA; RR of 58.9%, NNT of 2.2 | • **No sex-dependent AZA effect reported**<br>• **Females not characterized1**<br>• **No sex-dependent AZA effect reported**<br>• **1993 LLS version for AMS definition[2]**<br>• **Assessment <48 hours at target altitude** |
| Basnyat et al.[29] 2003 | Placebo-controlled RCT of 250 mg/day AZA on AMS (LLS headache + another symptom).<br>LLS at baseline at 4243 m; Altitude at 4937 m. | 155 (32.9% females) trekkers. | 24.7% AMS with placebo; 12.2% AMS with AZA; RR of 50.6%, NNT of 8 | • **No sex-dependent AZA effect reported**<br>• **Females not characterized[1]**<br>• **No baseline <1500 m**<br>• **Trekkers already acclimatized**<br>• **No sex-dependent AZA effect reported**<br>• **Modified AMS definition[2]**<br>• **Assessment <48 hours at target altitude** |
| Chow et al.[31] 2005 | Placebo-controlled RCT of 500 mg/day AZA or Ginkgo biloba on AMS (LLscore ≥3 including headache; also in combination with the clinical assessment score).<br>LLS at baseline at 1230 m; Altitude at 3800 m for 24 hours. | 47 (44.7% females) adult lowlanders residing <1200 m. | 60.0% AMS with placebo; 30.0% AMS with AZA; RR of 50%, NNT of 3.3<br>1 woman under placebo developed HAPE. | • **No sex-dependent AZA effect reported**<br>• **Females not characterized[1]**<br>• **No sex-dependent AZA effect reported**<br>• **Modified AMS definition[2]**<br>• **Assessment <48 hours at target altitude** |
| Gertsch et al.[32] 2004 | Placebo-controlled RCT of 500 mg/day AZA or Ginkgo biloba or both on AMS (LLscore ≥3 including headache).<br>LLS at baseline at 4280; Altitude at 4358 m after the first night. | 303 (29.3% females) healthy trekkers. | 34% AMS with placebo; 12% AMS with AZA. RR of 64.7%, NNT of 4.5. | • **No sex-dependent AZA effect reported**<br>• **Females not characterized[1]**<br>• **Dropout rate of 33%**<br>• **No baseline <1500 m**<br>• **Trekkers already acclimatized**<br>• **No sex-dependent AZA effect reported**<br>• **1993 LLS version for AMS definition[2]**<br>• **Assessment <48 hours at target altitude** |
| **No females included** | | | | |
| Hillenbrand et al.[28] 2006 | Placebo-controlled RCT of 250 mg/day AZA on AMS (LLscore ≥3 including headache).<br>LLS at baseline 3440 m; Altitude at 4930 m on arrival. | 400 (0% females) male Nepali porters | 11.1% AMS with placebo; 12.7% AMS with AZA; | • **No females included**<br>• **Dropout rate of 69%**<br>• **No baseline <1500 m**<br>• **Porters already acclimatized**<br>• **1993 LLS version for AMS definition[2]**<br>• **Assessment <48 hours at target altitude** |
| Moraga et al.[30] 2007 | Placebo-controlled RCT of 500 mg/day AZA or Ginkgo on AMS (LLscore ≥3 including headache).<br>LLS at baseline at 0 m; Altitude at 3696 m for 72 hours. | 36 (0% females) healthy adults residing <1000 m. | 56.0% AMS with placebo; 36.0% AMS with AZA; RR of 35.7%, NNT of 5. | • **No females included**<br>• **1993 LLS version for AMS definition[2]** |

[1] Females were not characterized in terms of their reproductive status (pre- / peri- or postmenopausal), use of contraception), hormonal supplementation, or menstrual cycle phase during the stay at altitude. [2]According to the 2018 revised recommendations for defining AMS, a Lake Louise Questionnaire score of at least 3 points including at least mild headache and at least one other symptom of nausea and vomiting, fatigue and/or weakness, dizziness

and/or light-headedness within the first 48 hours at high altitude, is defined as AMS.[15] RR, risk reduction; NNT, number needed to treat; AMS, acute mountain sickness; AZA, acetazolamide; RCT, randomized clinical trial; LLS, Lake Louise Questionnaire.

#### 2.2. CURRENT STATE OF OWN RESEARCH

**The current project partners** have successfully collaborated in various research projects in the field of high-altitude medicine and physiology, including the SNF-funded projects (32003B, 192048, 172980, 143875, 122081, IZK0Z3_168254). They have an outstanding expertise in carrying out major clinical trials in healthy and patients with respiratory disease at high altitude places worldwide. To strengthen their collaboration, they launched the “Swiss-Kyrgyz High Altitude Medicine and Research Initiative” endorsed by the University Hospital and the University of Zurich, the Kyrgyz Ministry of Health and the National Center of Cardiology and Internal Medicine (NCCIM) in Bishkek, Kyrgyz Republic.[51] The partners established several research facilities in Kyrgyzstan offering excellent infrastructure for clinical trials: the National Center of Cardiology and Internal Medicine (Bishkek, 760 m), the Tuja Ashu High Altitude Clinic (Tuja Ashu pass, 3’100 m), the Kumtor Gold Mine Operation Facility (Issyk Kul Oblast, 3600 ), and at the Aksay Health Post (3’200 m).

**Dr. sc. ETH Michael Furian** This proposal consolidates and exploits many years of my own experience and expertise in conducting and leading major international studies and clinical trials in Switzerland, Chile, Peru, France, Ecuador, Antarctica and Kyrgyzstan on high-altitude physiology and medicine. This proposal aims to foster my independency in the field of high-altitude medicine and physiology. Until today, I have investigated AMS,[36,37] sleep-disordered breathing,[38,39] exercise performance,[40-42] cerebrovascular reactivity,[43,44] and the effects of acclimatisation and repeated altitude exposure[45] in healthy lowlanders, patients with chronic obstructive pulmonary disease (COPD), and in highlanders permanently living at high altitude.[43,46] I led several randomised placebo-controlled trials as principal investigator investigating the preventive efficacy of nocturnal oxygen therapy and dexamethasone in patients with COPD staying at altitude.[38,39,44,47,48] In a land-mark trial in 185 patients with COPD and 345 healthy subjects ≥40 years staying for 2 days at 3100 m, we recently showed that prophylactic acetazolamide therapy reduces altitude-related adverse health effects in COPD patients and AMS in healthy individuals.[7] This trial further suggested that females were at increased risk of altitude-related illnesses and that the efficacy of acetazolamide might be sex-dependent.

**Prof. Dr. med. Konrad Bloch (KEB)** KEB has a longstanding experience in performing high altitude field studies including in Switzerland (Capanna Regina Margherita, Jungfraujoch, Davos, St. Moritz) and abroad. This research on effects of altitude on cardio-respiratory function,[49,50] exercise, sleep,[51-54] cognitive performance,[55,56] acclimatization and altitude-related illness in healthy individuals[51,57,58] is internationally well recognized. KEB was among the first to perform randomized, placebo-controlled trials in patients with respiratory conditions going to high altitude. These studies established current treatment recommendations for patients with obstructive sleep apnea travelling to altitude.[59-61] KEB has also performed physiological and clinical studies in COPD patients at low and high altitude.[40,41,62] His team performed the first randomized, placebo-controlled trials evaluating prevention of ARAHE in COPD patients with dexamethasone and AZA.[7,36,63-68]

**Prof. Dr. med. Silvia Ulrich (SU)** is director of the Dept. of Respiratory Medicine, University Hospital of Zurich. She has extensive clinical experience in respiratory and internal medicine and in particular in the invasive and non-invasive diagnosis of pulmonary hypertension (PH), treatment of PH, exercise and training enhancing effects of oxygen in cardiorespiratory

diseases and effects of hypoxia in patients with cardiorespiratory diseases and highlanders[63,68-91]. SU conducted several investigator-initiated studies on the pathogenesis and treatment of PH including several placebo-controlled trials on oxygen therapy in PH-patients,[66-68] exercise hemodynamics and exercise limiting factors in PH,[69,70] and the association of sleep disordered breathing with PH, [66] and acute and subacute effects of acetazolamide on hemodynamics and clinical outcomes in PH patients.[78,89,91,92] SU has performed extensive research in high altitude physiology and medicine with a special focus on the pulmonary circulation at rest and during exercise and patients with pulmonary vascular disease going to altitude and extensive collaborative studies with the applicant and collaborators.[69,70,85,86,93-95]

**Prof. Dr. med. Talant Sooronbaev (TS)** is director of the National Center of Cardiology and Internal Medicine (NCCIM) and head of the Dept. of Respiratory Medicine in Bishkek, Kyrgyzstan. He is Chief Pulmonologist in the Ministry of Health of the Kyrgyz Republic and member of the National Medical Research Council. TS directs the research facilities operated by the "Swiss-Kyrgyz High Altitude and Medicine Initiative". He performed various studies on cardio-respiratory disease in Kyrgyz highlanders[96-98] and is a main collaborator in various altitude studies conducted by our project team. TS has also studied the prevalence of COPD in high altitude residents in the Burden of Obstructive Lung Disease study (www.boldstudy.org) and in the FRESH AIR study http://www.theipcrg.org/freshair.

**PD Dr. med. Cornelia Betschart (CB)** is a gynecologist and obstetrician at the University Hospital Zurich since 2008 and board-certified urogynecologist since 2016. She is interested in AMS as a long-standing member of the Swiss Alpine Club. Scientifically she conducts several clinical multicenter observational, and prospective randomised placebo-controlled trials as principal investigator, also granted by the SNF and private foundations. She promotes inter-professional and inter-disciplinary research as founding member of the International Collaboration for Harmonising Outcomes, Research, and Standards in Urogynaecology and Women's Health (CHORUS), as president of the Swiss Urogynecological Association, steering-board member of Gender medicine USZ, co-founder and leader of Pelvic Floor Center USZ and deputy chairwoman of the Department of Gynecology USZ.

**Dr. sc. hum, MSc. med. Biometry & Statistics Nicola Benjamin (NB)** is the Head of study coordination and scientific project management of the Center for pulmonary hypertension at the University Clinic of Heidelberg, Germany. NB has a broad and extensive knowledge on clinical trial design and statistical analyses in the field of Pulmonology (including hypoxia and hypoxemia).[99-101] She contributed to the trial design and statistical considerations of this application, furthermore, NB will supervise the creation of the statistical analysis plan, the interim analysis and the final statistical analyses of this project.

**EXALT Research Consortium** is composed of outstanding high-altitude researchers situated in France. These researchers, namely Prof. Dr. Samuel Vergès (Université Grenoble Alpes), Prof. Dr. Julien Brugniaux (Université Grenoble Alpes), Dr. Benoit Champigneulle (Université Grenoble Alpes), Prof. Dr. Aurélien Pichon (Université de Poitiers), Prof. Dr. Paul Robach (Ecole Nationale de Ski et d'Alpinisme), Dr. Emeric Stauffer (Hospoces Civils de Lyon), have conducted landmark studies in France and in Peru (Expedition 5300 m).[102-106] They are specialized in comprehensive physiological and basic research at altitudes up to 5300 m. Due to SNF Postdoc.Mobility grant in 2020, the main applicant joined this team in Peru, La Rinconada in 2021 and invited them to research studies in Kyrgyzstan.[107,108] They will strongly contribute to the physiological project aims to understand physiological and mechanistic sex-differences in altitude tolerance and mechanisms of action of acetazolamide.

**PD Dr. med. Matthias Hilty** is senior attending physician in the University Hospital Zurich Intensive Care Unit. He has extensive experience in the micro-hemodynamic response to tissue hypoxia, both in high-altitude settings at Jungfraujoch research station and the Himalyas[109] and in critically ill patients presented with circulatory shock.[110] He has developed objective measures of sublingual microcirculatory diffusion and convection capacity in the tissue,[111] and the dynamic response to a nitro-glycerine stimulus,[112] to quantify tissue perfusion and the state of regulation mechanisms in the systemic microcirculation.

##### 2.2.1. Insights from two preceding pilot studies in females

Assessing sex-related differences at high altitude require meticulous planning, translational knowledge in interdisciplinary research fields and established high altitude research facilities. Therefore, to gain important insights into the expected challenges of high-altitude studies in females, and to enhance the feasibility of the proposed project, the main applicant of this proposal conducted two pilot studies in overall 42 healthy premenopausal females in 2022. All females were instructed to monitor their urinary hormone concentrations for 30 consecutive days by the *Full Cycle Hormone Insights Kit* (Proov, MFB Fertility Inc., CO, US) starting the day before traveling to 3100 m or 3600 m and staying there for 2 days/nights. These pilot studies revealed excellent hormone monitoring adherence, with 1201 of 1230 (97.6%) successful hormone analyses, providing a robust and reliable hormone measurement technique. The averaged and smoothed hormone profile illustrated in **Figure 2, Panel A** shows that ascending to high altitude, independent of the MCP, results in the expected random distribution of participants throughout the cycle days (**Figure 2, Panel B**). This important finding confirms that any large-scale randomized clinical trial does not require to schedule altitude ascents based on the Cycle Day or MCP. The equal distribution of the participants throughout the Cycle Days are a clear strength of the study design, since it avoids any biased conclusion based on comparisons of a few isolated Cycle Days in the luteal compared to follicular MCP (in case randomization is based on specific Cycle Days or MCP).

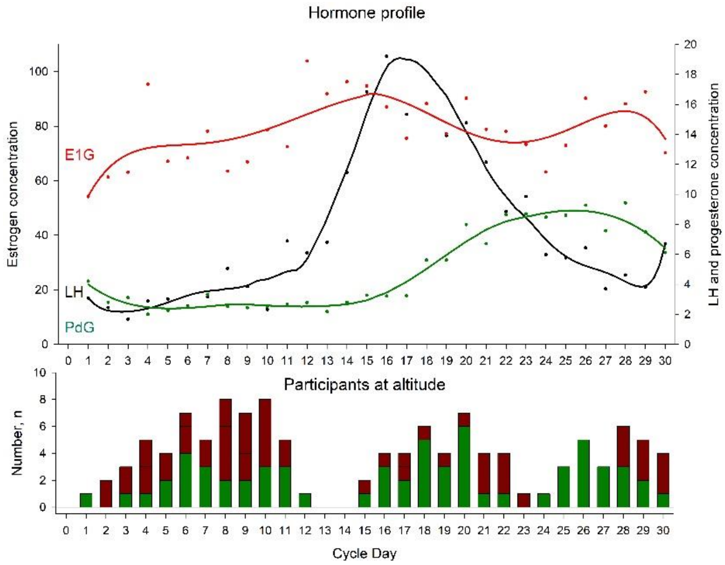

***Figure 2: Urinary hormone profile from 42 females travelling to high altitudes. Panel A:*** *Females daily monitored their urinary hormones E1G, LH and PdG for 30 consecutive days. Based on their first day of the menstruation, the individual Cycle Day at altitude ascent was calculated. Hormone concentrations were averaged for each Cycle Day and a curve was fitted for each hormone.*

***Panel B:*** *The total number of participants staying at altitude and in relation to their individual Cycle Day. The green proportion of the bars represent females without AMS; red proportion of the bars represent females with diagnosed AMS.*

In addition, when comparing the findings at 3100 to 3600 m (**Table 2**), the pilot studies provide important quantitative data related to AMS incidences. Therefore, an AMS incidence of 57% can be expected in premenopausal females staying for 2 days/nights at 3600 m.

**Table 2. Main findings of the two pilot studies.**

| | Group 1 (n = 21) | | | Group 2 (n = 21) | | | Mean difference in altitude effect (3600 m vs 3100 m) (95% CI) |
|---|---|---|---|---|---|---|---|
| | 760 m | 3100 m | Mean altitude effect (95% CI) | 760 m | 3600 m | Mean altitude effect (95% CI) | |
| **Clinical examination** | | | | | | | |
| Systolic BP, mmHg | 106 ± 2 | 106 ± 2 | 0 (-3 to 3) | 104 ± 2 | 105 ± 2 | 1 (-2 to 5) | 1 (-3 to 6) |
| Diastolic BP, mmHg | 73 ± 2 | 74 ± 2 | 1 (-3 to 5) | 70 ± 2 | 76 ± 2 | 6 (2 to 10)* | 5 (0 to 10) |
| Heart rate, bpm | 85 ± 2 | 87 ± 2 | 2 (-3 to 7) | 78 ± 2 | 95 ± 2 | 17 (12 to 22)* | 15 (8 to 23)* |
| $SpO_2$, % | 96.1 ± 0.4 | 93.0 ± 0.4 | -3.1 (-4.2 to -2.1)* | 97.3 ± 0.4 | 86.8 ± 0.4 | -10.5 (-11.6 to -9.4)* | -7.3 (-8.8 to -5.8)* |
| Acute mountain sickness | | | | | | | |
| 2018 Lake Louise Score | 0.0 ± 0.3 | 0.3 ± 0.3 | 0.3 (-0.4 to 1.0) | 0.0 ± 0.3 | 1.9 ± 0.3 | 1.9 (1.2 to 2.6)* | 1.6 (0.6 to 2.6)* |
| 2018 AMS incidence, % | 0% | 43% | | 0% | 57% | | |

BP, blood pressure; $SpO_2$, arterial oxygen saturation assessed by finger pulse oximetry.

#### 2.3. Detailed research plan

##### 2.3.1. Purpose

The primary purpose of this project is to compare the efficacy of preventive acetazolamide treatment against AMS in females compared to men. An important secondary purpose will be to compare the AMS incidence in females versus males at 3600 m and its relation to the extent of adaptation of tissue perfusion to counteract tissue hypoxia. Additional purposes are to investigate sex- and MCP-related differences in altitude tolerance and efficacy of acetazolamide prevention and to underpin clinical findings with various physiological measures providing important mechanistic insights. Findings will improve our understanding of AMS pathophysiology and will provide scientific evidence whether sex-related recommendations on altitude travel and prevention of altitude illness are necessary.

##### 2.3.2. Primary Hypothesis

The primary hypothesis will be that, during the time course of 2 days/nights at 3600 m, preventive 250 mg/day acetazolamide therapy starting 24 hours before ascending, will reduce the AMS incidence significantly more in females compared to men. AMS will be defined as a Lake Louise questionnaire score of ≥3 points including headache.[14]

##### 2.3.3. Additional Hypotheses

Altitude effects in females

1) Females under placebo staying at 3600 m experience altitude-induced exercise intolerance after arrival at 3600 m compared to 760 m assessed by a maximal cardiopulmonary bicycle exercise test.

Altitude effects between females and men

2) Females under placebo have a higher AMS incidence compared to males under placebo during the time course of 2 days and nights at 3600 m.
3) Females under placebo have less sleep-disordered breathing during the first night at 3600 m compared to males under placebo assessed by respiratory polygraphy.

Acetazolamide effects in females

4) Acetazolamide therapy mitigates altitude-induced exercise intolerance compared to placebo.

Acetazolamide effects between females and men

5) The acetazolamide plasma concentration in the morning after the first night at 3600 m is higher in females compared to men.
6) Perceived acetazolamide-related side effects are higher in females compared to males during the time course of 2 days and nights at 3600 m.

MCP-related differences

7) In females under placebo staying at 3600 m during the luteal MCP, the AMS incidence; altitude-induced exercise intolerance and cardiac repolarization disturbances are lower compared to females during the follicular MCP.

##### 2.3.4. Study design and setting

This is a randomized, placebo-controlled, double-blind, parallel trial (**Figure 3**) conducted over 2 consecutive summer seasons. Healthy females and males (according to physician assigned sex at birth) will perform measurements at the National Center of Cardiology and Internal Medicine, Department of Respiratory Medicine, Bishkek (760 m), Kyrgyzstan and in the Kumtor High Altitude Facility (3600 m), Kyrgyzstan. After written consent of female participants, they will be randomized to either acetazolamide or placebo and will start the sexual hormone monitoring in the morning urine as described in the **section 2.3.6** below. Baseline measurements at 760 m will be conducted 24 hours before ascending to 3600 m and altitude measurements will be performed while staying for 2 days and nights at 3600 m. Male participants will undergo the same measurements at 760 and 3600 m but will not monitor hormones. Transfers between locations will be performed by minibus within 6 hours.

##### 2.3.5. Participants

Inclusion criteria for female participants are premenopausal, eumenorrheic, non-smoking, healthy females with a BMI >18 kg/m$^2$ and <30 kg/m$^2$, aged 18 to 44 years, and who live at altitudes <1000 m. Exclusion criteria are any pre-existing diseases, regular intake of medication (including oral contraceptives), other types of contraceptives (hormonal intrauterine device, vaginal ring, subcutaneous injections or implants, among others), pregnancy or nursing, anaemic (haemoglobin concentration <10g/dl), and any altitude trip <4 weeks before the study. Additionally, all included females require a mobile phone compatible with the hormone monitoring app *proov*. Male participants fulfilling the above-mentioned, male-applicable inclusion and exclusion criteria will be recruited. Participants will be recruited by several

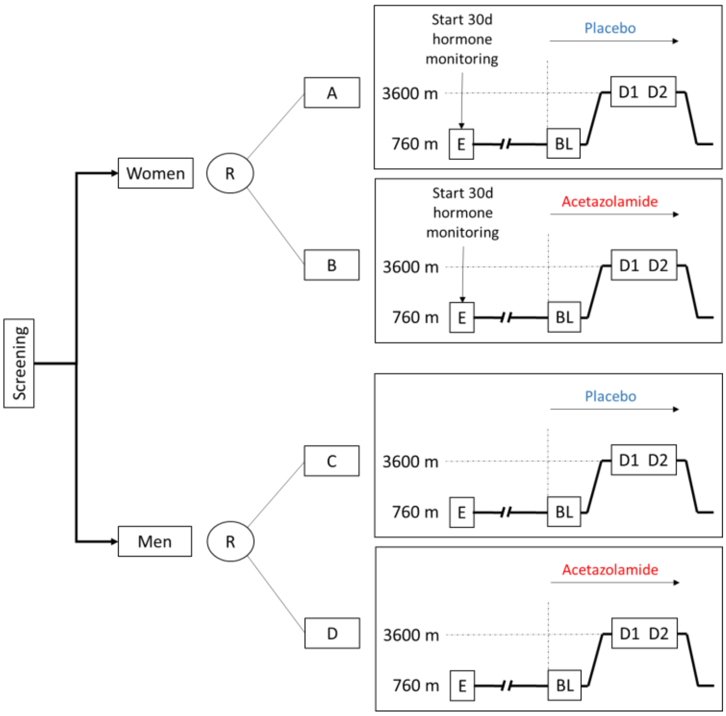

***Figure 3: Study design.*** *Preventive 250 mg/day acetazolamide will be administered 24 hours before travelling and staying for 2 days/ nights at 3600 m. AMS incidence will be assessed at 3600 m. Female participants will monitor urinary levels of sexual hormones for 30 days allowing to determine the MCP during the stay at 3600 m. E, study entry; BL, baseline; R, randomization; D1 – 2, day 1 to 2.*

teams advertising the study at various universities in Bishkek and in the surrounding villages. This recruitment procedure has been applied in several previous randomized clinical trials and has been proven to be extremely successful. The protocol will be submitted to the Ethic Committee of the National Center of Cardiology and Internal Medicine, Bishkek, Kyrgyzstan and to the Cantonal Ethics Committee of Zurich, Zurich Switzerland. Participants will be asked to provide written informed consent.

##### 2.3.6. Measurements during study visits

The schedule for the measurements is illustrated in **Table 3**.

| Table 3. Assessment schedule | | | | | | |
|---|---|---|---|---|---|---|
| | Screening | BL, Day 1, 760 m | BL, Day 2, 760 m | Day of ascent, 760 m / 3600 m | Day 1, 3600 m | Day 2, 3600 m |
| Time before ascent to 3600 m | up to -5 month | -1 week | -1 day | 0 | + 1 day | +2 days |
| Assessment | | | | | | |
| Informed consent (NCCIM) | x | | | | | |
| In-/exclusion criteria (NCCIM) | x | | | | | |
| Reproductive status (NCCIM) | x | | | | | |
| Clinical examination (USZ) | x | x | x | x | x | x |
| AMS questionnaires / Visualize (USZ) | | x | x | x | x | x |
| Respiratory polygraphy (USZ) | | x | | x | x | |
| Iron status (USZ, France) | | x | x | | x | |
| Hypoxic ventilatory response (USZ) | | x | | | | |
| Cerebrovascular reactivity (France) | | x | | | x | |
| Echocardiography (USZ) | | x | | | x | |
| Hemorheology (France) | | x | | | x | |
| OpCo CO-rebreathing (France) | | x | | | x | |
| Funduscopy / Pupillometry (USZ, France) | | x | | | x | |
| Sublingual microcirculation measurement (USZ) | | x | | | x | |
| Forced Oscillation Technique (USZ) | | x | | | x | |
| Acetazolamide concentration (Italy) | | x | | | x | |
| Urinary hormone testing (USZ) | | 30 consecutive days by urine hormone sampling | | | | |

Screening visit

After providing written informed consent, a complete medical history will be obtained. A reproductive status questionnaire will assess regularity of menstrual periods, menstrual cycle length, menarche, pregnancy and breastfeeding, use of contraception, use of other hormone supplementation and known gynecologic or other medical conditions. Inclusion and exclusion criteria will be applied. Physical examination will include weight, height, blood pressure, heart rate, and cardiac and pulmonary auscultation. A pregnancy test will be performed.

History, symptoms, and clinical examination

Repeated physical examinations will include weight, height, blood pressure, heart rate, and cardiac and pulmonary auscultation. AMS will be assessed using the current and the 1993 version of the Lake Louise score[14,113] and the environmental symptoms questionnaire (AMS-c).[114] Subjective sleepiness and sleep quality will be assessed using the Karolinska Sleepiness scale and a 100-mm visual analogue scale. The presence of migraine will be assessed using the ID Migraine Screener, a brief, self-administered screening instrument.[115] A standardised headache diary including information about the date, time, intensity (1 to 10), preceding symptoms, triggers and relief of headache will be distributed. This diary will be incorporated in the 30-day urinary hormone monitoring and will also be distributed to male participants.

Urinary hormone monitoring

Females will monitor the urinary hormone concentrations of estrone-1-glucuronide (E1G), pregnanediol-3-alpha-glucuronide (PdG) and luteinizing hormone (LH) for 30 consecutive days by the FDA approved *Full Cycle Hormone Insights Kit* (proov, MFB Fertility Inc., CO, US) starting before traveling to 3600 m. The *proov* hormone monitoring kit has been used in previous research studies and has been validated against other methods for hormone monitoring.[116] Measurements will be standardized in the morning, after awakening and before drinking or eating breakfast. The *proov* multi-hormone test strip lateral flow assay uses gold nanoparticles and buffered sample pads designed to adjust for pH and hydration levels, filters unwanted particulates and binds contaminants in urine that may interfere with the accuracy of the test. The strip contains three test lines and one control line, corresponding to E1G, PdG and LH (beta subunit). After waiting for 10 minutes, the user provides a photo of her urine test strip with the *proov* app. The application server uses machine learning specifically designed to analyse photographed images of the test strip, control for variations in camera, lightning, and operating system, check for input or output irregularities, and mathematically derive the associated hormone levels.[116] In case of a failure of the analysis due to loss of internet, then an offline analysis by the company is feasibly by providing the time, day and picture of the test strip of the participant. After the completion of the 30-day urine hormone monitoring and in consideration the first day of the last menstrual bleeding, Cycle Days and the participants' MCP will be defined by visually inspecting the 30-day hormone profile. The follicular phase will be defined from the Cycle Day 1 (first day of menstrual bleeding) until the LH peak; the luteal phase will be defined as the day after the LH peak until the day before the next menstrual bleeding.

Hypoxic-ventilatory response test

The hypoxic ventilatory response at exercise (HVRe) has been suggested to be a good predictor for AMS.[19,117] Especially in premenopausal females, it has been suggested that HVRe is higher in the luteal compared to the follicular phase, providing

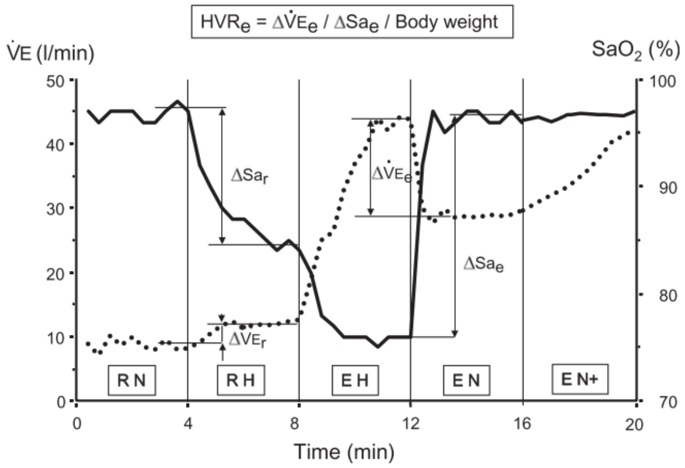

***Figure 4: Hypoxic exercise test.*** *Figure 1 from Richalet et al. 2012.[104] RN, RH, EH, and EN represent rest in normoxia, rest in hypoxia, exercise in hypoxia, and exercise in normoxia, respectively. Hypoxia will be produced by breathing normobaric hypoxic gas (fraction of inspired oxygen of 0.115). Exercise will be performed at 30% of maximal exercise capacity in phases EH and EN. In phase EN+, the exercise intensity will be adjusted so that the heart rate reaches the same value as in EH. HVRe, ventilatory response to hypoxia during exercise; $SaO_2$, arterial oxygen saturation assessed by finger pulse oximetry; $Sa_e$, arterial oxygen saturation during exercise; $Sa_r$, arterial oxygen saturation at rest; V'E, minute ventilation; $V'E_e$, minute ventilation during exercise. Dotted line represents V'E; solid line represents $SaO_2$.*

a potential underlying factor explaining MCP-related differences in AMS susceptibility. Therefore, subjects will perform an exercise test consisting of 5 consecutive phases: 1 – rest in normoxia; 2 – rest in hypoxia (FiO2 of 0.115 equivalent to 4800 m); 3 – exercise in hypoxia (EH) at 30% of maximal normoxic maximal exercise capacity (Ergoselect 200; Ergoline GmbH, Bitz, Germany); 4 – exercise in normoxia (EN) and 5 – EN with the same heart rate achieved during EH (EN+). Arterial oxygenation by finger pulse oximetry and respiratory gas concentrations, tidal volume, and respiratory rate will be measured breath-by-breath by a metabolic unit (Ergostick; Geratherm Medical AG, Gschwenda, Germany) to compute minute ventilation (V'E) and other outcomes. HVRe will be calculated as previously described and illustrated in **figure 4**.

Haemoglobin mass ($Hb_{mass}$) and intravascular volumes

Plasma volume (PV) contraction is an early mechanism allowing an increase in haemoglobin concentration ([Hb]) (and hence, in arterial oxygen content) after an acute high-altitude exposure, and thus, counteracting the decrease in oxygen availability.[118] Experimental studies distinctly conducted in both male and female voluntaries confirmed that this decrease in PV occurred early (*i.e.*, in the first 12-24-hours) and with a similar magnitude between sex, mainly through a fluid-redistribution mechanism from the intra to the extravascular compartment, rather than due to a hypoxic diuresis.[119,120] Beneficial effects of a preventive acetazolamide uptake on AMS are assumed to be mostly driven by the consecutive hyperventilation balancing the induced metabolic acidosis and, thus, increasing the arterial oxygen content; however, even at a such low dose, acetazolamide might have beneficial effect on arterial oxygen content, through its diuretic effect, by further decreasing the PV.[121] To investigate PV changes, an $Hb_{mass}$ measurement using the CO-rebreathing method, allowing the computation of PV, will be conducted at 760 m and after 12 hours staying at 3600 m. Briefly, the CO-rebreathing test will be conducted as previously descripted, using an automated system (OpCo, Detalo Health, Copenhagen, Danmark), after a 20-min rest period in supine position.[122] From a capillary blood sample, [Hb], pre-test percentage of carboxyhaemoglobin (Rapidpoint 500, Siemens AG, Zürich) and haematocrit (Hct, microcentrifuge method) will be determinate in quadruplicate. Next, while the participant will breathe a 100% mixture of oxygen in a rebreathing circuit, a bolus dose of 99.997% pure CO gas (1 mL per kg body mass for men, 0.8 mL per kg body mass for females) will be automatically administered by the Opco system. After a 6-min rebreathing period, the participant will be disconnected from the rebreathing circuit and the residual CO concentration in the circuit will be measured. Then, a secondary capillary blood sampling will be performed at 10 min, to measure, in quadruplicate, the post-test %HbCO (%$HbCO_{POST}$). From the post-pre-test difference in %HbCO and the amount of absorbed CO, the $Hb_{mass}$ value will be calculated and the derived intravascular volumes (PV, red blood cell volume and total blood volume) will be calculated.[122] Previous studies conducted in the field in high-altitude environment have confirmed both the feasibility and the safety of the CO-rebreathing method.[105,123,124]

Hemorheological analysis

Potential effects of a prophylaxis acetazolamide uptake in order to prevent AMS on blood viscosity could be leaded by an expected decrease in PV or by a direct effect on red blood cell (RBC) deformability due to a direct inhibition of RBC carbonic anhydrase,[118] albeit hemorheological evaluations have never been performed in this context. To explore the potential hemorheological effect of acetazolamide prophylaxis administration, and the potential difference of effect between males and females, we will plan to perform blood viscosity and RBC's deformability measurements at 760 m and after the first night at 3600 m. At both altitudes, from a 3 mL venous sample collected in an EDTA tube, the following analyses will be performed, in accordance with the guidelines for hemorheological laboratory techniques:[125]

- Blood viscosity will be measured at native and corrected Hct (40%, after dilution using autologous plasma) at several shear rates (22.5 $s^{-1}$, 45 $s^{-1}$, 90·$s^{-1}$).
- RBC's deformability will be measured by ektacytometry using the Laser Optical Rotational Red Cell Analyzer (LORRCA, RR Mechatronics, Hoorn, The Netherlands) at 3 and 30 Pa and the extent of RBC aggregation and the strength of RBC aggregates will be measured by syllectometry using the LORRCA (RR Mechatronics, Hoorn, The Netherlands) after adjustment of the Hct to 40% with autologous plasma.

Arterial and venous blood sampling

Effects of altitude and acetazolamide on arterial blood gases will be assessed in a radial artery sample after 15 minutes of quiet and awake supine position in the morning after awakening. As previously described, arterial blood will be analysed for pH, $PaO_2$, $PaCO_2$, $SaO_2$, haematocrit and electrolytes (Rapidpoint 500, Siemens AG, Zürich).[38] To quantify and confirm MCPs, absolute hormone concentrations and plasma concentration of acetazolamide and to provide mechanistic explanations to PV changes, a 20 mL peripheral venous blood sample in the morning after awakening will be withdrawn, centrifuged, and the plasma/serum immediately frozen, stored, and transported at -20°C for further analyses. Analysis at 760 m will include iron metabolism (ferritin, TSAT, hepcidin, erythroferrone, sTfR1). At both altitudes, circulating reproductive hormone concentrations (oestradiol, progesterone, LH) , plasma renin activity, aldosterone, copeptin (as a surrogate of the antidiuretic hormone) and midregional proANP (the precursor of the atrial natriuretic peptide) will be analysed. The plasma concentration of acetazolamide will be assessed using dried blood spots.[126] Therefore, 80-100 µl venous blood from the sample will be pipetted on to four spots on the dried blood spot card and analysed as previously described.

ELISA investigations

Serum samples taken at low altitude and high altitude, will be investigated for cytokines, PDGF, VEGF, BNP, and various bone morphogenetic proteins via commercially available ELISA kits.

Liquid chromatography–mass spectrometry metabolomics

Serum samples will be prepared as described earlier (PMID: 40343938). Chromatographic separation will be performed on a Vanquish UHPLC+ system (Thermo Fisher Scientific) equipped with an ACQUITY UPLC BEH Amide column (2.1 × 150 mm, 1.7 µm; Waters), using an 18 min gradient (400 µl/min) from 97% solvent A (ACN/ddH2O, 95/5, v/v; 10 mM NH4FA, 10 mM NH3) to 65% solvent B (ddH2O/ACN, 95/5, v/v; 20 mM NH4FA, 20 mM NH3). Metabolites will be identified either level 1 via accurate m/z of the [M-H]- ion (<5ppm) and comparison of the retention time (rt) and MS2 spectra to synthetical reference compounds or level 2 without reference rt. All peaks (QC) will be manually inspected in Freestyle (1.8 SP2) and peak extraction will be performed in Skyline (24.1.0.199). Only metabolites with <10% peak area variation in QC samples will be used for further processing. Peak areas will be blank subtracted and normalized to IS (rt-range clustered) and protein concentration. Metabolite data will be expressed as AU (analyte-IS ratio)/mg or µg protein.

High-throughput sequencing

DNA extracted from peripheral blood will undergo whole-genome sequencing using the Illumina TruSeq DNA PCR-Free Sample Preparation kit (Illumina Inc., San Diego, CA, USA) and Illumina HiSeq 2500 or HiSeq X sequencer, generating 100–150 bp reads with a minimum coverage of 15× for ~95% of the genome (mean coverage of 35×) (PMID: 29650961). The final libraries will be checked using the Roche LightCycler 480 II (Roche Diagnostics Corporation, Indianapolis, IN, USA) with KAPA Library Quantification Kit (Kapa Biosystems Inc., Wilmington, MA, USA) for concentration.

Cardiopulmonary exercise testing and electrocardiogram

As previously performed by our project team, a resting 12-lead ECG will be performed after resting for 10 minutes in a supine, awake position which will give insights into cardiac repolarization disturbances at 3600 m.[70] Thereafter, a bicycle exercise test with a progressive ramp protocol will be performed until exhaustion (Ergoselect 200, Ergoline GmbH, Bitz, Germany) as previously performed in patients with COPD and healthy;[40,67] and according to international guidelines to assess alterations in physical exercise performance at altitude and under acetazolamide.[42,127] Participants will rest on the bicycle for 10 minutes, followed by a 10 – 20 W/min work-load increase starting at 20 W. Respiratory gas concentrations, tidal volume, and respiratory rate will be measured breath-by-breath (Ergostick, Geratherm Medical AG, Gschwenda, Germany). Arterial oxygenation (by finger oximetry) and a 12-lead ECG will be applied. Before and at peak exercise, dyspnoea sensation and leg fatigue will be assessed with the BORG CR10 scale.[128]

Cardiorespiratory sleep studies

Continuous nocturnal measurements by portable devices (Alice PDX, Philips Respironics, Zofingen, Switzerland) will include nasal pressure swings, chest wall excursion, ECG, and pulse oximetry. Mean oxygen saturation and percentage time with $SpO_2$ <90% and <80% will be computed. The apnoea/hypopnea index and oxygen desaturation index ($SpO_2$ dips >3%) will be computed as mean number of events/h.[52] These measurements will give valuable insights into sleep-disordered breathing often seen at altitude and the effect of acetazolamide. This measurement has been successfully implemented by our project team in patients with COPD and healthy lowlanders and highlanders.[7,129] Additionally, actigraphy will be used to measure physical activity levels in participants suffering from AMS vs. participants without AMS.

Cerebrovascular reactivity assessment

Participants will undergo transcranial near-infrared spectroscopy (NIRS) measurements of the prefrontal cortex, finger pulse oximetry, transcranial Doppler ultrasound (TCD), electrocardiography (ECG), measurement of continuous blood pressure, end-expiratory $PCO_2$ measurement and breathing pattern while the participant is sitting in a comfortable position for 10 minutes breathing room air. After baseline measurements, the participant will be asked to perform squat jumps, hyperventilate and reduce their expired $PCO_2$ by 10 mmHg or do an isometric exercise by pressing a predefined load in the dominant hand by a hand grip dynamometer for 1 minute, in random order. The TCD probe will be used to insonate the middle cerebral artery on the same side as the hand grip maneuver is performed to eliminate neurovascular activity contamination. Between interventions a 5-minute wash-out phase will be implemented to avoid any carry-over effect. The last 2 minutes of quiet breathing room air will be used as baseline measurement and will be compared with hyperventilation in participants under placebo compared to participants under acetazolamide. Furthermore, blood flow response in the middle cerebral artery due to blood pressure alterations will be compared between altitudes, drug and sex.

To complement TCD's assessment of regional cerebral blood flow (CBF) velocity through the middle cerebral artery, global CBF will be assessed during the same procedures using Duplex ultrasound of both the right internal carotid and the right vertebral arteries (ICA and VA, respectively). Combined diameter and blood velocity measurements provide the necessary data to calculate blood flow through these vessels. Owing the ICA and VA are bilateral, the calculated blood flow will then be multiplied by a factor of 2 to obtain a value for total CBF.

Biobeat Wrist Monitor

The non-invasive Biobeat wrist monitor continuously monitors vital parameters including heart rate, breathing frequency, arterial oxygenation and systolic and diastolic blood pressure. The patented algorithm is based on artificial intelligence and machine-learning and has been used in other scientific settings.[130] A unique feature is that the vital parameters are not displayed on the Wrist Monitor, instead, are uploaded to the cloud and are live accessible by the physician through a remote patient monitoring system. This approach minimizes confounding the participant and allows high-quality remote monitoring of the participants. Moreover, vital parameters will be continuously recorded and might allow in the future to use an early warning system to prevent AMS. During this study, each participant will wear a biobeat Wrist Monitor during the complete duration of the altitude sojourn. The continuous data recording will be modelled and analysed by partners of the ETH Zurich.

Lung ultrasonography

Using a phased array probe, bilateral lung ultrasound will be performed in a supine from the second to fourth or fifth intercostal space (left and right hemithorax, respectively) down the parasternal, mid-clavicular, anterior axillary and mid-axillary lines, resulting in 28 total windows of interest (left: 12; right: 16). A B line will be defined as one echogenic, continuous, wedge-shaped, signal arising from the uninterrupted pleural interface with a narrow origin in the near field of the image. The number of B-lines will be counted for each lung field and totalled.[76]

Echocardiography

Cardiac morphological and functional parameters will be assessed by standard two-dimensional Doppler echocardiography. Doppler imaging of the tricuspid annulus and the pulmonary outflow tract will be assessed according to standard methods. This will allow us to assess the following parameters: systolic pulmonary artery pressure ($RV_{sys}$, calculated from the tricuspid regurgitation velocity by using the modified Bernoulli equation: $RV_{sys}$ = 4x ($v_{max}$) where $v_{max}$ is the maximum of the regurgitation velocity jet measured over the tricuspid valve) and the right ventricular outflow tract acceleration time; the right ventricular fractional area change [%] and the tricuspid annular plain systolic excursion [mm]. The fractional area change of the right ventricle will be determined. The tricuspid annular plane systolic excursion (TAPSE) will be measured in M-mode. The stroke volume will be measured based on the left ventricular outflow tract diameter and the velocity time integral over the aortic valve in the apical 5- chamber view or the apical long axis view.

Optic nerve sheath diameter (ONSD)

ONSD measurement will be done with a 10 Mhz ultrasound probe using a portable ultrasound machine (uSmart 3300, Terason, USA). Briefly, a thick layer of gel will be applied on the upper closed eyelid before the transducer will be positioned on it, avoiding any excessive pressure, on the temporal side of the eye. Positioning the transducer in the transverse axis, a first measurement will be done in triplicate; then, the transducer will be positioned in the longitudinal axis, to perform three other measurements. Measurements will be done 3 mm behind the ocular globe. The average value obtained for each eye from the six measured values (three in the transversal and three in the longitudinal axis) will be used for the further calculations. To reduce the measurement time, the ultrasound motions will be captured, and the measurements will be conducted in a second time. Duration of the measurement is estimated to one minute by eye.

Automated Pupillometry (AP)

Quantitative pupillometry will be performed using the NeurOptics® pupillometer (NeurOptics, Irvine, CA, USA) on both eyes (right, then left), following the ONSD measurement. The NeuOptics® pupillometer delivers a standardized light stimulation of fixed intensity (1000 Lux), allowing a rapid and precise measurement (0.05 mm limit) of the pupil size changes.[131] The following pupil parameters, measured by the NeurOptics® pupillometer will be recorded : pupil size (mm) before and after light stimulation, percentage of constriction (%), latency of constriction (s), constriction velocity (mm·s-1) and dilatation velocity (mm·s-1) and NPi (neurological pupil index, a computed parameter provides by the device and based on the previously cited parameters). A special attention will be paid to obtain standardized ambient light conditions (i.e., dark conditions) between the two places to avoid a potential confounder between the LA and the HA measurements, as ambient light level has been shown to impact pupil parameters, even in healthy subjects, excepted for latency time.[132]

Fundoscopy

We conduct fundus photography based on handheld retinal imaging system (VistaView™ by Volk optics) in healthy lowlanders ascending to 3600 meters above sea-level. We hope to gain insight in the feasibility and usefulness of retinal imaging for detection of papilledema as an early sign of increased intracerebral pressure. To ensure optimal recording conditions, mydriasis will be induced by topical application of atropine drops.

Pain visualization with SMaRT app

Pain sensation of AMS and any other pain-related symptoms will be recorded using the Sensation Mapping and Reporting Tool (SMaRT) app on a tablet.[133] This app allows the participants to describe their sensations by sketching it inside the image of a body. This will help to assess the intensity, characterization and location of sensations. Whenever the participants fill out the Lake Louis questionnaire, they will also use the SMaRT app to assess their sensations. Recording these additional symptoms or sensations will help to give a more holistic view of AMS in the participants.

Traditional Chinese Medicine (TCM) tongue diagnosis

The TCM syndrome diagnostics are of interest in the context of AMS, given that the main symptom of AMS is headache. Acupuncture is known to be a non-pharmacological treatment option for headache and TCM syndromes are mainly based on the investigation of the tongue's shape, colour and coating. However, before an interventional study with acupuncture can be planned, more information on the effect of hypoxia and AMS on the tongue appearance is required. In this study, we therefore take pictures of the participant's tongue at baseline and at high altitude. These pictures will then be analysed using machine learning algorithms analyzing the pictures in relation to hypoxia and AMS.

Measurement of the sublingual microcirculation and nitro-glycerine response

Non-invasive high resolution handheld vital microscopy (HVM) image sequences of the sublingual microcirculation are obtained using the incident dark field technology described in detail elsewhere[134] with a CytoCam hand-held microscope (Braedius Medical, Huizen, The Netherlands) connected to a portable computer. The microscope is gently placed on the sublingual mucosa, and three image sequence of four seconds duration are recorded per measurement timepoint and subject, as suggested in the current consensus guidelines set forth by an international task force of the European Society of Intensive Care Medicine.[135] The acetylcholine response is assessed 60 seconds after the topical administration of one drop (50 µl) of 1% ($6.8 \cdot 10^{-2}$ M) acetylcholine solution immediately after reconstitution of acetylcholine lyophilisate (Miochol E,

Bausch & Lomb Swiss, Zug, Switzerland) using distilled water, yielding a dose of 0.5 mg (3.4 µmol) per application. The nitro-glycerine response is assessed 60 seconds after the topical administration of three drops (150 µl) of 1% nitro-glycerine ($4.4 \times 10{-}2$ M, diluted in 1:100 NaCl 0.9%) for a final dose of 0.015 mg per application. Both protocols have previously been demonstrated to avoid impact on systemic blood pressure and leave no measurable traces of nitro-glycerine metabolites in the systemic circulatio.[112] Microcirculatory hemodynamic variables (total vessel density TVD; functional capillary density FCD; proportion of perfused vessels PPV; red blood cell velocity RBCv; capillary hematocrit cHct) and tissue red blood cell perfusion (tRBCp) are calculated from the image sequences via the experimentally and clinically validated MicroTools advances computer vision algorithm as described in detail elsewhere.[111,136] The means of three measurements recorded within the same timepoint and subject are reported.

Forced Oscillation Technique

FOT assesses the mechanical impedance of the respiratory system (Zrs), capturing the resistive and reactive forces required to facilitate an oscillating airflow through the respiratory tract.[137] Therefore, assessing impairments in both central and peripheral airways during pulmonary rehabilitation.[138] FOT measurements will be conducted using a multifrequency signal of 5 Hz from a device meeting European Respiratory Society technical standards (ResMon First).[138] Participants completed FOT assessment in low altitude and first day of high altitude in a seated position while wearing a nose clip and supporting their cheeks to minimize upper airway shunt compliance. The first three breaths were excluded from analysis, and at least 10 artifact-free breaths were automatically selected by the device, ensuring a recording duration exceeding 30 seconds for breathing rates up to 20 breaths per minute, with only full breathing cycles included in the analysis.[139]Outcomes

The primary outcome of this study is the absolute difference in efficacy of acetazolamide to reduce AMS incidence in females compared to men. AMS will be defined as a 2018 Lake Louise questionnaire score of ≥3 points including headache.[15] If a participant scores ≥3 points while staying at 3600 m, the participant will be deemed to have AMS.

Secondary outcomes will be derived from urinary sexual hormone concentrations, AMS severity, AMS symptom components, and AMS defined by different questionnaires and cut-off scores.[113,114] Vital parameters, parameters from arterial and venous blood sampling, hypoxic ventilatory response during exercise, cerebrovascular reactivity and respiratory sleep studies will be obtained and compared between sexes, between acetazolamide and placebo and between MCPs. Additionally, ECG morphology, cardiac repolarization disturbances, sleep-disordered breathing and exercise performance and exercise-limiting factors will be obtained and compared. Mechanistic insights of the prophylactic effect of acetazolamide for AMS will be derived from intravascular volume measurements, $Hb_{mass}$, blood viscosity and hemorheological assessments.

##### 2.3.7. Randomisation, intervention and blinding

Stratified randomization will be conducted for females and males using a computer generated schedule (MinimPy 0.3), stratifying for sex in an initial females-to-males ratio of 2:1, and revised to 1:1 ratio as outlined in the revised sample size estimation during the .[140] Within sex, participants will be 1:1 randomized to A: 250 mg/day acetazolamide (125 mg in the morning with breakfast, 125 mg in the evening with dinner) starting 24 hours before arriving and while staying for 2 days and nights at 3600 m (total of 6 capsules of 125 mg acetazolamide) or B: identically looking, organoleptic placebo capsules. An independent pharmacist will prepare identically looking verum and placebo capsules labelled with secret codes. The list of codes will be kept confidential to investigators and participants until data acquisition and analysis has been completed.

##### 2.3.8. Sample Size Estimation

Sample size estimation was performed using the Cochran Mantel-Haenszel test with continuity correction. The previously described preventive effect of acetazolamide against AMS has been suggested to be 44%.[34] To detect a clinically meaningful efficacy difference of 20% of acetazolamide therapy against AMS in females compared to males (i.e. treatment effect of 50% in females and 30% in men) with a group allocation ratio of 2:1, power of 90%, two-sided alpha level of 0.05, and accounting for dropouts, a total of 180 females and 90 males will be recruited. The females-to-males ratio of 2:1 was chosen to enable conclusive testing of the additional hypotheses within females (see **section 2.3.3**). During the blinded interim analysis outlined in section 2.3.10, which was conducted after the first study year, we found an AMS incidence of 44% in females and 16% in males (in totally 133 randomized participants). The lower-than-expected AMS incidences resulted in a re-sample size estimation. Therefore, a total of 180 females and 120 males will be recruited for this study. The project team has successfully recruited even higher numbers of COPD patients and healthy participants in previous randomized clinical trials at high altitude. As an example, in 2015, 124 COPD patients were recruited within 1 summer expedition and included in the final analysis[36]. In another trial of acetazolamide, 186 COPD patients and 345 healthy subjects were recruited and included in the final analysis within 3 summer expeditions.[7] Since the complex recruitment of these large numbers of chronically ill patients was successful, the anticipated number of healthy females and males required for this study is well feasible and within the capacity of the project team.

##### 2.3.9. Data Analysis and Statistics

In accordance with international guidelines for prospective clinical trials,[141] a statistical analysis plan, similarly to the studies published by the applicant in *NEJM Evidence,*[7] will be created before any statistical analyses. This study will be pre-registered at ClinicalTrials.gov and results reporting will follow the CONsolidated Standards Of Reporting Trials (CONSORT) guidelines.[142] Data will be summarised by numbers and proportions and means (SD) and medians (quartiles) for normally and non-normally distributed data, respectively. Altitude-induced differences will be calculated by mean or median differences (95% confidence intervals). The primary analysis will be performed on the intention-to-treat population including all randomized participants using the Cochran Mantel-Haenszel test comparing the sex-related difference in the acetazolamide effect against AMS.[143] In case of missing values in the primary outcome (AMS, binary), a sensitivity analysis with best / worst case scenario will be applied. Kaplan-Meier curves for AMS will be plotted for the four groups representing the combination of sex and treatment assignment: 1) males randomly assigned to receive acetazolamide; 2) males randomly assigned to receive placebo; 3) females randomly assigned to acetazolamide; 4) females randomly assigned to placebo. Comparisons will be performed using a log-rank test. Secondary, a multivariable Cox proportional-hazards analysis will be conducted to determine whether the interaction between sex and acetazolamide therapy is independent of other baseline factors.

Secondary outcomes will be analysed on the per-protocol population defined as participants with available data. Continuous variables will be analysed using mixed linear regression models, with the variable of interest as the dependent variable and drug, location, sex and MCP as fixed effects, including the interaction term drug*location*sex*MCP.

To proactively minimize any unforeseen deviations from the anticipated study progression, such as extreme benefit or harm of acetazolamide or futility and unexpected AMS incidences, a planned interim analysis will be conducted after the completion of the first study year. The planned interim analysis will be performed by an independent statistician blinded to drug. The statistician will report the blinded results to the project steering committee (composed by the applicant and the co-applicants), who will decide on continuation, termination, or on any appropriate protocol adaptation (e.g., sample size re-estimation). Termination of the study is defined by symmetric stopping boundaries at $p<0.001$ (Peto approach).[144] In the final analysis, a $p<0.05$ will be considered statistically significant.

##### 2.3.10. Further considerations

As in previous studies over the past few years, the applicant has access to a dedicated and experienced Swiss and Kyrgyz research team composed of assistant physicians and medical students, as well as access to pivotal medical devices needed for the described measurements such as ergometers, respiratory polygraphy, computers, and other hardware. This equipment is available from the *Swiss-Kyrgyz High Altitude Medicine and Research Initiative*. Medical devices and consumables will be transported to Kyrgyzstan for the study duration. During data acquisition periods, up to 6 medical bachelors' students from ETH Zürich will perform their 6-week research internship in Kyrgyzstan. Prof. Silvia Ulrich will dedicate additional Swiss staff for preparing and conducting this project during the years 2024 and 2025. The Kyrgyz team led by Prof. Sooronbaev (The Kyrgyz director of the Swiss-Kyrgyz High Altitude Research Center and Director of the University Hospital in Bishkek) will contribute the required personnel to recruit participants and for data acquisition, facility maintenances at low and high altitude, and transportation.

In regard of the health and safety of the participants at 3600 m, the Kumtor Gold mine Facility owns a fully equipped medical station including supplemental oxygen, emergency medications, medical staff and ambulances to quickly evacuate to lower altitudes.

#### 2.4. Schedule and milestones

The schedule and milestones of the project are outlined in **Figure 5**. The project partners will meet several times per year to decide on scientific, logistic and administrative aspects and other questions that might arise. The study preparations will be started on January 2024 and will recruit 140 participants in the summer season 2024. Milestone 1.1 (M1.1) represents the successful completion of the first expedition. In 2025, the second expedition with another 140 participants will take place (M1.2). Data analyses will be conducted in the following months, and manuscript submission can be expected thereafter (M1.3). In 2026, analysis of secondary outcomes such as pooled blood samples, exercise, cerebrovascular and ventilatory responses will be scheduled (M1.4).

| **01.01.2024 to 31.12.2026 (3 years)** | **2024** | | | | | | | | | | | | **2025** | | | | | | | | | | | | **2026** | | | | | | | | | | | |
|---|---|---|---|---|---|---|---|---|---|---|---|---|---|---|---|---|---|---|---|---|---|---|---|---|---|---|---|---|---|---|---|---|---|---|---|---|
| | J | F | M | A | M | J | J | A | S | O | N | D | J | F | M | A | M | J | J | A | S | O | N | D | J | F | M | A | M | J | J | A | S | O | N | D |
| Preparation | | | | | | | | | | | | | | | | | | | | | | | | | | | | | | | | | | | | |
| Data Collection | | | | **n = 140** | | | | | | M1.1 | | | | | | **n = 140** | | | | | | M1.2 | | | | | | | | | | | | | | |
| Interim Analysis | | | | | | | | | | | | | | | | | | | | | | | | | | | | | | | | | | | | |
| Primary data analysis | | | | | | | | | | | | | | | | | | | | | | | | | | | | M1.3 | | | | | | | | |
| Secondary data analysis | | | | | | | | | | | | | | | | | | | | | | | | | | | | | | | | | | | M1.4 | |
| | J | F | M | A | M | J | J | A | S | O | N | D | J | F | M | A | M | J | J | A | S | O | N | D | J | F | M | A | M | J | J | A | S | O | N | D |
| | **2024** | | | | | | | | | | | | **2025** | | | | | | | | | | | | **2026** | | | | | | | | | | | |

***Figure 5. Schedule and Milestones of the proposed study.*** *Grey areas indicate periods of scheduled work packages; Letters represent months of the years 2024-2026.*

#### 2.5. Relevance and impact

##### 2.5.1. Scientific relevance

Previous research has been conducted to elucidate AMS susceptibility, prevention, and treatments, but females have only occasionally been included and have never been appropriately characterized. No randomized clinical trial has been conducted to investigate and compare the efficacy of acetazolamide against AMS incidence in females compared to men, despite the knowledge that females are more prone to develop AMS compared to men. Differences in drug efficacy or side-effects based on sex-related differences are indicated in the literature and are likely due to lower blood volume and consequently higher acetazolamide plasma concentration, due to differences in the hypoxic chemosensitivity or other factors between females and men. The current gaps in our knowledge are mainly due to the demanding logistical and organisational challenges in conducting large-scale mountain medicine and physiology research, especially in females. The applicant and his partners have acquired the knowledge and study setting to overcome these challenges. Therefore, this study will provide conclusive results on the efficacy and side-effect of preventive acetazolamide against AMS in females compared to males and whether AMS susceptibility depends on female sex hormones or MCPs. Moreover, physiological, urine and blood analyses will provide comprehensive insights into physiological and clinical differences between females and males under hypoxic conditions. Findings from this large-scale project are expected to become milestones in the research history of high-altitude medicine, opening up new avenues for mechanistic research related to sex- and hormone-dependent tolerance to hypoxia.

##### 2.5.2. Broader Impact

Females are substantially underrepresented in heart failure, acute coronary syndrome and pulmonary disease studies, with females <55 years accounting for <10% of the study population and in case of an incident females present with a worse outcome.[30,145,146] The literature shows that many lung diseases are more commonly found in females and present with higher degree of severity, exacerbation rate, hospitalizations and mortality than in men.[147] In this regard, insights from this altitude study investigating hypoxia- and hypoxemia-related physiological and clinical adaptations in females compared to males might enhance our understanding of the known sex-related differences in lung diseases at low altitude. Furthermore, to date, there is limited performance of hormonal and cell immunological diagnostic endeavour and low inclusion of female participants in clinical trials on AMS prevention. Our findings are expected to have a large and broad impact, since a considerable number of summer and estimated 400 million annual winter visits will eventually be affected by altitude-related health impairments.[5,148] Based on the expected results, female athletes might be able to improve their high altitude performance or live-high, train-low training schedules based on the MCP. Exercise training based on the MCP is already common in top athletes, therefore, sports in the mountains are equally affected. Understanding sex differences in altitude tolerance may finally result in less accidents and emergency evacuations, as well as improved work efficiency in employees temporarily working at high altitudes. Moreover, translational knowledge transfer of these findings from hypoxemic conditions might contribute to a better understanding of acute cardiopulmonary diseases associated with hypoxemia (i.e. COVID), their progression, their therapy, and any related sex differences as well as to millions of highlanders living at altitudes >2500 m, who are chronically hypoxemic.

2. **Statistical analysis plan (SAP)**
   a. **Summary of SAP changes**

| Date | Protocol version | SAP version | Section number changed | Explanation |
|---|---|---|---|---|
| 10.06.2024 | 1.0 | 1.0 | | SAP created according to JAMA Guidelines.[1] |
| 03.02.2025 | 1.2 | 1.1 | 2.2, 4.3, 4.5 | Section 2.2: Nicola Benjamin has performed the planned interim analysis, and the SAP has been updated in accordance of the blinded study design modifications. Kay von Grünigen was added as project statistician. Section 4.3: The sample size estimation has been updated. Section 4.5: The planned interim analysis has been performed and the section has been updated. |
| 08.08.2025 | 1.2 | 1.2 | 7.2 | Section 7.2: The primary analysis using the Cochran Mantel-Haenszel (CMH) test was replaced with a modified Poisson regression with robust variance. The reason for this change lies in the inability of the CMH test to perform the originally intended aim to compared sex-related relative risk reductions of acetazolamide for AMS. |

### Sex-specific efficacy and safety of preventive acetazolamide for acute mountain sickness in healthy lowlanders: a randomised, double-blind, placebo-controlled trial.

### Statistical analysis plan

Trial registration: NCT06499727

Version 1.0
Date: June 10th, 2024

This document has been written based on information contained in the study protocol version 1.0, dated 10th June 2024.

### 2 Administrative Information

#### 2.1 Statistical analysis plan – revision history

| Protocol version | Updated SAP version No. | Section number changed | Description of and reason for change | Date changed |
|---|---|---|---|---|
| 1.0 | 1.0 | | SAP created according to JAMA Guidelines.[1] | 10.06.2024 |

#### 2.2 Roles and responsibility

Author: Aijan Taalaibekova[1]
Statistician: Prof. Michael Furian[2]
Chief investigator: Prof. Michael Furian[2]

Affiliations:
*[1]National Center of Cardiology and Internal Medicine, Pulmonology Department, Bishkek, Kyrgyzstan;*
*[2]University Hospital Zurich, Pulmonology Department, Zurich, Switzerland;*

#### 2.3 Signatures of Approval

Date: 10.06.2024
Version: 1.0

| Signatures | | | | | | |
|---|---|---|---|---|---|---|
| | **Name** | **Trial Role** | | **Signature** | | **Date** |
| | Michael Furian | Sponsor Investigator | | 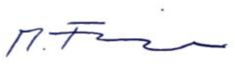 | | 10.06.2024 |
| | Aijan Taalaibekova | Principal Investigator | | 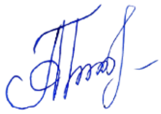 | | 10.06.2024 |

### 3 Introduction

#### 3.1 Background and rationale

Millions of people travel to high altitude for work or leisure activities and are exposed to reduced inspiratory oxygen partial pressure and hypoxemia that may lead to altitude illness, among which the most common form is acute mountain sickness (AMS). The main AMS symptoms are headache, malaise, weakness, and fatigue. Prospective studies have shown that 20–60% of newcomers at 2500–4000 m develop AMS requiring them to take medications, while, at very high altitudes, AMS may progress to high altitude cerebral oedema. Whether females are more susceptible to AMS remains insufficiently understood since no prospective study controlled for sex hormones, use of hormone contraception or assessed menstrual cycle phase (MCP) at altitude. Therefore, females remain underrepresented and poorly characterized in high altitude studies. In addition, the efficacy and safety of 250 mg/day acetazolamide, the standard recommendation for AMS prevention, has never been compared between sexes, although, females have presumably higher acetazolamide plasma concentration due to lower blood volume. Given the known dose-dependent preventive but also side effects of acetazolamide and equal proportion of females and males among mountain travellers, there is an urgent need to conclusively quantify the efficacy and safety of preventive acetazolamide therapy against AMS in females compared to males.

#### 3.2 Objectives

##### 3.2.1 Primary objective and hypothesis

The primary objective of this study is to compare the effectiveness of preventive acetazolamide treatment in reducing the incidence of AMS during a 2-day sojourn at 3600 m in healthy female compared to male lowlanders. AMS is defined as present, when a participant has a Lake Louise Score (LLS, 2018 version) ≥3 during at least one assessment while staying at 3600 m.

The primary hypothesis will be that, during the time course of 2 days/nights at 3600 m, preventive 250 mg/day acetazolamide therapy starting 24 hours before ascending, will reduce the AMS incidence significantly more in females compared to males. AMS will be defined as a Lake Louise questionnaire score of ≥3 points including headache.[2]

##### 3.2.2 Secondary objectives

Secondary objectives are:

a. To study the AMS incidence in females and males under placebo intervention at 3600 m.
b. To study differences in incidence of AMS between acetazolamide and placebo, and between females and males, using different AMS assessments, the LLS and the environmental symptoms questionnaire.
c. To compare altitude-induced changes in vital parameters and in parameters from arterial blood gas analysis and respiratory sleep studies between drugs and sexes.
d. To assess differences in cumulative incidence and severity of medication side effects between drugs and sexes during the stay at 3600 m.
e. To assess acetazolamide plasma concentration in females and males in the morning after spending one night at 3600 m.

### 4 Trial methods

#### 4.1 Trial design

This study is a single centre, randomised, double-blind, placebo-controlled parallel trial. Healthy participants are randomised in a 1:1 allocation ratio to either 250 mg of acetazolamide per day (125 mg in the morning, 125 mg in the evening) or identically looking placebo capsules as control. Treatment will start after baseline measurements in Bishkek, Kyrgyzstan (760m) 24h before ascent to 3500m and will end after the 2-day sojourn at Kumtor Gold Mine at 3600 m.

#### 4.2 Randomisation and blinding

Stratified randomization will be conducted for females and males using a computer generated schedule (MinimPy 0.3), stratifying for sex in a female-to-male ratio of 2:1.[3] Within sex, participants will be 1:1

randomized to A: 250 mg/day acetazolamide (125 mg in the morning with breakfast, 125 mg in the evening with dinner) starting 24 hours before arriving and while staying for 2 days and nights at 3600 m (total of 6 capsules of 125 mg acetazolamide) or B: identically looking, organoleptic placebo capsules. An independent pharmacist will prepare identically looking verum and placebo capsules labelled with secret codes. The list of codes will be kept confidential to investigators and participants until data acquisition and analysis has been completed.

#### 4.3 Sample size

Sample size estimation was performed using the Cochran Mantel-Haenszel test with continuity correction. The previously described preventive effect of acetazolamide against AMS has been suggested to be 44%.[4] Based on previous studies and a pilot study conducted by our research team, we expected an AMS incidence of 50% in females and 30% in males at an altitude of 3600 m (data not yet published). To detect a clinically meaningful efficacy difference of 20% of acetazolamide therapy against AMS in females compared to males (i.e. treatment effect of 50% in females and 30% in males) with a group allocation ratio of 2:1, power of 90%, two-sided alpha level of 0.05 and accounting for dropouts, a total of 180 females and 90 males will be recruited. The female-to-male ratio of 2:1 was chosen to enable conclusive testing of the additional hypotheses within females (see study protocol section 2.3.3).

#### 4.4 Framework

With the primary objective of determining whether acetazolamide reduces the incidence of AMS more in females compared to males, this study has been designed as a superiority trial. Secondary objectives will also be tested for superiority.

#### 4.5 Statistical interim analysis

A planned, blinded interim-analysis on the primary outcome will be performed after the completion of the first study year (2024). The statistician Dr. Nicola Benjamin will report to a data monitoring and safety committee that will make a recommendation to the steering committee on continuation or termination of the trial or on any appropriate adaptation of the protocol such as changes of eligibility criteria, sample size adaption, allocation ratio, or drug dose. In the interim analysis, symmetric stopping boundaries at $P<0.001$ will be applied (Peto approach) to account for strong efficacy or harm of acetazolamide. In the final analysis a $P<0.05$ will be considered statistically significant.

#### 4.6 Timing of final analysis

Final analysis will take place collectively for all outcomes at the end of the second year (2025) after completion of the measurements at altitude, and when the target sample size will be achieved.

#### 4.7 Timing of outcome assessments

As described in Figure 3 of the study protocol, participants will undergo recruitment and baseline measurements in Bishkek, Kyrgyzstan. After randomisation, participants will be invited back for a two-day high-altitude sojourn at Kumtor Gold Mine, Kyrgyzstan, at an altitude of 3600 m, with participants according to their randomisation taking either acetazolamide or placebo, beginning 24h before ascent and ending with the last day at altitude. The schedule of study procedures performed at baseline and at altitude is described in detail in Table 3 of the study protocol. The questionnaires related to the primary outcome AMS will be completed at 3600 m after arrival, as well as in the mornings and evenings. In case of symptom worsening, the participants and staff are instructed to complete additional questionnaires.

### 5 Statistical Principles

#### 5.1 Confidence Intervals and p-values

All applicable statistical tests will be two-sided with a significance level of 5%. Confidence intervals will be presented at the 95% level and will be two-sided.

#### 5.2 Adherence and protocol deviations

Adherence will be defined as taking all doses of the prescribed drug over the scheduled duration. We will describe number and percentage of participants adherent to treatment, with results provided by treatment group. Non-adherent participants, as in participants not taking all the anticipated doses, will not be included in the per-protocol analysis. Participants in whom exclusion and inclusion criteria will be applied erroneously will be included in the intention-to-treat but not in the per-protocol analysis.

Protocol deviations are classified prior to unblinding. Number and percentage of participants with protocol deviations will be summarised by treatment group. Participants with protocol deviations will be excluded from the per-protocol analysis. No formal statistical testing will be undertaken.

#### 5.3 Analysis populations

The primary analysis will be performed in the intention-to-treat-population. The intention-to-treat-population will include all participants randomised.
Additional analyses will be performed on the per-protocol population, defined as all participants not erroneously randomized, with no major protocol deviations, who adhered to treatment, and completed the measurements required in the respective analysis.

### 6 Trial population

#### 6.1 Screening data

The total number of people assessed for eligibility and the number of participants randomised will be presented in a flow chart together with the number and reasons for pre- and post-randomization exclusions.

#### 6.2 Eligibility

Inclusion and exclusion criteria for this data are summarised in the paragraph “2.3.5 Participants” in the study protocol. The number of ineligible participants randomised will be reported together with the reasons for ineligibility in the flow chart mentioned above.

#### 6.3 Recruitment

A CONSORT flow diagram will be used to summarise the following information regarding participant numbers and reasons for exclusion:
- Number of people assessed for eligibility according to predefined criteria
- Number of ineligible people screened, including reasons for ineligibility
- Number of eligible people randomised
- Number of participants excluded due to ineligibility post-randomisation
- Number of participants randomised undergoing intervention

#### 6.4 Withdrawal

Number and reasons for withdrawal will be presented in the CONSORT diagram mentioned above.
Data of participants who withdraw from the study after randomisation by not showing up for the high-altitude sojourn and data of participants actively withdrawing consent to data collection will not be included in the final analysis. Data of participants withdrawing consent to medication intake but not to data collection are planned to be included in the intention-to-treat analysis.

#### 6.5 Baseline characteristics

Participants included in the intention-to-treat analysis will be described both overall and separately for sex and drug groups. Aspects described will be age, height, weight and body mass index.
Continuous data will be represented as mean and standard deviation. Tests of statistical significance will be conducted between sexes but not between drugs.

### 7 Analysis

#### 7.1 Outcome definitions

The primary outcome of this study is defined as the sex-related mean difference of acetazolamide efficacy for AMS incidence. The incidence of AMS is measured with the help of the 2018 Lake Louise Score (LLS), a validated questionnaire for AMS. For AMS to be present, a subject must have an LLS of ≥3. Headache must be present in addition to one of the following symptoms, rated on a scale of 0 to 3 (0 = not present, 1 = mild, 2 = moderate, 3

= severe): gastrointestinal discomfort, fatigue or weakness, and dizziness or light-headedness. The sum of the responses on these questions is then calculated to attain the Lake Louise Score.
The LLS is measured at several timepoints specified in table 2 of the study protocol. If a participant scores an LLS ≥3 at one timepoint during the stay at altitude, the participant is deemed to have AMS, independent of the LLS measured at later timepoints.

As a secondary outcome AMS severity is compared between sexes and treatment groups using the scores obtained from the LLS for every timepoint at which measurements were made at altitude.
Furthermore, the incidence of AMS is compared using different measurement systems, on the one hand with the LLS described above, on the other hand with the environmental symptoms questionnaire (AMS-c). A cut-off of ≥ 0.7 for the AMS-c is used. Measurements occurred at the same timepoints as the LLS.
Additional LLS cut-offs and version will be compared. First the 2018 vs. 1993 LLS will be compared, as well as LLS ≥6 will represent the incidence of moderate-to-severe AMS.

Incidence and severity of side effects are evaluated using a standardised questionnaire during clinical examination. The severity of the following side effects was recorded on a scale from none over slightly and moderate to severe:
- tingling sensation
- impaired sense of taste
- increased urinary urge
- others (specified)

Data collection will be carried out at the same timepoints as the assessment of AMS and can be looked up in table 2 of the study protocol.

Vital parameters and parameters from arterial blood gas analysis are obtained. For the specific timepoints of measurement please refer to table 2 of the study protocol. It is planned to assess the difference in sex- and altitude-induced changes between the acetazolamide and placebo group of the following variables:

- Vital parameters
  - Arterial blood pressure [mmHg]
  - Heart rate [bpm]
  - Oxygen saturation ($SpO_2$) [%]
- Arterial blood gas and dried blood spot analyses
  - Arterial partial pressure of oxygen ($PaO_2$) [kPa]
  - Arterial partial pressure of carbon dioxide ($PaCO_2$) [kPa]
  - Arterial oxygen saturation ($SaO_2$) [%]
  - Bicarbonate [mM]
  - pH
  - Hemoglobin [g/l]
  - Hematocrit [%]
  - Acetazolamide plasma concentration

#### 7.2 Analysis methods

The primary outcome will be analysed in the intention-to-treat population, secondary outcomes in the per-protocol population. The primary analysis will be a Cochran Mantel-Haenszel test comparing the sex-related difference in the acetazolamide effect against AMS. For other AMS-related comparisons, Chi-square statistics and Cox-proportional hazard models will be applied.

For secondary outcomes such as vital parameters, results from arterial blood gases, mixed regression models will be fitted with sex, drug, altitude, and their interaction as fixed effects, and participants as random effects. It is planned to present unadjusted models, if however, imbalances in baseline characteristics between the acetazolamide and placebo group are found, additional analysis with adjustment for these factors will be performed.
The incidence and severity of medication side effects will be evaluated using Chi-square statistics or Fisher's Exact tests.

#### 7.3 Missing data

Missing values in the primary outcome due to premature termination of the high-altitude sojourn will be imputed as being AMS positive. This approach is chosen to avoid overestimation of a potential treatment effect of acetazolamide. Sensitivity analyses of the primary outcome include the analysis in the per-protocol population. Missing values in secondary outcomes will not be replaced.

#### 7.4 Additional analyses

Analyses related to the primary outcome AMS, defined by the 2018 LLS, will be repeated for the 1993 LLS AMS definition. This analysis will allow to compare the novel findings with previously published AMS data.

#### 7.5 Harms

For every treatment arm the number and percentage of participants experiencing adverse events (or serious adverse events, if any) will be presented categorized by severity. No formal statistical testing will be undertaken.

#### 7.6 Statistical software

Statistical analyses will be performed with Stata version 14.2 or higher, and R software. Other packages may be used if necessary.

### Sex-specific efficacy and safety of preventive acetazolamide for acute mountain sickness in healthy lowlanders: a randomised, double-blind, placebo-controlled trial.

#### Statistical analysis plan

Trial registration: NCT06499727

Version 1.1
Date: February 3rd, 2025

This document has been written based on information contained in the study protocol version 1.2, dated 3rd February 2025.

### 2 Administrative Information

#### 2.1 Statistical analysis plan – revision history

| Protocol version | Updated SAP version No. | Section number changed | Description of and reason for change | Date changed |
|---|---|---|---|---|
| 1.0 | 1.0 | | SAP created according to JAMA Guidelines.[1] | 10.06.2024 |
| 1.2 | 1.1 | 2.2, 4.3, 4.5 | Section 2.2: Nicola Benjamin has performed the planned interim analysis, and the SAP has been updated in accordance of the blinded study design modifications. Kay von Grünigen was assigned as the project Statistician<br><br>Section 4.3: The sample size estimation has been updated.<br><br>Section 4.5: The planned interim analysis has been performed and the section has been updated. | 03.02.2025 |

#### 2.2 Roles and responsibility

Author: Aijan Taalaibekova[1]
Statistician: Kay von Grünigen[2]
Chief investigator: Michael Furian[2]

Contributor: Dr. Nicola Benjamin[3]

Affiliations: [1]National Center of Cardiology and Internal Medicine, Pulmonology Department, Bishkek, Kyrgyzstan;
[2]University Hospital Zurich, Pulmonology Department, Zurich, Switzerland;
[3]Center for Pulmonary Hypertension, Heidelberg University Hospital, Heidelberg, Germany.

#### 2.3 Signatures of Approval

Date: 03.02.2025
Version: 1.1

| Signatures | | | |
|---|---|---|---|
| Name | Trial Role | Signature | Date |
| Michael Furian | Sponsor Investigator | 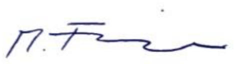 | 03.02.2025 |

| | | | | | | |
|---|---|---|---|---|---|---|
| | Aijan Taalaibekova | Principal Investigator | | | | 03.02.2025 |
| | Kay von Grünigen | Statistician | | | | 03.02.2025 |

### 3 Introduction

#### 3.1 Background and rationale

Millions of people travel to high altitude for work or leisure activities and are exposed to reduced inspiratory oxygen partial pressure and hypoxemia that may lead to altitude illness, among which the most common form is acute mountain sickness (AMS). The main AMS symptoms are headache, malaise, weakness, and fatigue. Prospective studies have shown that 20–60% of newcomers at 2500–4000 m develop AMS requiring them to take medications, while, at very high altitudes, AMS may progress to high altitude cerebral oedema. Whether females are more susceptible to AMS remains insufficiently understood since no prospective study controlled for sex hormones, use of hormone contraception or assessed menstrual cycle phase (MCP) at altitude. Therefore, females remain underrepresented and poorly characterized in high altitude studies. In addition, the efficacy and safety of 250 mg/day acetazolamide, the standard recommendation for AMS prevention, has never been compared between sexes, although, females have presumably higher acetazolamide plasma concentration due to lower blood volume. Given the known dose-dependent preventive but also side effects of acetazolamide and equal proportion of females and males among mountain travellers, there is an urgent need to conclusively quantify the efficacy and safety of preventive acetazolamide therapy against AMS in females compared to males.

#### 3.2 Objectives

##### 3.2.1 Primary objective and hypothesis

The primary objective of this study is to compare the effectiveness of preventive acetazolamide treatment in reducing the incidence of AMS during a 2-day sojourn at 3600 m in healthy female compared to male lowlanders. AMS is defined as present, when a participant has a Lake Louise Score (LLS, 2018 version) ≥3 during at least one assessment while staying at 3600 m.

The primary hypothesis will be that, during the time course of 2 days/nights at 3600 m, preventive 250 mg/day acetazolamide therapy starting 24 hours before ascending, will reduce the AMS incidence significantly more in females compared to males. AMS will be defined as a Lake Louise questionnaire score of ≥3 points including headache.[2]

##### 3.2.2 Secondary objectives

Secondary objectives are:

a. To study the AMS incidence in females and males under placebo intervention at 3600 m.
b. To study differences in incidence of AMS between acetazolamide and placebo, and between females and males, using different AMS scores, the LLS and the environmental symptoms questionnaire.
c. To compare altitude-induced changes in vital parameters and in parameters from arterial blood gas analysis and respiratory sleep studies between drugs and sexes.
d. To assess differences in cumulative incidence and severity of medication side effects between drugs and sexes during the stay at 3600 m.
e. To assess acetazolamide plasma concentration in females and males in the morning after spending one night at 3600 m.

### 4 Trial methods

#### 4.1 Trial design

This study is a single centre, randomised, double-blind, placebo-controlled parallel trial. Healthy participants are randomised in a 1:1 allocation ratio to either 250 mg of acetazolamide per day (125 mg in the morning, 125 mg in the evening) or identically looking placebo capsules as control. Treatment will start after baseline measurements in Bishkek, Kyrgyzstan (760m) 24h before ascent to 3500m and will end after the 2-day sojourn at Kumtor Gold Mine at 3600 m.

#### 4.2 Randomisation and blinding

Stratified randomization will be conducted for females and males using a computer generated schedule (MinimPy 0.3), stratifying for sex in a female-to-male ratio of 2:1.[3] Within sex, participants will be 1:1 randomized to A: 250 mg/day acetazolamide (125 mg in the morning with breakfast, 125 mg in the evening with dinner) starting 24 hours before arriving and while staying for 2 days and nights at 3600 m (total of 6 capsules of 125 mg acetazolamide) or B: identically looking, organoleptic placebo capsules. An independent pharmacist will prepare identically looking verum and placebo capsules labelled with secret codes. The list of codes will be kept confidential to investigators and participants until data acquisition and analysis has been completed.

#### 4.3 Sample size

Sample size estimation was performed using the Cochran Mantel-Haenszel test with continuity correction. The previously described preventive effect of acetazolamide against AMS has been suggested to be 44%.[4] Based on previous studies and a pilot study conducted by our research team, we expected an AMS incidence of 50% in females and 30% in males at an altitude of 3600 m (data not yet published). To detect a clinically meaningful efficacy difference of 20% of acetazolamide therapy against AMS in females compared to males (i.e. treatment effect of 50% in females and 30% in males) with a group allocation ratio of 2:1, power of 90%, two-sided alpha level of 0.05 and accounting for dropouts, a total of 180 females and 90 males will be recruited. The female-to-male ratio of 2:1 was chosen to enable conclusive testing of the additional hypotheses within females (see study protocol section 2.3.3).
Based on the findings of the interim analysis (see section 4.5), the total number of participants and the proportion of males should be increases during the second part of the project. A total of 180 females and 120 males will be recruited, and the female-to-male ratio of 2:1 has been dismissed.

#### 4.4 Framework

With the primary objective of determining whether acetazolamide reduces the incidence of AMS more in females compared to males, this study has been designed as a superiority trial. Secondary objectives will also be tested for superiority.

#### 4.5 Statistical interim analysis

A planned, blinded interim-analysis on the primary outcome has been performed by Dr. Nicolas Benjamin on 16th of January 2025. Findings are shown in Table 1 and 2. Based on the interim analysis, the AMS incidences and treatment effects in females and males deviated from the stated values defined in the study protocol 1.0. Therefore, sample size re-estimation has been discussed. The Scientific Committee concluded, that the study should continue but that **Option C** (Table 2) should be applied. Based on this decision, a total of 180 females and 120 males, will be randomized.

**Table 1. Acute mountain sickness incidences obtained during the planned blinded interim analysis in January, 2025.**

| | Drug 1 | | Drug 2 | |
|---|---|---|---|---|
| | Females | Males | Females | Males |
| AMS positive | **19 (44.2%)** | **4 (16.0%)** | **13 (30.2%)** | **3 (13.6%)** |
| AMS negative | 24 (55.8%) | 21 (84.0%) | 30 (69.8%) | 19 (86.4%) |

| | | | | |
|---|---|---|---|---|
| Total | 43 (100%) | 25 (100%) | 43 (100%) | 22 (100%) |

**Table 2. Possible scenarios for sample size re-estimation discussed during the interim analysis.**

| Version | Considerations | AMS f/m | Treatment effect f/m | Power | Required N f/m (without dropouts) |
|---|---|---|---|---|---|
| Original | Original sample size estimation | 50% / 30% | 50% / 30% | 90% | 140 / 70 |
| Option A | 20% treatment difference with current AMS incidences | 44% / 16% | 30% / 10% | 90% | 540 / 270 |
| Option B | Lowering power to 80% | 44% / 16% | 30% / 10% | 80% | 400 / 200 |
| Option C | Assuming Tx effects to be unchanged | 44% / 16% | 50% / 30% | 90% | 180 / 80 |
| Option D | Lowering power to 80% | 44% / 16% | 50% / 30% | 80% | 130 / 60 |
| Option E | Interim incidences not reliable, stay with original assumptions, but use new Tx difference | 50% / 30% | 30% / 10% | 90% | 460 / 220 |
| Option F | Interim incidences and Tx difference not reliable, stay with original plan | 50% / 30% | 50% / 30% | 90% | 140 / 70 |

#### 4.6 Timing of final analysis

Final analysis will take place collectively for all outcomes at the end of the second year (2025) after completion of the measurements at altitude, and when the target sample size will be achieved.

#### 4.7 Timing of outcome assessments

As described in Figure 3 of the study protocol, participants will undergo recruitment and baseline measurements in Bishkek, Kyrgyzstan. After randomisation, participants will be invited back for a two-day high-altitude sojourn at Kumtor Gold Mine, Kyrgyzstan, at an altitude of 3600 m, with participants according to their randomisation taking either acetazolamide or placebo, beginning 24h before ascent and ending with the last day at altitude. The schedule of study procedures performed at baseline and at altitude is described in detail in Table 3 of the study protocol. The questionnaires related to the primary outcome AMS will be completed at 3600 m after arrival, and in the mornings and evenings. In case of symptom worsening, the participants and staff are instructed to complete additional questionnaires.

### 5 Statistical Principles

#### 5.1 Confidence Intervals and p-values

All applicable statistical tests will be two-sided with a significance level of 5%. Confidence intervals will be presented at the 95% level and will be two-sided.

#### 5.2 Adherence and protocol deviations

Adherence will be defined as taking all doses of the prescribed drug over the scheduled duration. We will describe number and % of participants adherent to treatment, with results provided by treatment group. Non-adherent participants, as in participants not taking all the anticipated doses, will not be included in the per-protocol analysis. Participants in whom exclusion and inclusion criteria will be applied erroneously will be included in the intention-to-treat but not in the per-protocol analysis.

Protocol deviations are classified prior to unblinding. Number and percentage of participants with protocol deviations will be summarised by treatment group. Participants with protocol deviations will be excluded from the per-protocol analysis. No formal statistical testing will be undertaken.

#### 5.3 Analysis populations

The primary analysis will be performed in the intention-to-treat-population. The intention-to-treat-population will include all participants randomised.
Additional analyses will be performed on the per-protocol population, defined as all participants not erroneously randomized, with no major protocol deviations, who adhered to treatment, and completed the measurements required in the respective analysis.

### 6 Trial population

#### 6.1 Screening data

The total number of people assessed for eligibility and the number of participants randomised will be presented in a flow chart together with the number and reasons for pre- and post-randomization exclusions.

#### 6.2 Eligibility

Inclusion and exclusion criteria for this data are summarised in the paragraph “2.3.5 Participants” in the study protocol. The number of ineligible participants randomised will be reported together with the reasons for ineligibility in the flow chart mentioned above.

#### 6.3 Recruitment

A CONSORT flow diagram will be used to summarise the following information regarding participant numbers and reasons for exclusion:
- Number of people assessed for eligibility according to predefined criteria
- Number of ineligible people screened, including reasons for ineligibility
- Number of eligible people randomised
- Number of participants excluded due to ineligibility post-randomisation
- Number of participants randomised undergoing intervention

#### 6.4 Withdrawal

Number and reasons for withdrawal will be presented in the CONSORT diagram mentioned above.
Data of participants who withdraw from the study after randomization by not showing up for the high-altitude sojourn and data of participants actively withdrawing consent to data collection will not be included in the final analysis. Data of participants withdrawing consent to medication intake but not to data collection are planned to be included in the intention-to-treat analysis.

#### 6.5 Baseline characteristics

Participants included in the intention-to-treat analysis will be described both overall and separately for sex and drug groups. Aspects described will be age, height, weight and body mass index.
Continuous data will be represented as mean and standard deviation. Tests of statistical significance will be conducted between sexes but not between drugs.

### 7 Analysis

#### 7.1 Outcome definitions

The primary outcome of this study is defined as the sex-related mean difference of acetazolamide efficacy for AMS incidence. The incidence of AMS is measured with the help of the 2018 Lake Louise Score (LLS), a validated questionnaire for AMS. For AMS to be present, a subject must have an LLS of ≥3. Headache must be present in addition to one of the following symptoms, rated on a scale of 0 to 3 (0 = not present, 1 = mild, 2 = moderate, 3

= severe): gastrointestinal discomfort, fatigue or weakness, and dizziness or light-headedness. The sum of the responses on these questions is then calculated to attain the Lake Louise Score.
The LLS is measured at several timepoints specified in table 2 of the study protocol. If a participant scores an LLS ≥3 at one timepoint during the stay at altitude, the participant is deemed to have AMS, independent of the LLS measured at later timepoints.

As a secondary outcome AMS severity is compared between sexes and treatment groups using the scores obtained from the LLS for every timepoint at which measurements were made at altitude.
Furthermore, the incidence of AMS is compared using different measurement systems, on the one hand with the LLS described above, on the other hand with the environmental symptoms questionnaire (AMS-c). A cut-off of ≥ 0.7 for the AMS-c is used as definition of AMS incidence. Measurements occurred simultaneously with the LLS assessment.
Additional LLS cut-offs and questionnaire versions will be compared. First the 2018 vs. 1993 LLS will be compared, as well as LLS ≥6 will represent the incidence of moderate-to-severe AMS.

Incidence and severity of side effects are evaluated using a standardised questionnaire during clinical examination. The severity of the following side effects was recorded on a scale from none over slightly and moderate to severe:
- tingling sensation
- impaired sense of taste
- increased urinary urge
- others (specified)

Data collection will be carried out at the same timepoints as the assessment of AMS and can be looked up in table 2 of the study protocol.

Vital parameters and parameters from arterial blood gas analysis are obtained. For the specific timepoints of measurement please refer to table 2 of the study protocol. It is planned to assess the difference in sex- and altitude-induced changes between the acetazolamide and placebo group of the following variables:

- Vital parameters
    - Arterial blood pressure [mmHg]
    - Heart rate [bpm]
    - Oxygen saturation ($SpO_2$) [%]
- Arterial blood gas and dried blood spot analyses
    - Arterial partial pressure of oxygen ($PaO_2$) [kPa]
    - Arterial partial pressure of carbon dioxide ($PaCO_2$) [kPa]
    - Arterial oxygen saturation ($SaO_2$) [%]
    - Bicarbonate [mM]
    - pH
    - Haemoglobin [g/l]
    - Haematocrit [%]
    - Acetazolamide plasma concentration

#### 7.2 Analysis methods

The primary outcome will be analysed in the intention-to-treat population, secondary outcomes in the per-protocol population. The primary analysis will be a Cochran Mantel-Haenszel test comparing the sex-related difference in the acetazolamide effect against AMS. For other AMS-related comparisons, $Chi^2$-square statistics and Cox-proportional hazard models will be applied.

For secondary outcomes such as vital parameters, results from arterial blood gases, mixed regression models will be fitted with sex, drug, altitude, and their interaction sex*drug*altitude as fixed effects, and participants as random effects. It is planned to present unadjusted models, if however, imbalances in baseline characteristics between the acetazolamide and placebo group are found, additional analysis with adjustment for these factors will be performed.
The incidence and severity of medication side effects will be evaluated using Chi-square statistics or Fisher's Exact tests.

#### 7.3 Missing data

Missing values in the primary outcome due to premature termination of the high-altitude stay will be imputed as being AMS positive. This approach is chosen to avoid overestimation of a potential treatment effect of acetazolamide. Sensitivity analyses of the primary outcome include the analysis in the per-protocol population. Missing values in secondary outcomes will not be replaced.

#### 7.4 Additional analyses

Analyses related to the primary outcome AMS, defined by the 2018 LLS, will be repeated for the 1993 LLS AMS definition. This analysis will allow to compare the novel findings with previously published AMS data.

#### 7.5 Harms

For every treatment arm the number and percentage of participants experiencing adverse events (or serious adverse events, if any) will be presented categorized by severity. No formal statistical testing will be undertaken.

#### 7.6 Statistical software

Statistical analyses will be performed with Stata version 14.2 or higher, and R software. Other packages may be used if necessary.

### Sex-specific efficacy and safety of preventive acetazolamide for acute mountain sickness in healthy lowlanders: a randomised, double-blind, placebo-controlled trial.

#### Statistical analysis plan

Trial registration: NCT06499727

Version 1.2
Date: August 8$^{th}$, 2025

This document has been written based on information contained in the study protocol version 1.2, dated 3$^{rd}$ February 2025.

### 2 Administrative Information

#### 2.1 Statistical analysis plan – revision history

| Protocol version | Updated SAP version No. | Section number changed | Description of and reason for change | Date changed |
|---|---|---|---|---|
| 1.0 | 1.0 | | SAP created according to JAMA Guidelines.[1] | 10.06.2024 |
| 1.2 | 1.1 | 2.2, 4.3, 4.5 | Section 2.2: Nicola Benjamin has performed the planned interim analysis, and the SAP has been updated in accordance of the blinded study design modifications.<br><br>Section 4.3: The sample size estimation has been updated.<br><br>Section 4.5: The planned interim analysis has been performed and the section has been updated. | 03.02.2025 |
| 1.2 | 1.2 | 7.2 | Section 7.2: The primary analysis using the Cochran Mantel-Haenszel (CMH) test was replaced with a modified Poisson regression with robust variance. The reason for this change lies in the inability of the CMH test to perform the originally intended aim to compared sex-related relative risk reductions of acetazolamide for AMS. | 08.08.2025 |

#### 2.2 Roles and responsibility

Author: Aijan Taalaibekova[1]
Statistician: Kay von Grünigen[2]
Chief investigator: Michael Furian[2]

Contributor: Nicola Benjamin[3]

Affiliations: [1]National Center of Cardiology and Internal Medicine, Pulmonology Department, Bishkek, Kyrgyzstan; [2]University Hospital Zurich, Pulmonology Department, Zurich, Switzerland; [3]Center for Pulmonary Hypertension, Heidelberg University Hospital, Heidelberg, Germany.

#### 2.3 Signatures of Approval

Date: 08.08.2025
Version: 1.2

| Signatures | | | |
|---|---|---|---|
| **Name** | **Trial Role** | **Signature** | **Date** |
| Michael Furian | Sponsor Investigator | | 08.08.2025 |
| Aijan Taalaibekova | Principal Investigator | | 08.08.2025 |
| Kay von Grünigen | Statistician | | 06.08.2025 |

### 3 Introduction

#### 3.1 Background and rationale

Millions of people travel to high altitude for work or leisure activities and are exposed to reduced inspiratory oxygen partial pressure and hypoxemia that may lead to altitude illness, among which the most common form is acute mountain sickness (AMS). The main AMS symptoms are headache, malaise, weakness, and fatigue. Prospective studies have shown that 20–60% of newcomers at 2500–4000 m develop AMS requiring them to take medications, while, at very high altitudes, AMS may progress to high altitude cerebral oedema. Whether females are more susceptible to AMS remains insufficiently understood since no prospective study controlled for sex hormones, use of hormone contraception or assessed menstrual cycle phase (MCP) at altitude. Therefore, females remain underrepresented and poorly characterized in high altitude studies. In addition, the efficacy and safety of 250 mg/day acetazolamide, the standard recommendation for AMS prevention, has never been compared between sexes, although, females have presumably higher acetazolamide plasma concentration due to lower blood volume. Given the known dose-dependent preventive but also side effects of acetazolamide and equal proportion of females and males among mountain travellers, there is an urgent need to conclusively quantify the efficacy and safety of preventive acetazolamide therapy against AMS in females compared to males.

#### 3.2 Objectives

##### 3.2.1 Primary objective and hypothesis

The primary objective of this study is to compare the effectiveness of preventive acetazolamide treatment in reducing the incidence of AMS during a 2-day sojourn at 3600 m in healthy female compared to male lowlanders. AMS is defined as present, when a participant has a Lake Louise Score (LLS, 2018 version) ≥3 during at least one assessment while staying at 3600 m.

The primary hypothesis will be that, during the time course of 2 days/nights at 3600 m, preventive 250 mg/day acetazolamide therapy starting 24 hours before ascending, will reduce the AMS incidence significantly more in females compared to males. AMS will be defined as a Lake Louise questionnaire score of ≥3 points including headache.[2]

##### 3.2.2 Secondary objectives

Secondary objectives are:

a. To study the AMS incidence in females and males under placebo intervention at 3600 m.
b. To study differences in incidence of AMS between acetazolamide and placebo, and between females and males, using different AMS scores, the LLS and the environmental symptoms questionnaire.

c. To compare altitude-induced changes in vital parameters and in parameters from arterial blood gas analysis and respiratory sleep studies between drugs and sexes.
d. To assess differences in cumulative incidence and severity of medication side effects between drugs and sexes during the stay at 3600 m.
e. To assess acetazolamide plasma concentration in females and males in the morning after spending one night at 3600 m.

### 4 Trial methods

#### 4.1 Trial design

This study is a single centre, randomised, double-blind, placebo-controlled parallel trial. Healthy participants are randomised in a 1:1 allocation ratio to either 250 mg of acetazolamide per day (125 mg in the morning, 125 mg in the evening) or identically looking placebo capsules as control. Treatment will start after baseline measurements in Bishkek, Kyrgyzstan (760m) 24h before ascent to 3500m and will end after the 2-day sojourn at Kumtor Gold Mine at 3600 m.

#### 4.2 Randomisation and blinding

Stratified randomization will be conducted for females and males using a computer generated schedule (MinimPy 0.3), stratifying for sex in a female-to-male ratio of 2:1.[3] Within sex, participants will be 1:1 randomized to A: 250 mg/day acetazolamide (125 mg in the morning with breakfast, 125 mg in the evening with dinner) starting 24 hours before arriving and while staying for 2 days and nights at 3600 m (total of 6 capsules of 125 mg acetazolamide) or B: identically looking, organoleptic placebo capsules. An independent pharmacist will prepare identically looking verum and placebo capsules labelled with secret codes. The list of codes will be kept confidential to investigators and participants until data acquisition and analysis has been completed.

#### 4.3 Sample size

Sample size estimation was performed using the Cochran Mantel-Haenszel test with continuity correction. The previously described preventive effect of acetazolamide against AMS has been suggested to be 44%.[4] Based on previous studies and a pilot study conducted by our research team, we expected an AMS incidence of 50% in females and 30% in males at an altitude of 3600 m (data not yet published). To detect a clinically meaningful efficacy difference of 20% of acetazolamide therapy against AMS in females compared to males (i.e. treatment effect of 50% in females and 30% in males) with a group allocation ratio of 2:1, power of 90%, two-sided alpha level of 0.05 and accounting for dropouts, a total of 180 females and 90 males will be recruited. The female-to-male ratio of 2:1 was chosen to enable conclusive testing of the additional hypotheses within females (see study protocol section 2.3.3).
Based on the findings of the interim analysis (see section 4.5), the total number of participants and the proportion of males should be increases during the second part of the project. A total of 180 females and 120 males will be recruited, and the female-to-male ratio of 2:1 has been dismissed.

#### 4.4 Framework

With the primary objective of determining whether acetazolamide reduces the incidence of AMS more in females compared to males, this study has been designed as a superiority trial. Secondary objectives will also be tested for superiority.

#### 4.5 Statistical interim analysis

A planned, blinded interim-analysis on the primary outcome has been performed by Dr. Nicolas Benjamin on 16th of January 2025. Findings are shown in Table 1 and 2. Based on the interim analysis, the AMS incidences and treatment effects in females and males deviated from the stated values defined in the study protocol 1.0. Therefore, sample size re-estimation has been discussed. The Scientific Committee concluded, that the study should continue but that **Option C** (Table 2) should be applied. Based on this decision, a total of 180 females and 120 males, will be randomized.

| **Table 1. Acute mountain sickness incidences obtained during the planned blinded interim analysis in January, 2025.** | | | | |
|---|---|---|---|---|
| | Drug 1 | | Drug 2 | |
| | Females | Males | Females | Males |
| AMS positive | **19 (44.2%)** | **4 (16.0%)** | **13 (30.2%)** | **3 (13.6%)** |
| AMS negative | 24 (55.8%) | 21 (84.0%) | 30 (69.8%) | 19 (86.4%) |
| Total | 43 (100%) | 25 (100%) | 43 (100%) | 22 (100%) |

**Table 2. Possible scenarios for sample size re-estimation discussed during the interim analysis.**

| **Considerations** | **Version** | **AMS f/m** | **Treatment effect f/m** | **Power** | **Required N f/m (without dropouts)** |
|---|---|---|---|---|---|
| Original sample size estimation | Original | 50% / 30% | 50% / 30% | 90% | 140 / 70 |
| 20% treatment difference with current AMS incidences | Option A | 44% / 16% | 30% / 10% | 90% | 540 / 270 |
| Lowering power to 80% | Option B | 44% / 16% | 30% / 10% | 80% | 400 / 200 |
| Assuming Tx effects to be unchanged | Option C | 44% / 16% | 50% / 30% | 90% | 180 / 80 |
| Lowering power to 80% | Option D | 44% / 16% | 50% / 30% | 80% | 130 / 60 |
| Interim incidences not reliable, stay with original assumptions, but use new Tx difference | Option E | 50% / 30% | 30% / 10% | 90% | 460 / 220 |
| Interim incidences and Tx difference not reliable, stay with original plan | Option F | 50% / 30% | 50% / 30% | 90% | 140 / 70 |

#### 4.6 Timing of final analysis

Final analysis will take place collectively for all outcomes at the end of the second year (2025) after completion of the measurements at altitude, and when the target sample size will be achieved.

#### 4.7 Timing of outcome assessments

As described in Figure 3 of the study protocol, participants will undergo recruitment and baseline measurements in Bishkek, Kyrgyzstan. After randomisation, participants will be invited back for a two-day high-altitude sojourn at Kumtor Gold Mine, Kyrgyzstan, at an altitude of 3600 m, with participants according to their randomisation taking either acetazolamide or placebo, beginning 24h before ascent and ending with the last day at altitude. The schedule of study procedures performed at baseline and at altitude is described in detail in Table 3 of the study protocol. The questionnaires related to the primary outcome AMS will be completed at 3600 m after arrival, and in the mornings and evenings. In case of symptom worsening, the participants and staff are instructed to complete additional questionnaires.

### 5 Statistical Principles

#### 5.1 Confidence Intervals and p-values

All applicable statistical tests will be two-sided with a significance level of 5%. Confidence intervals will be presented at the 95% level and will be two-sided.

#### 5.2 Adherence and protocol deviations

Adherence will be defined as taking all doses of the prescribed drug over the scheduled duration. We will describe number and % of participants adherent to treatment, with results provided by treatment group. Non-adherent participants, as in participants not taking all the anticipated doses, will not be included in the per-protocol analysis. Participants in whom exclusion and inclusion criteria will be applied erroneously will be included in the intention-to-treat but not in the per-protocol analysis.

Protocol deviations are classified prior to unblinding. Number and percentage of participants with protocol deviations will be summarised by treatment group. Participants with protocol deviations will be excluded from the per-protocol analysis. No formal statistical testing will be undertaken.

#### 5.3 Analysis populations

The primary analysis will be performed in the intention-to-treat-population. The intention-to-treat-population will include all participants randomised.
Additional analyses will be performed on the per-protocol population, defined as all participants not erroneously randomized, with no major protocol deviations, who adhered to treatment, and completed the measurements required in the respective analysis.

### 6 Trial population

#### 6.1 Screening data

The total number of people assessed for eligibility and the number of participants randomised will be presented in a flow chart together with the number and reasons for pre- and post-randomization exclusions.

#### 6.2 Eligibility

Inclusion and exclusion criteria for this data are summarised in the paragraph “2.3.5 Participants” in the study protocol. The number of ineligible participants randomised will be reported together with the reasons for ineligibility in the flow chart mentioned above.

#### 6.3 Recruitment

A CONSORT flow diagram will be used to summarise the following information regarding participant numbers and reasons for exclusion:
- Number of people assessed for eligibility according to predefined criteria
- Number of ineligible people screened, including reasons for ineligibility
- Number of eligible people randomised
- Number of participants excluded due to ineligibility post-randomisation
- Number of participants randomised undergoing intervention

#### 6.4 Withdrawal

Number and reasons for withdrawal will be presented in the CONSORT diagram mentioned above.
Data of participants who withdraw from the study after randomization by not showing up for the high-altitude sojourn and data of participants actively withdrawing consent to data collection will not be included in the final analysis. Data of participants withdrawing consent to medication intake but not to data collection are planned to be included in the intention-to-treat analysis.

#### 6.5 Baseline characteristics

Participants included in the intention-to-treat analysis will be described both overall and separately for sex and drug groups. Aspects described will be age, height, weight and body mass index.

Continuous data will be represented as mean and standard deviation. Tests of statistical significance will be conducted between sexes but not between drugs.

### 7 Analysis

#### 7.1 Outcome definitions

The primary outcome of this study is defined as the sex-related mean difference of acetazolamide efficacy for AMS incidence. The incidence of AMS is measured with the help of the 2018 Lake Louise Score (LLS), a validated questionnaire for AMS. For AMS to be present, a subject must have an LLS of ≥3. Headache must be present in addition to one of the following symptoms, rated on a scale of 0 to 3 (0 = not present, 1 = mild, 2 = moderate, 3 = severe): gastrointestinal discomfort, fatigue or weakness, and dizziness or light-headedness. The sum of the responses on these questions is then calculated to attain the Lake Louise Score.
The LLS is measured at several timepoints specified in table 2 of the study protocol. If a participant scores an LLS ≥3 at one timepoint during the stay at altitude, the participant is deemed to have AMS, independent of the LLS measured at later timepoints.

As a secondary outcome AMS severity is compared between sexes and treatment groups using the scores obtained from the LLS for every timepoint at which measurements were made at altitude.
Furthermore, the incidence of AMS is compared using different measurement systems, on the one hand with the LLS described above, on the other hand with the environmental symptoms questionnaire (AMS-c). A cut-off of ≥ 0.7 for the AMS-c is used. Measurements occurred at the same timepoints as the LLS.
Additional LLS cut-offs and version will be compared. First the 2018 vs. 1993 LLS will be compared, as well as LLS ≥6 will represent the incidence of moderate-to-severe AMS.

Incidence and severity of side effects are evaluated using a standardised questionnaire during clinical examination. The severity of the following side effects was recorded on a scale from none over slightly and moderate to severe:
- tingling sensation
- impaired sense of taste
- increased urinary urge
- others (specified)

Data collection will be carried out at the same timepoints as the assessment of AMS and can be looked up in table 2 of the study protocol.

Vital parameters and parameters from arterial blood gas analysis are obtained. For the specific timepoints of measurement please refer to table 2 of the study protocol. It is planned to assess the difference in sex- and altitude-induced changes between the acetazolamide and placebo group of the following variables:

- Vital parameters
    - Arterial blood pressure [mmHg]
    - Heart rate [bpm]
    - Oxygen saturation ($SpO_2$) [%]
- Arterial blood gas and dried blood spot analyses
    - Arterial partial pressure of oxygen ($PaO_2$) [kPa]
    - Arterial partial pressure of carbon dioxide ($PaCO_2$) [kPa]
    - Arterial oxygen saturation ($SaO_2$) [%]
    - Bicarbonate [mM]
    - pH
    - Haemoglobin [g/l]
    - Haematocrit [%]
    - Acetazolamide plasma concentration

#### 7.2 Analysis methods

The primary outcome will be analysed in the intention-to-treat population, secondary outcomes in the per-protocol population. The primary analysis will be a modified Poisson regression with robust variance comparing the sex-related relative risk reduction with acetazolamide against AMS. For other AMS-related comparisons, $Chi^2$-square statistics and Cox-proportional hazard models will be applied.

For secondary outcomes such as vital parameters, results from arterial blood gases, mixed regression models will be fitted with sex, drug, altitude, and their interaction sex*drug*altitude as fixed effects, and participants as random effects. It is planned to present unadjusted models, if however, imbalances in baseline characteristics between the acetazolamide and placebo group are found, additional analysis with adjustment for these factors will be performed.
The incidence and severity of medication side effects will be evaluated using chi-square statistics or Fisher Exact tests.

#### 7.3 Missing data

Missing values in the primary outcome due to premature termination of the high-altitude stay will be imputed as being AMS positive. This approach is chosen to avoid overestimation of a potential treatment effect of acetazolamide. Sensitivity analyses of the primary outcome include the analysis in the per-protocol population. Missing values in secondary outcomes will not be replaced.

#### 7.4 Additional analyses

Analyses related to the primary outcome AMS, defined by the 2018 LLS, will be repeated for the 1993 LLS AMS definition. This analysis will allow to compare the novel findings with previously published AMS data.

#### 7.5 Harms

For every treatment arm the number and percentage of participants experiencing adverse events (or serious adverse events, if any) will be presented categorized by severity. No formal statistical testing will be undertaken.

#### 7.6 Statistical software

Statistical analyses will be performed with Stata version 14.2 or higher, and R software. Other packages may be used if necessary.
