## Supplementary Appendix 3 for "Sex-specific efficacy of acetazolamide for acute mountain sickness. A randomised clinical trial"

**Funding**

This study received funding from the Swiss National Sciences Foundation (10001630), the Heuberg Foundation (2023-009), and the Swiss Lung Foundation. Siemens Healthineers AG provided medical equipment for arterial blood gas analyses.

**Correspondence**

**Table of contents**


| | Page |
|---|---|
| **Supplementary methods** | 3 – 5 |
| **Table S1.** Predictors of acute mountain sickness, defined by the 2018 Lake Louise questionnaire – Cox proportional hazards analysis – per protocol analysis. | 6 |
| **Table S2.** Predictors of acute mountain sickness, defined by the 1993 Lake Louise questionnaire – Cox proportional hazards analysis – per protocol analysis. | 7 |
| **Table S3.** Acute mountain sickness symptoms and other intercurrent illness and/or symptoms requiring medical intervention and/or study termination based on the pre-specified safety regulations of the independent physician. | 8 |
| **Table S4.** Secondary outcomes, per-protocol population | 9 |
| **Table S5.** Acute mountain sickness symptom assessment at 760 m, on the morning of the ascent. | 10 |

### Supplementary methods

#### ***Questionnaire and evaluation***

AMS was assessed using the 2018 revised LLS,[10] which includes self-rated evaluation of four symptoms: headache, fatigue, gastrointestinal discomfort, and dizziness. Each symptom is scored from 0 (absent) to 3 (severe), yielding a total score ranging from 0 (no symptoms) to 12 (severe). For comparing the AMS incidences and severity with previous studies, the original LLS score, incorporating the fifth questionnaire item related to sleep disturbance, was assessed. The Environmental Symptoms Questionnaire cerebral score (AMSc) was also administered.[11] This instrument consists of 11 items assessing AMS-related symptoms, each rated on a scale from 0 (not at all) to 5 (extreme), yielding a weighted composite score from 0 (not at all) to 5 (extreme). A weighted AMSc score ≥0.7, is considered indicative of AMS. The AMS questionnaires were administered at baseline, after arrival and thereafter, twice daily, after awakening and in the evening. In case of symptom worsening, supplemental questionnaires were administered accordingly.

#### ***Safety considerations***

Due to the safety rules by the Kumtor Gold Facility, any participant presenting with a Lake Louise Score (LLS) of ≥3 received symptomatic treatment for AMS (ibuprofen 400 mg and/or antiemetics) upon request. If the LLS reached ≥6 or the participant presented any other health condition requiring medical intervention, supplemental oxygen (2–4 L/min via nasal cannula) or medical treatment was administered by an independent physician. The participant was subsequently scheduled for relocation to lower altitude on the next possible occasion.

#### ***Sample size estimation***

The RRR of acetazolamide for AMS was estimated to be approximately 44%.[7] The AMS incidence at 3600 m was assumed to be 50% in females and 30% in males.[13] A clinically meaningful sex-related RRR difference by acetazolamide was defined as 20%. Correspondingly, assuming a RRR of 50% in females (absolute risk reduction from 50% to 25%) and RRR of 30% in males (absolute risk reduction from 30% to 21%), with a 2:1 female-to-male ratio, a power of 90%, and a two-sided alpha level of 0.05, and accounting for potential dropouts, a total of 180 females and 90 males were initially planned for recruitment. The female-to-male ratio of 2:1 was selected to enable robust testing of additional secondary hypotheses within the female participants. Based on the pre-specified interim analysis, performed after the first study year, the recruitment target was increased to 180 female and 120 male participants.

#### *Interim analysis*

To minimise any unforeseen deviations from the anticipated study progression, such as extreme benefit or harm of acetazolamide or futility and unexpected AMS incidences, a planned interim analysis was scheduled after the completion of the first study year. Any premature termination of the study was defined by symmetric stopping boundaries at $p<0.001$ (Peto approach) on the primary outcome of the study. The planned interim analysis was performed by an independent statistician blinded to the drug assignment.

The blinded interim analysis was therefore conducted in January 2025. The analysis included 133 randomised participants. The observed incidence of AMS was 44% in females (instead of assumed 50%) and only 16% in males (instead of assumed 30%). Especially the low number of males (due to the pre-defined 2:1 ratio), combined with the lower-than-expected AMS incidence, resulted in a sample size re-estimation. A total of 180 female and 120 male participants were set as the new target sample size.

#### *Statistical Analysis*

In accordance with international guidelines for prospective clinical trials,[1] a statistical analysis plan (SAP) was created, and result reporting follows the CONsolidated Standards Of Reporting Trials (CONSORT) guidelines.[2] The study protocol and SAP versions are available in the **Supplementary Appendix 1;** the CONSORT checklist in the **Supplementary Appendix 2**. Data are summarized by numbers and proportions and means ± SD. The primary analysis was performed on the intention-to-treat population including all randomized participants using the modified Poisson regression with robust variance comparing the sex-related mean difference (95% CI) in the acetazolamide effect, reported as RRR, against AMS. In case of missing values in the primary outcome (AMS incidence), the missing data were imputed as AMS positive, for not overestimating the treatment efficacy.

Absolute risk reduction (ARR) was defined as the difference in outcome risk between treatment groups within each sex (acetazolamide vs. placebo). Sex differences in treatment effect were assessed as differences in ARR between females and males. Confidence intervals were obtained using non-parametric bootstrapping (20,000 resamples), with percentile-based 95% CI. The number needed to treat (NNT) was calculated using Bender's 95% CI. Kaplan-Meier curves were plotted for AMS, and Cox proportional-hazards analysis has been conducted to determine whether sex and acetazolamide therapy are independent risk factors. The appropriateness of the proportionality assumption was evaluated by visual inspection of Kaplan-Meier and log-log plots. As the proportionality assumption was not fulfilled, a time-varying covariate was introduced with two levels, separated at the time where the lowest model AIC was achieved (t <3h after arriving at altitude). Secondary outcomes were analyzed on the per-protocol population, defined as participants with available data. Continuous variables were analyzed using mixed linear regression models, with the variable of interest as the dependent variable and drug, location and sex as fixed effects, including the interaction term drug*location*sex. For improving the interpretability and comparability to other studies reporting AMS, different AMS definitions were calculated, among them, 2018 and 1993 LLQ score of ≥6, and an AMSc score ≥0.7. Comparisons of the AMS incidences between females and males within drugs were performed using Chi-Square statistics or Fisher Exact tests. Analyses were conducted using R software (version 4.3+). A $P<0.05$ or 95% CI excluding zero were considered to indicate statistical significance. In secondary outcomes, mean differences with 95% CI are presented without adjustment for multiplicity and without P values.

**Supplementary Tables**

**Table S1. Predictors of acute mountain sickness, defined by the 2018 Lake Louise questionnaire – Cox proportional hazards analysis – per protocol analysis.**

| Predictor | Hazard ratio | SE | 95% CI | P value |
|---|---|---|---|---|
| Drug effect in males, acetazolamide vs. placebo, ≥3 hours after arrival at 3600 m | 0.46 | 0.14 | 0.25 to 0.82 | 0.009 |
| Sex effect with placebo, females versus males | 2.57 | 0.69 | 1.52 to 4.34 | <0.001 |
| Time-varying covariate, <3 versus ≥3 hours after arrival at 3600 m[a] | 2.65 | 1.26 | 1.05 to 6.73 | 0.040 |

[a]As the proportionality assumption for the Cox regression analysis was not fulfilled (see Figure 2), a time-varying covariate with two levels (<3 versus ≥3 hours after arrival at 3600 m) was introduced.

**Table S2. Predictors of acute mountain sickness, defined by the 1993 Lake Louise questionnaire – Cox proportional hazards analysis – per protocol analysis.**

| Predictor | Hazard ratio | SE | 95% CI | P value |
|---|---|---|---|---|
| Drug effect in males, acetazolamide vs. placebo, ≥3 hours after arrival at 3600 m | 0.49 | 0.14 | 0.29 to 0.84 | 0.010 |
| Sex effect with placebo, females versus males | 2.44 | 0.61 | 1.50 to 3.98 | <0.001 |
| Time-varying covariate, <3 versus ≥3 hours after arrival at 3600 m[a] | 2.45 | 1.12 | 1.00 to 6.01 | 0.050 |

[a]As the proportionality assumption for the Cox regression analysis was not fulfilled (see Figure 2), a time-varying covariate with two levels (<3 versus ≥3 hours after arrival at 3600 m) was introduced.

**Table S3. Acute mountain sickness symptoms and other intercurrent illness and/or symptoms requiring medical intervention and/or study termination based on the pre-specified safety regulations of the independent physician.**

| Time of event | Sex | Treatment group | Symptoms and signs | Actions taken | Outcome |
|---|---|---|---|---|---|
| Day 1 evening | female | Placebo | AMS[1] (LLS[2]=5), Severe nausea | Oxygen, premature study termination | Full recovery <3 h after oxygen |
| Day 1 evening | female | Placebo | AMS (LLS=4), vomiting | Oxygen | Full recovery <3 h after oxygen |
| Day 1 evening | female | Placebo | AMS (LLS=6), dyspnoea | Paracetamol 500 mg, oxygen (nights 2–3) | Full recovery <3 h after oxygen |
| Day 1 evening | male | Placebo | AMS (LLS=7), hypotension | Oxygen | Full recovery <3 h after oxygen |
| Day 3 morning | female | Placebo | AMS (LLS=3), syncope | Oxygen, Paracetamol 500 mg | Full recovery <3 h after oxygen |
| Day 1 evening | female | Acetazolamide | Panic attack, stomach pain, dyspnoea | Salbutamol 300 µg, omeprazole 20 mg | Full recovery <3 h after oxygen |
| Day 1 evening | female | Acetazolamide | LLS = 0, AMSc[3]=1.9 | Oxygen (decision of chief medical physician) | Full recovery <3 h after oxygen |
| Day 2 evening | female | Acetazolamide | Vomiting | No specific treatment | Full recovery before descent |
| Day 2 morning | female | Acetazolamide | AMS (LLS=4), vomiting | No specific treatment | Full recovery before descent |
| Day 2 morning | male | Acetazolamide | Severe abdominal/back pain, vomiting, microhematuria (ureteral stone) | Paracetamol, spasmolytics, painkiller i.m.; medical evacuation | Kidney stone treated in lowland clinic; discharged day 5 |

[1]AMS=acute mountain sickness; [2]LLS= Lake Louise acute mountain sickness score ranging from 0 to 12 points (absent to severe).
[3]AMSc score= Environmental Symptoms questionnaire cerebral score ranging from 0 to 5 points (not at all to severe)

**Table S4. Secondary outcomes, per-protocol population**

| Variable | Placebo | | | | Acetazolamide | | | |
|---|---|---|---|---|---|---|---|---|
| | Males | | Females | | Males | | Females | |
| | 760 m | 3600 m | 760 m | 3600 m | 760 m | 3600 m | 760 m | 3600 m |
| **Clinical examination** | | | | | | | | |
| Heart rate, bpm | 75.5 ± 1.3 | 75.2 ± 1.3 | 81.2 ± 1.1 | 85.7 ± 1.1 | 78.2 ± 1.3 | 77.3 ± 1.3 | 77.9 ± 1.1 | 84.1 ± 1.1 |
| $SpO_2$, % | 95.9 ± 0.3 | 87.3 ± 0.3 | 96.4 ± 0.2 | 87.5 ± 0.2 | 95.8 ± 0.3 | 89.9 ± 0.3 | 96.1 ± 0.2 | 89.7 ± 0.2 |
| Systolic BP, mmHg | 122 ± 1 | 125 ± 1 | 109 ± 1 | 112 ± 1 | 121 ± 1 | 124 ± 1 | 110 ± 1 | 110 ± 1 |
| Diastolic BP, mmHg | 77 ± 1 | 86 ± 1 | 73 ± 1 | 80 ± 1 | 76 ± 1 | 84 ± 1 | 74 ± 1 | 77 ± 1 |
| **Arterial blood gases** | | | | | | | | |
| pH | 7.41 ± 0.00 | 7.43 ± 0.00 | 7.42 ± 0.00 | 7.43 ± 0.00 | 7.41 ± 0.00 | 7.38 ± 0.00 | 7.42 ± 0.00 | 7.37 ± 0.00 |
| $PaCO_2$, kPa | 5.2 ± 0.1 | 4.6 ± 0.0 | 4.9 ± 0.0 | 4.2 ± 0.0 | 5.3 ± 0.1 | 4.0 ± 0.1 | 5.0 ± 0.0 | 3.7 ± 0.0 |
| $PaO_2$, kPa | 12.2 ± 0.1 | 7.4 ± 0.1 | 12.4 ± 0.1 | 7.6 ± 0.1 | 12.2 ± 0.1 | 8.5 ± 0.1 | 12.3 ± 0.1 | 8.5 ± 0.1 |
| $SaO_2$, % | 96.9 ± 0.3 | 89.2 ± 0.3 | 97.2 ± 0.2 | 89.9 ± 0.2 | 97.2 ± 0.3 | 91.5 ± 0.3 | 97.3 ± 0.2 | 91.7 ± 0.2 |
| Hematocrit, % | 43 ± 1 | 44 ± 0 | 36 ± 0 | 36 ± 0 | 43 ± 0 | 46 ± 0 | 34 ± 0 | 39 ± 0 |
| Hemoglobin conc., g/L | 146 ± 1 | 149 ± 1 | 121 ± 1 | 123 ± 1 | 145 ± 1 | 156 ± 1 | 117 ± 1 | 132 ± 1 |
| $HCO_3^-$, mmol/l | 25.1 ± 0.3 | 23.0 ± 0.2 | 23.7 ± 0.2 | 21.2 ± 0.2 | 25.6 ± 0.3 | 17.7 ± 0.3 | 24.1 ± 0.2 | 16.0 ± 0.2 |

Data are presented as mean ± SE from 760 m and the morning following the first night at 3600 m. AMS, acute mountain sickness; $SpO_2$, arterial oxygenation assessed by finger pulse oximetry; $PaCO_2$, partial pressure of arterial $CO_2$; $PaO_2$, partial pressure of arterial $O_2$, $SaO_2$, arterial oxygen saturation; $HCO_3^-$, bicarbonate.

**Table S5. Acute mountain sickness symptom assessment at 760 m, on the morning of the ascent.**

| | Placebo | | Acetazolamide | |
|---|---|---|---|---|
| **Variable** | **Males** | **Females** | **Males** | **Females** |
| **Primary outcome, n ITT** | 64 | 91 | 59 | 89 |
| 2018 LLQ ≥3, n ITT (%) | 1 (2%) | 0 (0%) | 0 (0%) | 0 (0%) |
| **Secondary AMS definitions, n PP** | 60 | 90 | 54 | 87 |
| 2018 LLQ ≥3 - 5, n PP (%) | 0 (0%) | 0 (0%) | 0 (0%) | 0 (0%) |
| 2018 LLQ ≥6, n PP (%) | 0 (0%) | 0 (0%) | 0 (0%) | 0 (0%) |
| 1993 LLQ ≥3, n ITT (%) | 1 (2%) | 3 (3%) | 0 (0%) | 0 (0%) |
| 1993 LLQ ≥3-5, n PP (%) | 0 (0%) | 0 (0%) | 0 (0%) | 0 (0%) |
| 1993 LLQ ≥6, n PP (%) | 0 (0%) | 0 (0%) | 0 (0%) | 0 (0%) |
| AMSc score[1] ≥0.7, n PP (%) | 0 (0%) | 0 (0%) | 0 (0%) | 0 (0%) |

Values are presented as numbers (proportions). AMS, acute mountain sickness; LLQ, Lake Louise Questionnaire; ITT, intention-to-treat; PP, per protocol; RRR, relative risk reduction calculated by a modified Poisson regression with robust variance. [1]AMSc score= Environmental Symptoms questionnaire cerebral score ranges from 0 to 5 points (not at all to severe).
